# Clinical outcomes of antimicrobial resistance in cancer patients: A systematic review of multivariable models

**DOI:** 10.1101/2022.07.15.22277667

**Authors:** Anders Skyrud Danielsen, Léa Franconeri, Samantha Page, Anders Eivind Myhre, Ragnhild Agathe Tornes, Oliver Kacelnik, Jørgen Vildershøj Bjørnholt

## Abstract

**Background:** Infections are major causes of disease in cancer patients and pose a major obstacle to the success of cancer care. The global rise of antimicrobial resistance threatens to make these obstacles even greater and hinder continuing progress in cancer care. To prevent and handle such infections, better multivariable models building on current knowledge are needed. This internally funded systematic review (PROSPERO registration: CRD42021282769) aimed to review multivariable models of resistant infections/colonisations and corresponding mortality, what risk factors have been investigated, and with what methodological approaches.

**Methods:** We employed two broad searches of antimicrobial resistance in cancer patients, using terms associated with antimicrobial resistance, in MEDLINE and Embase through Ovid, in addition to Cinahl through EBSCOhost and Web of Science Core Collection. Primary, observational studies in English from January 2015 to November 2021 on human cancer patients that explicitly modelled infection/colonisation or mortality associated with antimicrobial resistance in a multivariable model were included. We extracted data on the study populations and their malignancies, risk factors, microbial aetiology, and methods for variable selection, and assessed the risk of bias using the NHLBI Study Quality Assessment Tools.

**Results:** Two searches yielded a total of 27151 unique records, of which 144 studies were included after screening and reading. Of the outcomes studied, mortality was the most common (68/144, 47%). Forty-five per cent (65/144) of the studies focused on haemato-oncological patients, and 27% (39/144) studied several bacteria or fungi. Studies included a median of 200 patients and 46 events. One-hundred-and-three (72%) studies used a p-value-based variable selection. Studies included a median of seven variables in the final (and largest) model, which yielded a median of 7 events per variable. An in-depth example of vancomycin-resistant enterococci was reported.

**Conclusions:** We found the current research on this topic to be heterogeneous, in both the methodological and epidemiological approaches. Methodological choices resulting in very diverse models made it difficult or even impossible to draw statistical inferences and summarise what risk factors were of clinical relevance. The development and adherence to more standardised protocols that build on existing literature are urgent.

## Background

Cancer patients have a higher risk and worse outcomes of infectious disease complications, compared with healthy people [1]. Autopsy studies have indicated that infections may play a role in more than half of all cancer patient fatalities [1]. A recent study has also shown that cancer patients have a higher risk of contracting infections with antimicrobial-resistant organisms [2]. Importantly, infections often necessitate caregivers to postpone or withhold adequate cancer treatment, which may impair cancer outcomes. In recent years, there have been major changes in disease-causing microbial ecology, particularly in hospitals [3]. Bacteria and fungi are becoming increasingly resistant to antimicrobial drugs, and in Europe alone it is estimated that more than 33,000 people die each year from resistant microbes [4,5]. Not only are microbes acquiring antimicrobial resistance, but the microbial spectrum is changing, with an increasing proportion of species with a propensity for intrinsic resistance [6]. Infections in cancer patients are increasingly often caused by resistant organisms, which threaten recent years’ advances in the treatment of cancer [7]. In other words, there is a pressing need to understand the changing epidemiology of bacterial and fungal infections among cancer patients, but also to design better preventive measures.

To adapt to this new reality, it is essential to understand the mechanisms by which infections occur and cause disease. The research that leads to the discovery and description of these mechanisms, as well as testing them in preliminary models, has been called ‘prognostic factor research’ by the PROGRESS Group [8]. Prognostic factor research forms the basis for more advanced risk stratification tools and scoring systems clinicians may use to guide anti-infective therapy. Such risk stratification tools or scoring systems are usually based on clinical prediction models, which are typically regression models where individual-level clinical data are used to predict a clinical outcome of interest [9,10]. Some examples widely used in the management of infectious complications in cancer patients are the Multinational Association for Supportive Care in Cancer (MASCC) risk indices for febrile neutropenic patients or the Pitt bacteraemia score. These risk indices were developed in relatively small patient cohorts in the early 1990s [11,12].

Because of the changing epidemiology, there is a need to update the multivariable regression models that estimate and predict the risks associated with antimicrobial resistance in cancer care, like the additional risk of death attributable to resistance. In this work, it is important to build on already existing multivariable models and use factors that previously have been shown to be associated with the outcomes of interest. It is thus necessary to map the existing literature on such multivariable models to facilitate the use of current knowledge in future research. In this systematic review, we aimed to review multivariable models of resistant infections/colonisations and corresponding mortality in cancer patients, what risk factors have been included, and with what methodological approaches.

## Methods

### Protocol registration and reporting standards

To review what risk factors for resistant infections and/or carriage/colonisation (hereafter “infections/colonisations”) and corresponding mortality in cancer patients have been investigated, we conducted a systematic review employing a broad and extensive search for literature published from 1^st^ of January 2015 to 19^th^ of November 2021. It was not possible to separate studies on infection and colonisation, so these outcomes were combined. Although first coined in 1961, the term ‘risk factor’ remains an elusive term as it is used to describe any covariate associated with an outcome [13,14]. We will here use the term ‘risk factor’ colloquially as a common designation for both causal risk factors and predictive risk markers, as this terminology is often used in the primary studies included [15]. This systematic review was registered at PROSPERO (ID: CRD42021282769) [16] and follows the Preferred Reporting Items for Systematic Reviews and Meta-Analyses (PRISMA) guideline (checklist can be found in Supplementary Material 7) [17].

### Search strategy

Our search strategy was implemented in three steps. First, we performed preliminary searches in PubMed to identify some main keywords to be included and to get an overview of the size of the literature. We then performed a first search and, after screening and sorting the results of the search, we manually reviewed the records and references to discover keywords that were not covered by our initial search. We then performed a second search designed to expand the first search findings.

The first search was conducted by a research librarian from 22nd to 24th June, 2021. It consisted of terms covering all cancers and terms covering antibiotic resistance and infections. A spectrum of synonyms with appropriate truncations and proximity operators was used for searching title, abstract, and author keywords. In addition, controlled subject headings were searched when available. The search strategy was tailored to each database’s search interface. The search was run in the OVID MEDLINE and OVID Embase, in addition to EBSCO Cinahl and Web of Science Core Collection (Science Citation Index Expanded, Social Sciences Citation Index, Arts & Humanities Citation Index and Emerging Sources Citation Index - searched simultaneously). The strategies were limited to Danish, English, Norwegian, Spanish, and Swedish. They also included a time limit for publications from the 1^st^ of January 2015 onwards. A total of 25 881 records were retrieved. After removing duplicate records in EndNote, 14 153 references were identified.

The second search expanded on this by including specific antibiotics, resistance mechanisms, and bacteria and fungi often associated with either acquired resistance or high levels of intrinsic resistance. The terms covering antibiotics were variations of piperacillin/tazobactam, meticillin/methicillin, cephalosporin, carbapenem, aminoglycoside, gentamicin, amikacin, fluoroquinolone, linezolid, vancomycin, echinocandin, azole, colistin. The terms covering mechanisms of resistance or microbial properties associated with resistance were beta-lactamase/β-lactamase, extended-spectrum beta-lactamase/β---lactamase/betalactamase, carbapenemase, biofilm-producing, non-fermenting. The terms covering bacteria and fungi were *Pseudomonas aeruginosa, Acinetobacter spp., Acinetobacter baumannii, Stenotrophomonas maltophilia, Clostridium/Clostridioides difficile, Enterococcus faecium, Enterococcus faecalis*, coagulase-negative staphylococci, Candida non-albicans, *Candida auris, Aspergillus fumigatus*. Those were selected from a wide range of terms during a multidisciplinary meeting which led to a consensus. This search was run on 19^th^ of November 2021 in the same databases (except for Cinahl) and with the same limitations as the June version.

Search strategies can be found in Supplementary Materials 1 and 2.

### Study selection

The titles of all records were first screened by AD and LF to exclude any records that were not about antimicrobial resistance and cancer using the Rayyan tool [18]. We then screened all abstracts of the records where the subject was antimicrobial resistance and cancer to sort these into different study designs. The remaining original records with an observational study design were then read in full text by both AD and LF to see if they matched the eligibility criteria (Table 1). If there was uncertainty about the inclusion of a record, both authors discussed it until reaching a decision.

**Table 1.** Eligibility criteria.

| Inclusion criteria | Exclusion criteria |
| --- | --- |
| • Primary study with original data<br>• Observational study design (prospective/retrospective cohort study, case-control study, cross-sectional study)<br>• The study population consists only of cancer patients, haematological or otherwise, of any age, sex, or ethnicity, and in any institution in any country.<br>• The study includes a multivariable model.<br>• A resistant bacterial or resistant fungal infection is either a risk factor or an outcome<br>• The outcome is either infection (or colonisation) or death<br>• Published after 1st of January 2015.<br>• Published in English.<br>• Studies in humans. | • Any other publication type than a primary observational study.<br>• Cancer is a covariate or comparator, not everyone in the study population has cancer.<br>• Resistance to antimicrobial drugs is not included in the model (neither as a risk factor nor as an outcome).<br>• The outcome is not infection or mortality, e.g. distribution of bacteria or bacterial strains.<br>• Studies where the causal assumption is reversed, so that infection causes cancer (e.g. increased risk of gastric cancer after *Helicobacter pylori* infection) |

### Data extraction and statistical analysis

#### Data extraction

The risk of bias in all included studies was assessed by AD and LF separately in a blinded process using the National Institutes of Health study quality assessment tools [19], and an arbitration meeting was held where OK/JB acted as an arbiter to come to a consensus about the final risk of bias assessment. The guidance for the use of these tools was followed, but the tool was modified to include an item with a question of whether the outcome was well defined in case-control studies.

Data extraction was then performed by AD and LF. We extracted the year of publication, the title of the study, the authors, the number of patients, the country of the study setting, the study aim statement, the patient population statement, the risk factors included in the final model, the microbial aetiology, whether the studies employed a screening for statistical significance, and the events per variable in the final model. This was included together with the risk of bias assessment and a short commentary in three different tables of all included studies, one for studies with an infection/colonisation outcome, one for studies with a mortality outcome and one for studies with both outcomes. These three tables may be found in the Supplementary Material S3, S4, and S5. The studies were checked against pre-specified criteria for a potential meta-analysis, being that models would have the same aetiology, outcome, and risk factors. These criteria were not met for any aetiology. To provide a genuine example of the heterogeneity of the models, population, variables and outcomes investigated, a qualitative in-depth example of the investigation of risk factors was reported. We chose researches that included vancomycin-resistant enterococci (VRE), as they represented a typical cross-section of the studies selected. Furthermore, VRE is a typical hospital-associated microbe that may readily be prevented.

#### Statistical analysis

A table describing the outcomes and extracted data on the country, microbial aetiology, patients, events, events per variable, variables screened, variables in the final model, and p-value-based variable selection were created. The table summarised the findings by presenting frequencies with percentages for categorical variables and medians with interquartile ranges for continuous variables. To summarise what risk factors have been investigated, we grouped the microbial aetiology into five large groups (*Clostridioides difficile*, fungi, Gram-negative bacteria, Gram-positive bacteria, and several bacteria/fungi) and categorised the risk factors. We then created a table of how many times the different types of risk factors had been included in the final multivariable model in the included studies for each of the large microbial aetiology groups. A full list of the risk factors and their respective categories can be found in the appendix. Analyses were performed in R using RStudio version 4.1.1 [20], and the scripts used to produce the results can be found on <u>GitHub</u> [21].

## Results

### Study selection

After excluding 47048 duplicates, the two searches yielded a total of 27151 unique records from a total of three major databases - Ovid MEDLINE, Ovid Embase, and Web of Science - in addition to the Cinahl database (EBSCO) (Figure 1). Title screening for relevance to both antimicrobial resistance and cancer excluded 25341 records, whereas most excluded records were only about cancer, e.g. basic research on the drug resistance of the cancer disease. Abstract screening for a non-observational study design excluded a further 845 records, where 103 had a review design, 50 had an interventional design, 454 were case reports, and 238 were miscellaneous, mostly commentary articles and conference abstracts. Finally, after full-text screening; 821 records were excluded, of which 165 included other diagnoses than cancer (including healthy individuals), 197 did not include an infection/colonisation or mortality outcome (often microbial distribution or endpoints like the length of stay), 130 did not explicitly include antimicrobial resistance in the model, 311 did not include a multivariable model, 16 were not in English, and we failed to gain access to two leaving 144 studies included.

**Figure 1:**
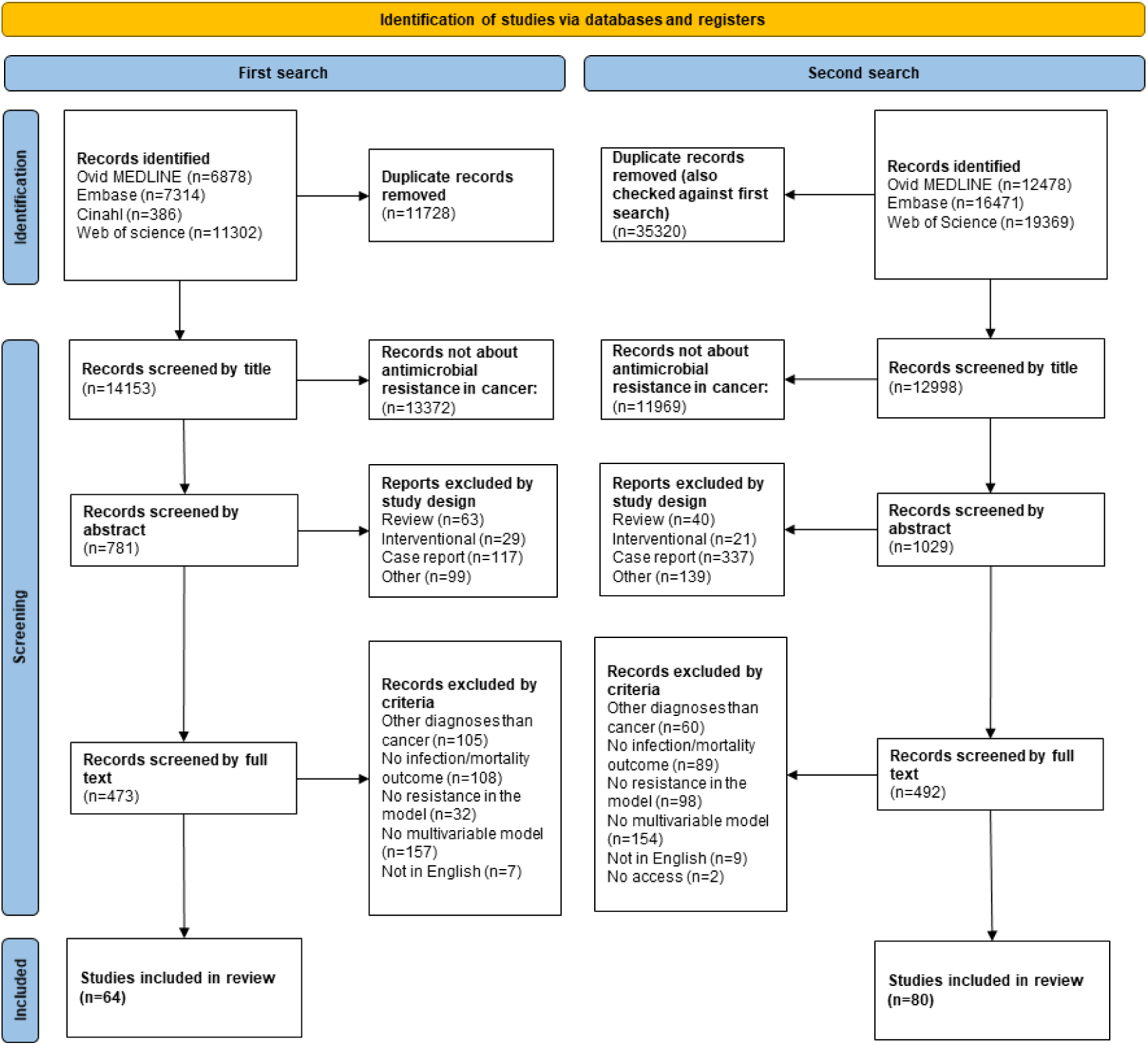
PRISMA flowchart of the study selection [17].

### Study characteristics

Of the 144 studies, 55/144 (38%) had an infection/colonisation outcome, 66/144 (46%) had a mortality outcome, and 23/144 (16%) had both outcomes, all of which are listed in detail and cited in tables with all extracted data, including the investigated risk factors, in Supplementary Material S3, S4, and S5, respectively. In total, there were 23/144 (16%) studies of patients with solid cancers [22–44], 65/144 (45%) studies of patients with haematological cancers [45–109], and 56/144 (39%) studies with patients of both or unspecified cancer types [110–165]. Most studies selected (39/144, 27%) reported and modelled several bacteria and/or fungi that were tested for resistance towards several antimicrobials [22–37,45–57,110–119]. Eight of 144 (6%) reporting and modelling several microorganisms focused only on Gram-negative bacteria [58–62,120–122] and 1/144 (1%) focused only on fungi [63]. Twelve of 144 (8%) studies studied the family *Enterobacterales* (especially *Escherichia coli* and *Klebsiella pneumoniae* combined), either ESBL- or carbapenemase-producing [38,64–68,123–128]. The most commonly studied single organism was *Clostridioides difficile* with 27/144 (19%) studies [39–41,69–81,129–139]. Also studied were three non-fermenters with 5/144 (3%) studies about *Acinetobacter baumannii* [82,140–143], 4/144 (3%) studies about *Pseudomonas aeruginosa* [83,84,144,145], and 5/144 (3%) studies about *Stenotrophomonas maltophilia* [85–88,146], often specified as either multidrug- or extensively drug-resistant. We also found several studies focusing on four well-known healthcare-associated bacteria, with 11/144 (8%) focusing on VRE [89–98,147,148], 7/144 (5%) focusing on carbapenemase-producing *K. pneumoniae* (KPC) [99–102,149–151], 2/144 (1%) focusing on methicillin-resistant *Staphylococcus aureus* (MRSA) [42,152], and 6/144 (4%) focusing on extended-spectrum beta-lactamase-producing *Escherichia coli* (ESBL-E) [103,104,153–156]. We also found that fungi typically resistant to antifungals were studied, 3/144 (2%) studies about *Aspergillus* spp. [43,105,106] and 9/144 (6%) studies about *Candida* non-albicans [44,107,157–163], respectively. Finally, there was one of 144 (1%) studies for each of the microbes *Staphylococcus epidermidis* (with linezolid-resistance) [108], *Bacillus* spp. [164], *Streptococcus pneumoniae* (several resistance mechanisms) [165], and *Aeromonas sobria* [109].

### Summary of findings

The most common microbial aetiologies were several bacteria/fungi (39, 25%), followed by *C. difficile* (27, 19%) and Enterobacteriaceae (12, 8%) (Table 2). The most common country for the study setting was the United States of America (USA) (40, 28%), followed by China (15, 10%), and Italy (11, 8%). The selected studies included a median of 200 patients (IQR 102-338) and 46 events (IQR 25-83.5). 103 (72%) studies used a p-value-based variable selection, either bivariable screening or stepwise regression. These studies screened a median of 16 variables (IQR 9-27) and included a median of 7 variables in the final (and largest) model. The median events per variable were close to 7.

**Table 2.**
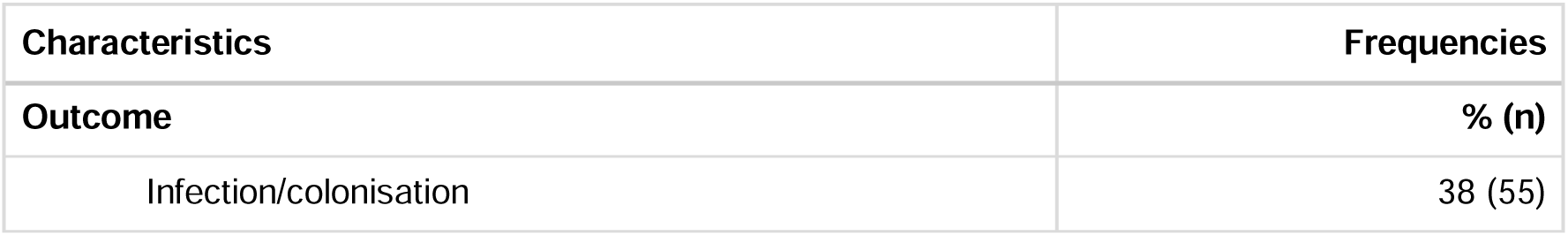
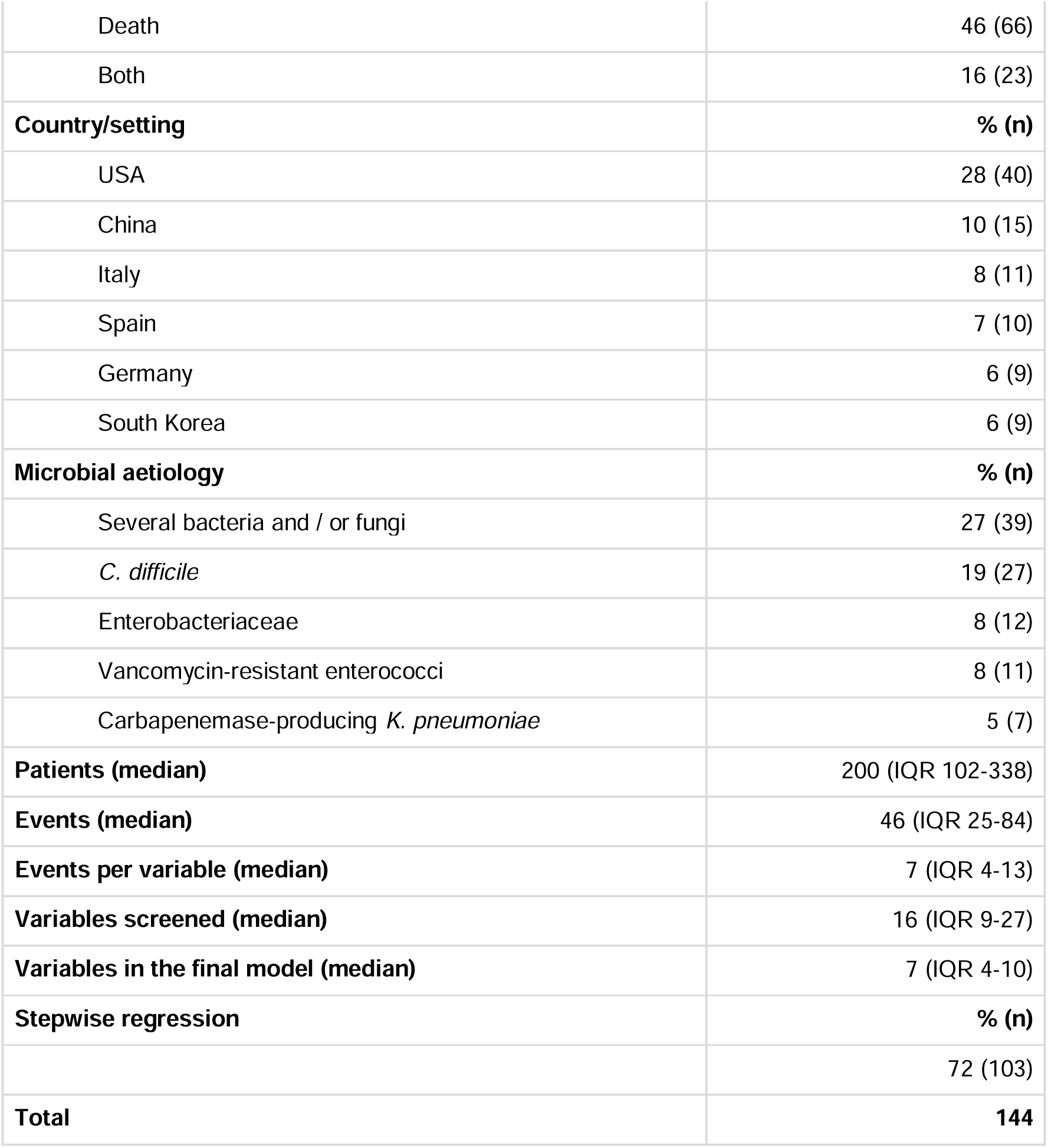
Summary of findings.

The most commonly investigated risk factors in models of resistant infection and/or colonisation in cancer patients were antibiotic use, with a total of 118 occurrences in the included studies (Table 3). In the models of mortality, however, the most commonly investigated risk factors were related to infection with a total of 208 occurrences. A full list of the risk factors and their respective categories may be found in Supplementary material S6.

**Table 3.** Categories of risk factors investigated in the final, multivariable model in the included studies, by large groups of microbial aetiology.

| **Infection/colonisation** |  | **Mortality** |  |
| --- | --- | --- | --- |
| **Risk factors by microbial aetiology** | **Count** | **Risk factors by microbial aetiology** | **Count** |
| ***Clostridioides difficile*** |  | ***Clostridioides difficile*** |  |
| Basic characteristics | 29 | Basic characteristics | 10 |
| Antibiotic use | 28 | Comorbidities/clinical condition | 8 |
| Cancer-related | 18 | Hospital-related | 6 |
| Comorbidities/clinical condition | 14 | Infection-related | 6 |
| Hospital-related | 14 | Other | 3 |
| **Fungi** |  | **Fungi** |  |
| Antifungal use | 7 | Infection-related | 21 |
| Basic characteristics | 7 | Hospital-related | 9 |
| Other | 6 | Basic characteristics | 7 |
| Infection-related | 6 | Comorbidities/clinical condition | 7 |
| Cancer-related | 4 | Antibiotic use | 6 |
| **Gram-negative bacteria** |  | **Gram-negative bacteria** |  |
| Antibiotic use | 65 | Infection-related | 71 |
| Infection-related | 35 | Comorbidities/clinical condition | 40 |
| Hospital-related | 23 | Cancer-related | 31 |
| Basic characteristics | 21 | Antibiotic use | 27 |
| Cancer-related | 15 | Basic characteristics | 24 |
| **Gram-positive bacteria** |  | **Gram-positive bacteria** |  |
| Antibiotic use | 14 | Infection-related | 14 |
| Infection-related | 11 | Cancer-related | 11 |
| Cancer-related | 7 | Basic characteristics | 8 |
| Basic characteristics | 5 | Neutropenia | 6 |
| Chemotherapy/immunosuppressants | 5 | Antibiotic use | 5 |
| **Several bacteria and/or fungi** |  | **Several bacteria and/or fungi** |  |
| Transplantation-related | 5 | Infection-related | 96 |
| Antibiotic use | 4 | Comorbidities/clinical condition | 39 |
| Laboratory findings (non-microbiological) | 4 | Cancer-related | 31 |
| Surgery-related | 4 | Basic characteristics | 31 |
| Comorbidities/clinical condition | 2 | Antibiotic use | 27 |

### In-depth example of VRE

As a model organism, we explored how risk factors for and of VRE in cancer patients were described and modelled in the selected studies. Of our selection, 46/144 (32%) of the studies mention either *Enterococcus* spp., *E. faecium*, or *E. faecalis*, which is either described as vancomycin-resistant or “multidrug-resistant” (sometimes not further specified), or where the resistance mechanism is not described at all. Most of the studies (30/46, 65%) that mention the organism include it in a larger group, like Gram-positive bacteria or multidrug-resistant organisms, which hampers any specific focus on VRE [22–34,45–53,110–117].

Eight studies model the risk of being either colonised or infected with VRE. The number of patients included ranged from 72 to 342, and the variables screened ranged from 6 to 46. One study focused on patients undergoing liver transplantation for hilar cholangiocarcinoma, and seven focused on haematological cancer patients. Although a plethora of risk factors was investigated, five of the studies selected variables based on their corresponding p-values in bivariable analyses, and as such dropped several variables from the analysis due to their failure to achieve the p-value criterion. The authors did not necessarily conclude that all risk factors investigated in the multivariable model were of importance. The risk of bias was rated low for three, medium for five, while none had a “fatal flaw”. However, five of eight studies had less than ten events per variable in the final model, which may lead to a lack of statistical power. Among the common conclusions from the studies of VRE infection or colonisation, four concluded that antibiotic exposure was a risk factor for VRE infection or colonisation, and two highlighted neutropenia. None of the antibiotic exposure risk factors was the same (one was vancomycin, one was carbapenem, one was general antibiotic exposure and one was daptomycin). When looking at the findings in detail, Aktürk et al. found that severe neutropenia and previous bacteraemia with another pathogen may increase the risk of progressing from VRE colonisation to VRE infection in paediatric haematological cancer patients [147]. In 2015, Ford et al. found that severe neutropenia and the number of stools per day were associated with VRE bloodstream infections in leukaemia patients, and in 2019 some of the same authors concluded that VRE colonisation rates fell when the hospital started using less carbapenem [89,90]. Herc et al reported that only previous daptomycin exposure was associated with daptomycin-resistant VRE infections in patients with haematological malignancies [91]. Hefazi et al found that VRE colonisation is associated with VRE infection in stem cell transplantation patients and Ramanan et al found that VRE colonisation pre-transplantation was associated with any infection post-transplantation [35,92]. Heisel et al found that cephalosporin use and intravenous vancomycin were associated with VRE infections in patients with acute myeloid leukaemia or myelodysplastic syndrome undergoing intensive induction therapy, and finally, Klein et al found that in multiple myeloma patients, granulocyte-colony stimulating factor was associated with fewer VRE cases than antibiotic prophylaxis [93,94].

Another eight studies analyse the deaths associated with VRE in cancer patients. The number of patients included ranged from 95 to 1424, and the variables screened ranged from 11 to 56, although the exact number was indeterminable for one of the studies. Six studies selected variables based on a specified p-value threshold, but for one study the method for variable selection could not be determined. We assessed the risk of bias among these studies and found four studies at low risk and four studies at a medium risk of bias. Of the eight studies, six had less than 10 events per variable in the final model. The studies had few conclusions about risk factors for mortality among cancer patients associated with VRE in common. However, three studies found that VRE bacteremia was a risk factor for death and two studies found no risk factors after running their model. When looking at the findings in detail, Akhtar et al. found that only shock (not further specified) was associated with the difference in mortality between VRE and VSE bacteraemia in cancer patients [148]. Kamboj et al did not find any factors that were associated with higher mortality in stem cell transplantation patients with VRE bacteraemia [95]. Kern et al modelled the mortality associated with enterococcal bacteraemia in haematological cancer patients but did not specify vancomycin resistance [54]. Kirkizlar et al found that in leukaemia patients colonised with VRE, a low neutrophil count and coinfection were associated with increased mortality [96]. Mendes et al included VRE in a bivariable screening but discarded the factor as it did not meet the criterion of p<0.1 [55]. Ornstein et al found that leukaemia patients with a VRE bacteraemia at the induction of chemotherapy had poorer survival than patients with other bloodstream infections [97]. Papanicolaou et al found that VRE bacteraemia increased the mortality in patients receiving their first stem cell transplantation, but did not disclose how variables were selected for the multivariable model [98]. Finally, Pugliese et al modelled the risk of mortality associated with several bacteria in leukaemia inpatients, among them *Enterococcus* spp. (no vancomycin resistance mentioned), but did not find an association [56].

## Discussion

In our systematic review of studies with multivariable models with risk factors for infection/colonisation and mortality associated with antimicrobial resistance in cancer patients, we selected 144 studies that were eligible for inclusion. Most studies focused on haematological cancer patients and explored a host of different microbes. Studies on infection/colonisation with resistant microbes as outcomes most often investigated risk factors relating to antibiotic use, while studies with mortality as an outcome often included risk factors relating to the infection itself. Studies often had small sample sizes, screened a large number of variables, and used p-value-based methods for variable selection like bivariable screening or stepwise regression. In general, the models were highly heterogeneous, with nuances in the study populations and microbial aetiologies, and major differences in which risk factors were modelled, as we have exemplified through the research on VRE in cancer patients.

The issues with heterogeneity when performing a systematic review of basic prognostic factor research have been described before [166]. Although it is not possible to infer which factors are of importance, we find that known general risk factors for infections like immunosuppression and specific risk factors for resistant infections like previous antibiotic use are recurring, as other non-systematic reviews do [1,2,6,7]. A comprehensive list of these factors can be found in the supplementary material. To the best of our knowledge, no previous systematic review has investigated the risk factors for all resistant infections in cancer patients. However, three systematic reviews have looked at risk factors for methicillin-resistant *S. aureus*, extended-spectrum β lactamase-producing *Enterobacteriaceae*, and vancomycin-resistant enterococci, specifically [167–169]. These studies were able to pool the prevalence of such infections and identify some risk factors of importance, providing a good basis for future research. Some systematic reviews also investigate the changing epidemiology of antimicrobial-resistant infections in cancer patients without a particular focus on risk factors [6,170].

Some conservative choices were made in the selection process which may have reduced the final number of studies included. Some studies were excluded because study participants had other diagnoses than cancer, e.g. recipients of hematopoietic stem cell transplantation due to non-malignant haematological disorders [171,172]. Several of these studies did not describe the full diagnostic panorama, although comprehensive patient characteristics remain an important backbone in epidemiological research. Other studies did not explicitly model resistant microbes by including resistance as a variable or an outcome [173,174]. Given the rapid increase in antimicrobial resistance, current research in the epidemiology of infectious diseases in healthcare settings should include detailed data on antimicrobial resistance. The most common reason for excluding observational studies during full-text reading was the lack of a multivariable model [175,176]. These studies seemed to have a low number of patients, and a multivariable model may have been avoided due to low statistical power. As discussed by the PROGRESS Research Group in their recommendations for prognostic factor research, the discovery and investigation into new prognostic factors should rely on multivariable modelling to discern these from factors already known to be of importance, and to provide a basic adjustment of confounders [8].

Studies simply testing whether factors differ between groups often rely heavily on null-hypothesis statistical significance testing and are subject to the multiple comparison problem [177]. Although exploratory studies are important, such studies sometimes either test risk factors that are already known to be associated with the outcome, test factors in too small samples, or test too many factors at once. This may lead to wrong inferences due to known issues such as sparse data bias or winner’s curse inflating effect sizes [178,179]. An alternative can be to establish larger research collaborations that can pool data into larger cohorts as we found several examples of [63,144]. Worth noting, we found two clinical prediction models. In one of them, the IRONIC group developed a scoring system for the risk of multidrug resistance in bloodstream infections by *P. aeruginosa*, and in another, Colombian researchers developed a scoring system for the risk of ESBL-producing *Enterobacteriacaea* [128,144].

Studies were assessed for their risk of bias using the NIH Quality Assessment tool, in which we found several recurring issues. Most studies did not include a sample size calculation or described the blinding of exposure assessors. Differences in the risk of bias assessments were often determined by allowing continuous variables to be treated continuously or by the correct definition of the exposure. We also found that several studies lacked a reliable definition of the outcome, in particular the definition of mortality. As discussed by the NIH Quality Assessment reviewers, even though death as an outcome seems to be objective by nature (i.e. researchers rely on “face validity”), even this outcome should be defined. Such a definition should include information on particulars like from which register or medical record information about the death was collected if the patient perished within or outside of the institution, and if the latter was the case, who reported the death. Most studies did have a sufficient follow-up time to capture infectious disease outcomes (in particular mortality), but these follow-up times varied greatly for all studies. However, the NIH Quality Assessment tool included items that were not relevant to all studies (e.g. “the adjustment of confounders” in a prediction context) and the issue with finding good quality assessment tools for prognostic factor research has been described earlier [180]. Systematic reviews of clinical prediction models should use other more specialised tools like PROBAST [181].

We chose to summarise the research by describing an in-depth example of studies on VRE, which provided us with a representative cross-section in terms of the patient population, sample size, methodological approaches, and the number of variables. This example showed how heterogeneity in how the subject is studied may hamper our ability to build on this basic research, either through the development of more comprehensive predictive models or through pooling or meta-analysis. Most studies studying resistant enterococci group the bacteria together with other bacteria and/or fungi. Furthermore, authors often study the bacteria in a highly selected patient population, in which it is difficult to infer how baseline characteristics pertain to the risk factors studied, either from a table of characteristics or the model. However, the major source of heterogeneity that reduces our ability to build on the research is the arbitrary way in which variables are selected.

We find that throughout the entire material, there was widespread use of either stepwise regression or a bivariable screening of variables as a method for variable selection in regression models, where variables that achieve some pre-specified p-value threshold are included in the final model. This method is, however, not recommended for either estimating the effect of an exposure or predicting an outcome [10,182–184]. In short, the reason is its reliance on p-values as a criterion for variable selection, although the p-value is not an indicator of a causal relationship or predictive power. A factor confounding a causal effect may not be statistically significant, and there may be situations in which a variable that is not statistically significant in a bivariable analysis may increase the predictive power of a multivariable model. Consequently, including all candidate factors in a multivariable model and testing them through statistical significance may not be a valid method of discovering new prognostic factors. Unfortunately, there is no alternative to these methods based on statistical significance that is as simple and practical, or as automated and data-driven. The basis for all modelling is a theoretical understanding of covariates, outcomes and the relationship between them. All in all, it is difficult and maybe even impossible to summarise what risk factors are shown to be of relevance in this literature. Simply counting how many times a certain risk factor is found to be statistically significant tells us little of its relative importance.

The strengths of our study are the large scope of our searches, which has resulted in a comprehensive overview of the current state of this research topic. However, there are several limitations to our study. First, we only searched for studies where any type of antimicrobial resistance was mentioned in the title, abstract, controlled vocabulary, or keywords, which may have excluded some studies that only mention infections in broader terms, but still model the risk of contracting resistant infections or any potential outcomes of such infections. Furthermore, we narrowed our inclusion criteria to studies that model either infection/colonisation or mortality as an outcome, but this does not fully cover how resistant microbes may increase the disease burden among cancer patients, like repeated hospitalisations, increased costs and/or increased length of stays. Several of the studies that were excluded mainly had patients with a haematological malignancy, but also some patients with aplastic anaemia or other haematological disorders. Other studies did not explicitly include (acquired or intrinsic) resistance in the models, although they may have included resistance in other implicit ways, like the failure of an empiric carbapenem cure. Patients with other haematological disorders or other infectious etiologies may have similar risk profiles, and their exclusion may have unreasonably narrowed the scope of the review. We also searched for records in Swedish, Danish and Spanish, but records in these languages were not included to ensure this systematic review would be available and reproducible for all readers. As many of the studies we included were from countries where English is not the native language, the exclusion of other languages may represent a limitation of our review.

### Conclusions

In this systematic review, we found a great level of heterogeneity in the approach to studying risk factors for resistant infections/colonisations and mortality due to resistant infections among cancer patients. We argue that it’s difficult or even impossible to use these models to infer which risk factors are of particular importance. This is due to differences in the patient populations selected, and the different ways of grouping microbes. Furthermore, studies on this subject often have a small sample size and use p-value-based variable selection methods, which lead to very diverse models. The consequence of this heterogeneity is not only that the literature is unprepared for either a meta-analysis or a pooled analysis. It also means that it is difficult to use this research to understand the mechanisms by which resistant microbes cause disease in cancer patients, and thus that it is difficult for clinicians to use the research to guide their prevention of such conditions. This represents a serious shortcoming of this body of literature. There is a need to develop and adhere to more standardised protocols when investigating new risk factors. Such protocols should include a clear aim of what risk factors are to be explored and build on existing literature, e.g. by selecting similar patient populations and being careful to include factors previously shown to be of importance regardless of their p-value in a bivariable screening.

## Data Availability

All data extracted or generated and then analysed are included in the article or the supplementary material.

## Declarations

### Acknowledgements

We would like to thank bachelor’s student Sofie Almark Jeppesen for assistance in the screening process. We would also like to thank the authors who shared their research with us to which we did not otherwise have access. Thanks also to the South-Eastern Norway Regional Health Authority for funding.

### Ethics approval and consent to participate

Not applicable

### Consent for publication

Not applicable

### Authors’ contributions

AD conceived the idea, drafted the manuscript, screened the search, extracted and organised the data, and performed the risk of bias assessment. LF revised the manuscript, screened the search, extracted and organised the data, analysed the data, and performed the risk of bias assessment. SP revised the manuscript. AM revised the manuscript. RT revised the manuscript and performed the search. OK revised the manuscript, and performed the risk of bias assessment. JB conceived the idea and revised the manuscript.

## Supplementary material 1

### First search

**Database: Ovid MEDLINE(R) and Epub Ahead of Print, In-Process, In-Data-Review & Other Non-Indexed Citations, Daily and Versions(R) <1946 to June 21, 2021>**

**Date:** 22.06.21

**Hits:** 6878

|  |  |  |
| --- | --- | --- |
| 1 | exp Neoplasms/ | 3485612 |
| 2 | (tumor? or tumour? or cancer? or neoplas* or paraneoplastic or precancer* or cyst? or malignanc* or carcinogenes#s or oncogenes#s or tumorigenes#s or cocarcinogenes#s or metastas#s or metastatic or incidentaloma?).tw,kf. | 3441687 |
| 3 | 1 or 2 | 4533774 |
| 4 | exp Drug Resistance, Microbial/ or exp Drug Resistance, Bacterial/ | 170727 |
| 5 | ((antibacterial or bacterial or bacterium or antibiotic or antimicrobial or microbial or multidrug or multiple drug) adj2 (resistance or resistant)) or antibiotic non-susceptibility or antibiotic nonsusceptibility).tw,kf. | 136901 |
| 6 | 4 or 5 | 254631 |
| 7 | 3 and 6 | 22036 |
| 8 | limit 7 to (danish or english or interlingua or multilingual or norwegian or spanish or swedish) | 20721 |
| 9 | limit 8 to yr="2015 -Current" | 6878 |

**Database: Embase <1974 to 2021 June 22>**

**Date:** 23.06.21

**Hits:** 7314

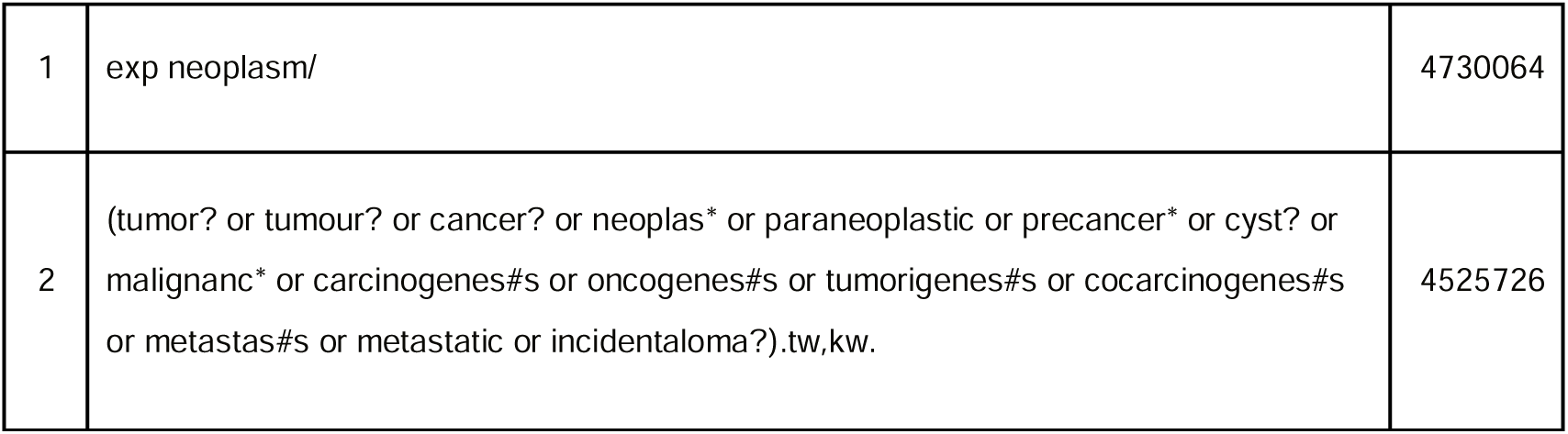
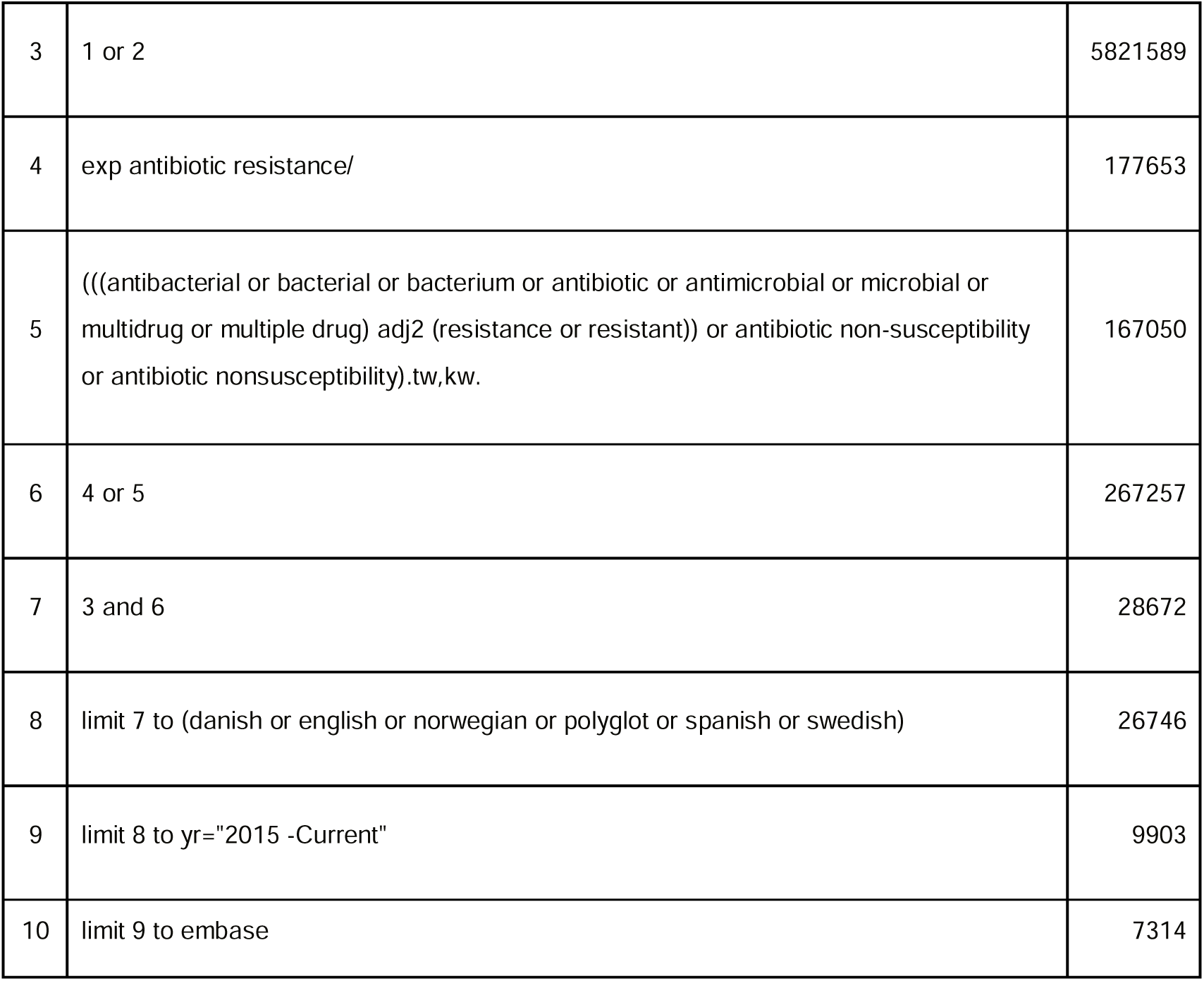

|  |  |  |
| --- | --- | --- |
| 1 | exp neoplasm/ | 4730064 |
| 2 | (tumor? or tumour? or cancer? or neoplas* or paraneoplastic or precancer* or cyst? or malignanc* or carcinogenes#s or oncogenes#s or tumorigenes#s or cocarcinogenes#s or metastas#s or metastatic or incidentaloma?).tw,kw. | 4525726 |
| 3 | 1 or 2 | 5821589 |
| 4 | exp antibiotic resistance/ | 177653 |
| 5 | ((antibacterial or bacterial or bacterium or antibiotic or antimicrobial or microbial or multidrug or multiple drug) adj2 (resistance or resistant)) or antibiotic non-susceptibility or antibiotic nonsusceptibility).tw,kw. | 167050 |
| 6 | 4 or 5 | 267257 |
| 7 | 3 and 6 | 28672 |
| 8 | limit 7 to (danish or english or norwegian or polyglot or spanish or swedish) | 26746 |
| 9 | limit 8 to yr="2015 -Current" | 9903 |
| 10 | limit 9 to embase | 7314 |

**Database: Web of Science (Science Citation Index Expanded (SCI-EXPANDED) --1987-present, Social Sciences Citation Index (SSCI) --1987-present, Arts & Humanities Citation Index (A&HCI) -- 1987-present, Emerging Sources Citation Index (ESCI) --2015-present)**

**Date:** 24.06.21

**Hits:** 11,302

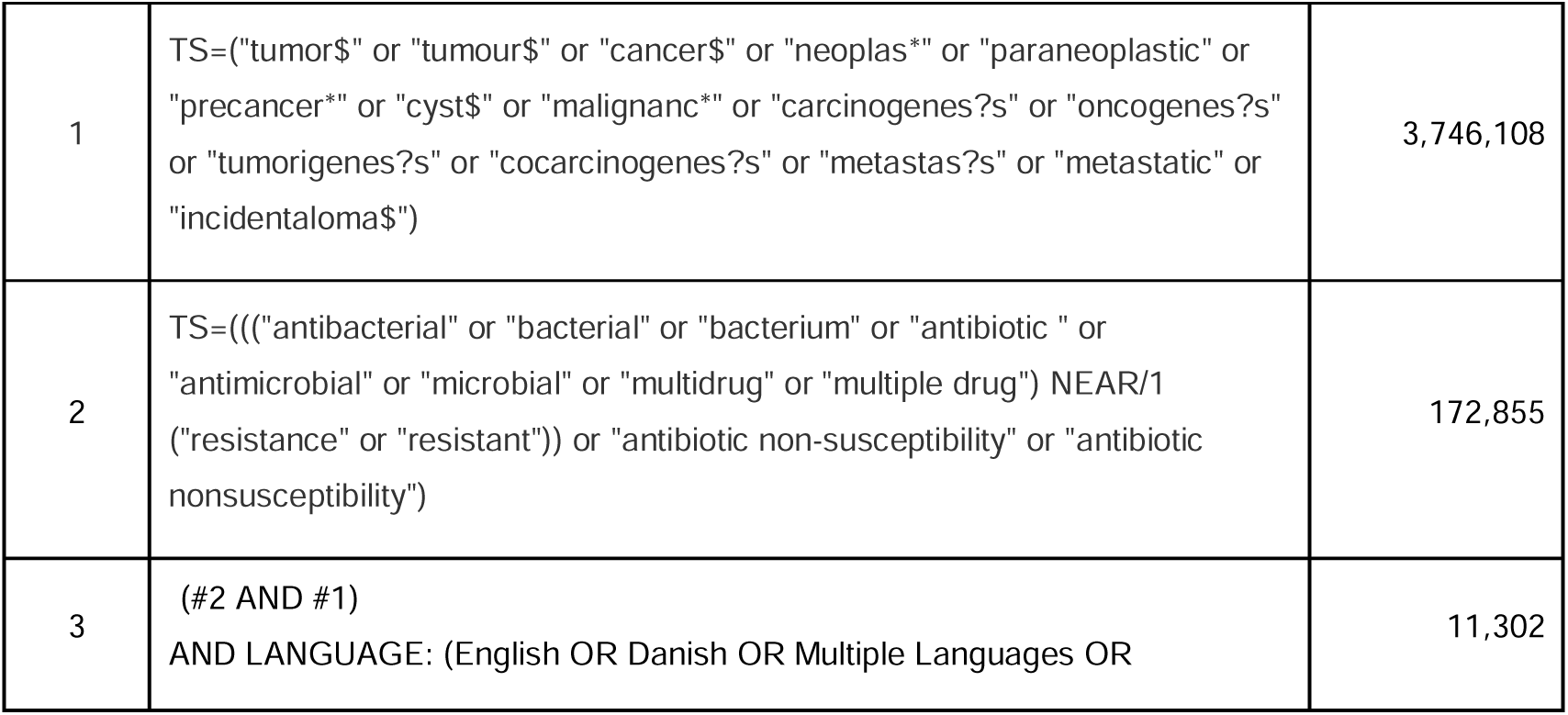
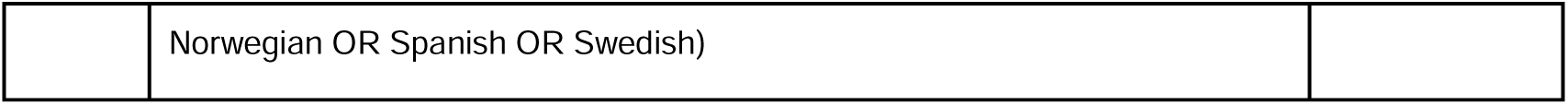

|  |  |  |
| --- | --- | --- |
| 1 | TS=("tumor\$" or "tumour\$" or "cancer\$" or "neoplas*" or "paraneoplastic" or "precancer*" or "cyst\$" or "malignanc*" or "carcinogenes?s" or "oncogenes?s" or "tumorigenes?s" or "cocarcinogenes?s" or "metastas?s" or "metastatic" or "incidentaloma\$") | 3,746,108 |
| 2 | TS=(((antibacterial" or "bacterial" or "bacterium" or "antibiotic " or "antimicrobial" or "microbial" or "multidrug" or "multiple drug") NEAR/1 ("resistance" or "resistant")) or "antibiotic non-susceptibility" or "antibiotic nonsusceptibility") | 172,855 |
| 3 | (#2 AND #1)<br>AND LANGUAGE: (English OR Danish OR Multiple Languages OR | 11,302 |

|  |  |
| --- | --- |
|  | Norwegian OR Spanish OR Swedish) |

## Supplementary material 2

### Second search

**Database: Ovid MEDLINE(R) and Epub Ahead of Print, In-Process, In-Data-Review & Other Non-Indexed Citations, Daily and Versions(R) <1946 to November 18, 2021>**

**Date:** 19.11.21

**Hits:** 12 478

|  |  |  |
| --- | --- | --- |
| 1 | exp Neoplasms/ | 3573100 |
| 2 | (tumor? or tumour? or cancer* or neoplas* or paraneoplas* or preneoplas* or precancer* or premalign* or malign* or carcinogenes#s or oncogenes#s or tumorigenes#s or cocarcinogenes#s or metastas#s or metastatic or incidentaloma? or leukemia? or leucemia? or lymphoma? or sarcoma? or carcinoma* or melanoma? or glioma ? or blastoma?).tw,kf. | 3958905 |
| 3 | 1 or 2 | 4744972 |
| 4 | exp Drug Resistance, Microbial/ or exp Drug Resistance, Bacterial/ | 174771 |
| 5 | ((antibacterial or bacterial or bacterium or antibiotic or antifungal or antimicrobial or microbial or multidrug or multiple drug or multiple antibiotics or penicillin or meth#cillin? or met#cillin? or cephalosporin* or cephalosporanic acid? or cefepim? or cefuroxime or cefotaxim? or ceftazidime or ceftriaxon? or cefixime or ceftaroline or carbapenem? or aminoglycoside? or gentam#cin? or amikacin? or fluoroquinolone? or linezolid? or vancomycin? or echinocandin? or azole? or colistin? or colimycin? or polymyxin e or intrinsic* or penicillin or macrolide or tobramycin? or fluconazol? or voriconazole or posaconazole or caspofungin or anidulafungin or micafungin or ciproflox* or ciproxin or levofloxacin or clindamycin? or dalacin or clarithromycin or CLAR or erythromycin or azt?reonam or azactam) adj4 (resistance or resistant or nonsusceptib* or insusceptib* or non-susceptib* or susceptibility)).tw,kf. | 235262 |
| 6 | (piperacillin adj3 tazobactam adj4 resistant).tw,kf. | 202 |
| 7 | beta-Lactamases/ or Methicillin-Resistant Staphylococcus aureus/ or Vancomycin-Resistant Enterococci/ or Vancomycin-Resistant Staphylococcus aureus/ | 40508 |
| 8 | (b*-lactamase? or "ec 3.5.2.6" or beta lactam?hydrolase or cefinase or ESBL or carbapenemase? or biofilm or non-fermenting).tw,kf. | 88504 |
| 9 | Pseudomonas aeruginosa/ or (pseudomonas aeruginosa or "P. aeruginosa").tw,kf. | 74856 |
| 10 | Acinetobacter/ or (acinetobacter* or "A. baumannii").tw,kf. | 20486 |
| 11 | Stenotrophomonas Maltophilia/ or ((Stenotrophomonas or Pseudomonas or Xanthomonas) adj malt#philia).tw,kf. | 3477 |
| 12 | Clostridioides difficile/ or ((clostridium or clostridioides) adj difficil*).tw,kf. | 17332 |
| 13 | Enterococcus faecium/ or ((enterococcus or streptococcus) adj faecium).tw,kf. | 7433 |
| 14 | (candida adj1 (non albicans or auris)).tw,kf. | 1779 |
| 15 | Aspergillus fumigatus/ or ((aspergillus or neosartorya) adj fumigat*).tw,kf. | 12302 |
| 16 | or/4-15 | 456787 |
| 17 | 3 and 16 | 35096 |
| 18 | limit 17 to (danish or english or interlingua or multilingual or norwegian or spanish or swedish) | 32942 |
| 19 | limit 18 to yr="2015 -Current" | 12478 |

**Database: Embase <1974 to 2021 November 17>**

**Date:** 19.11.21

**Hits:** 16471

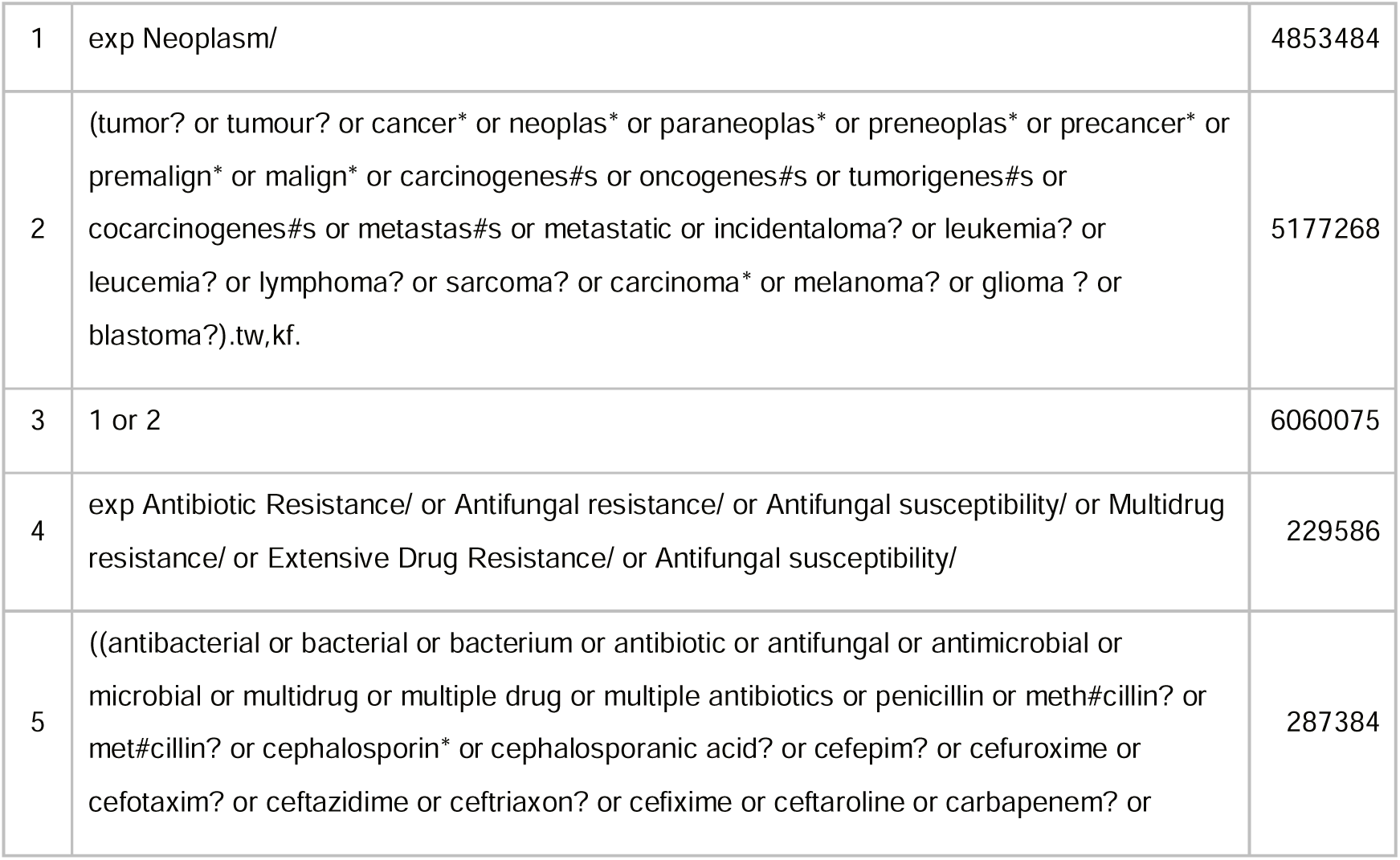
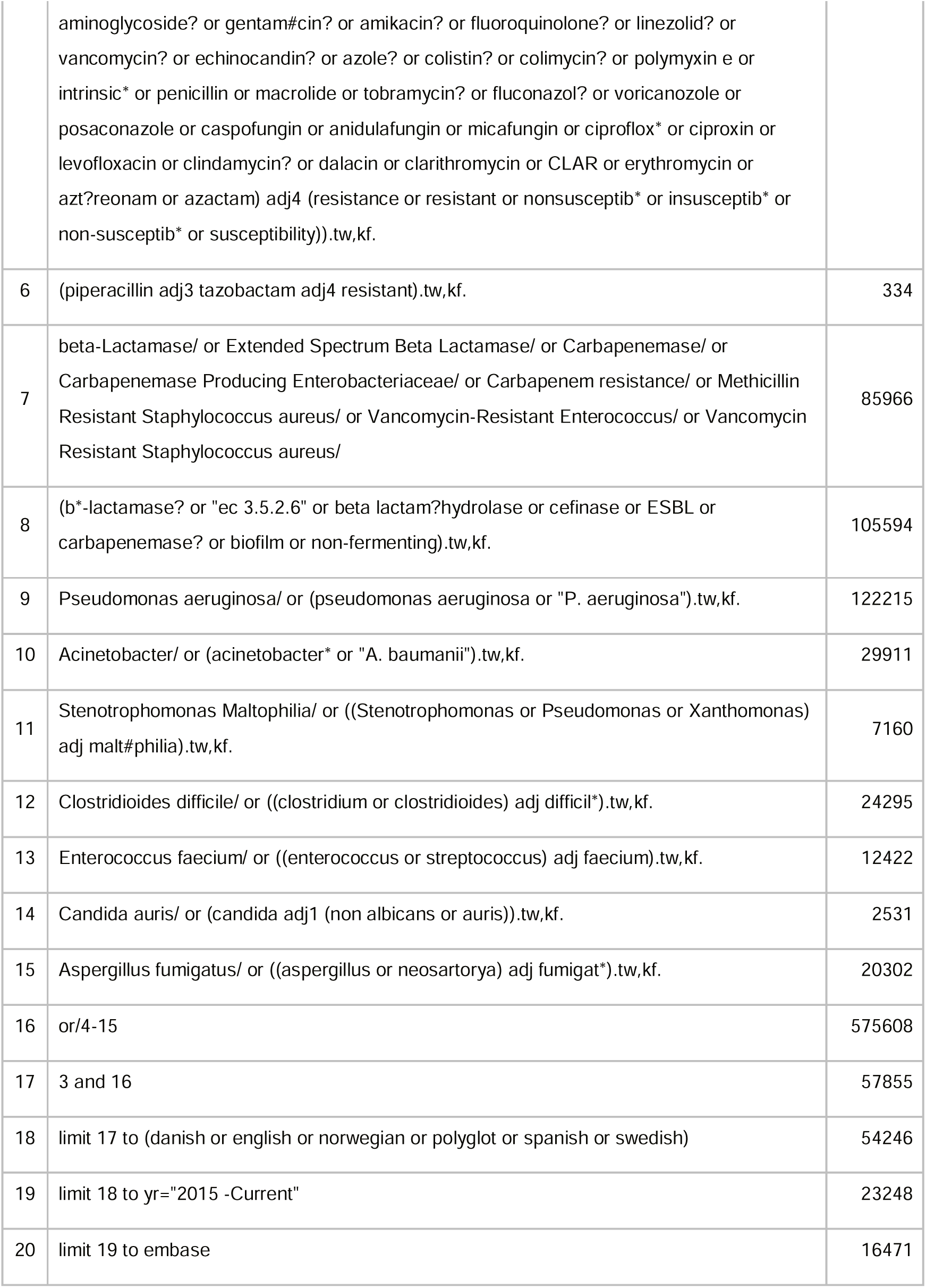

**Database: Web of Science Core Collection: Science Citation Index Expanded (SCI-EXPANDED) - -1987-present, Social Sciences Citation Index (SSCI) --1987-present, Arts & Humanities Citation Index (A&HCI) --1987-present, Emerging Sources Citation Index (ESCI) --2015-present**

**Date:** 19.11.21

**Hits:** 19 369

|  |  |  |
| --- | --- | --- |
| 14 | (#13) AND LA=(English OR Danish OR Multiple Languages OR Norwegian OR Spanish OR Swedish) | 19,369 |
| 13 | (#12) AND PY=(2015-2021) | 19,528 |
| 12 | #1 and #11 | 46,063 |
| 11 | #2 or #3 or #4 or #5 or #6 or #7 or #8 or #9 or #10 | 454,205 |
| 10 | TS=((("aspergillus" or "neostartorya") NEAR/0 "fumigat*")) | 13,687 |
| 9 | TS=("candida" NEAR/0 ("non albicans" or "auris")) | 1,906 |
| 8 | TS=((("enterococcus" or "streptococcus") NEAR/0 "faecium")) | 8,677 |
| 7 | TS=((("clostridium" or "clostridioides") NEAR/0 "difficil*")) | 20,940 |
| 6 | TS=((("Stenotrophomonas" or "Pseudomonas" or "Xanthomonas") NEAR/0 "malt?philia")) | 4,062 |
| 5 | TS=("acinetobacter*" or "A. baumannii") | 23,475 |
| 4 | TS=("pseudomonas aeruginosa" or "P. aeruginosa") | 88,969 |
| 3 | TS=("b*-lactamase\$" or "ec 3.5.2.6" or "beta lactam hydrolase" or "beta lactamhydrolase" or "cefinae" or "ESBL" or "carbapenemase\$" or "biofilm" or "non-fermenting") | 118,459 |
| 2 | TS=((("antibacterial" or "bacterial" or "bacterium" or "antibiotic\$" or "antifungal" or "antimicrobial" or "microbial" or "multidrug" or "multiple drug" or "multiple antibiotics" or "penicillin" or "meth?cillin\$" or "met?cillin\$" or "cephalosporin*" or "cephalosporanic acid\$" or "cefepim\$" or "cefuroxime" or "cefotaxim\$" or "ceftazidime" or "ceftriaxon\$" or "cefixime" or "ceftaroline" or "carbapenem\$" or "aminoglycoside\$" or "gentam?cin\$" or "amikacin\$" or "fluoroquinolone\$" or "linezolid\$" or "vancomycin\$" or "echinocandin\$" or "azole\$" or "colistin\$" or "colimycin\$" or "antibacterial" or "bacterial" or "bacterium" or "antibiotic" or "antifungal" or "antimicrobial" or "microbial" or "multidrug" or "multiple drug" or "multiple antibiotics" or "penicillin" or "meth?cillin\$" or "met?cillin\$" or "cephalosporin*" or "cephalosporanic acid\$" or "cefepim\$" or "cefuroxime" or "cefotaxim\$" or "ceftazidime" or "ceftriaxon\$" or "cefixime" or "ceftaroline" or "carbapenem\$" or "aminoglycoside\$" or "gentam?cin\$" or "amikacin\$" or "fluoroquinolone\$" or "linezolid\$" or "vancomycin\$" or "echinocandin\$" or "azole\$" or | 274,926 |
| | "colistin\$" or "colimycin\$" or "polymyxin e" or "intrinsic*" or "penicillin" or "macrolide" or "tobramycin\$" or "fluconazol\$" or "voriconazole" or "posaconazole" or "caspofungin" or "anidulafungin" or "micafungin" or "ciproflox*" or "ciproxin" or "levofloxacin" or "clindamycin\$" or "dalacin" or "clarithromycin" or "CLAR" or "erythromycin" or "aztreonam" or "azactam" or "polymyxin e" or "intrinsic*" or "penicillin" or "macrolide" or "tobramycin\$" or "fluconazol\$" or "voriconazole" or "posaconazole" or "caspofungin" or "anidulafungin" or "micafungin" or "ciproflox*" or "ciproxin" or "levofloxacin" or "clindamycin\$" or "dalacin" or "clarithromycin" or "CLAR" or "erythromycin" or "aztreonam" or "azactam") NEAR/3 ("resistance" or "resistant" or "nonsusceptible*" or "insusceptible*" or "non-susceptible*" or "susceptibility")) | |
| 1 | TS=("tumor\$" or "tumour\$" or "cancer\$" or "neoplas*" or "paraneoplas*" or "preneoplas*" or "precancer*" or "prealign*" or "malign*" or "carcinogenes?s" or "oncogenes?s" or "tumorigenes?s" or "cocarcinogenes?s" or "metastas?s" or "metastatic" or "incidentaloma\$" or "leukemia\$" or "leucemia\$" or "lymphoma\$" or "sarcoma\$" or "carcinoma*" or "melanoma\$" or "glioma\$" or "blastoma\$") | 4,410,329 |

## Supplementary material 3

**Table S3.**
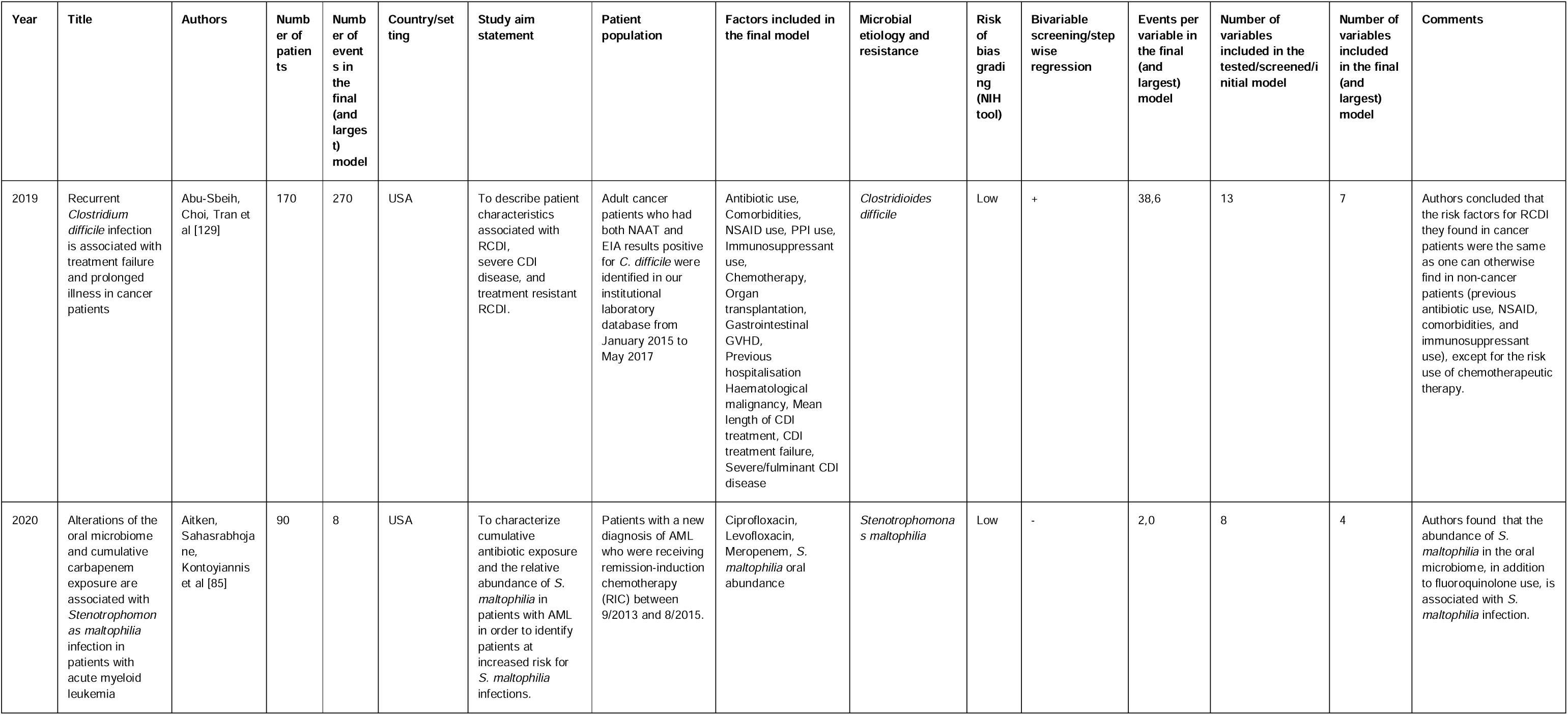
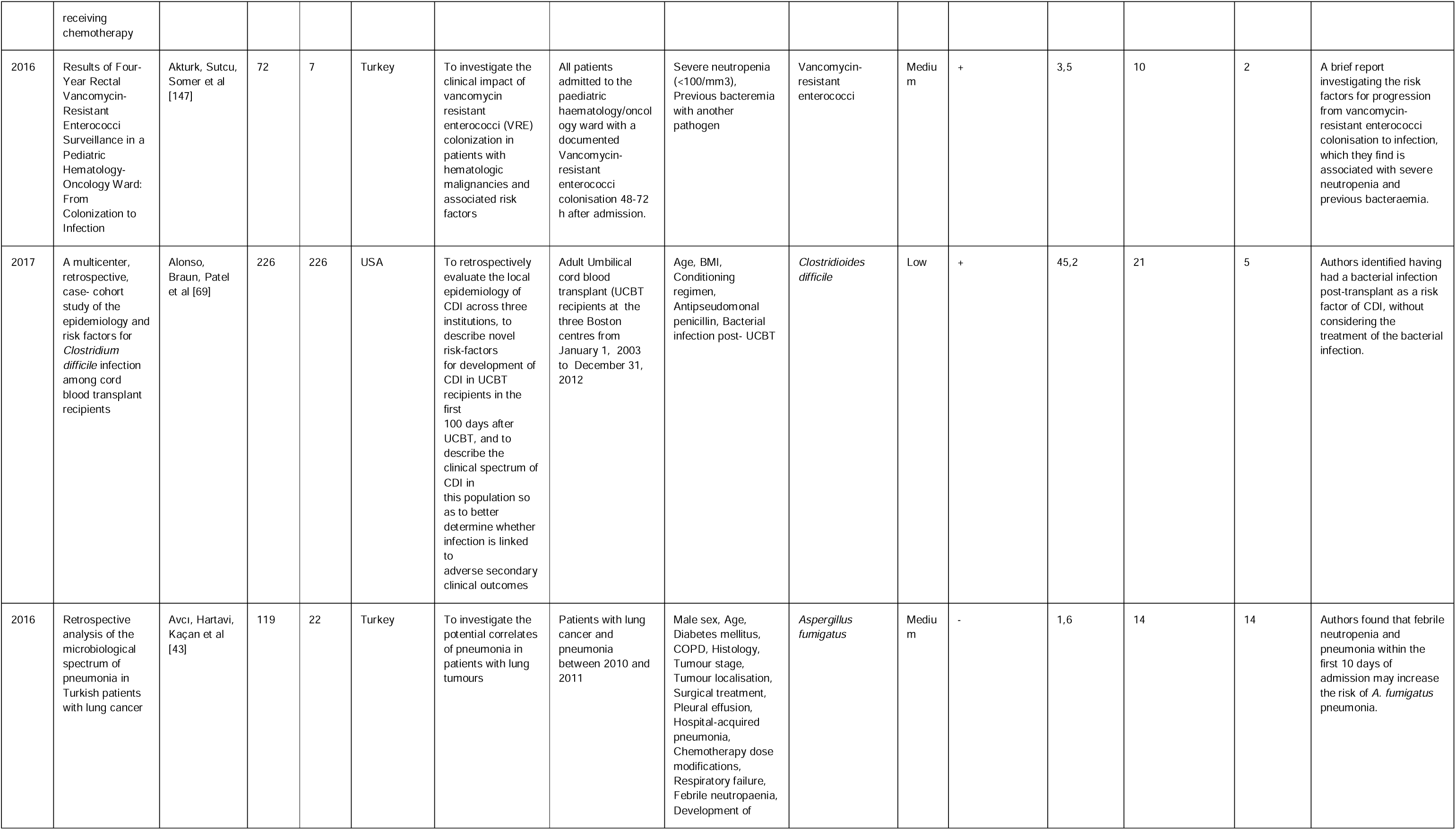
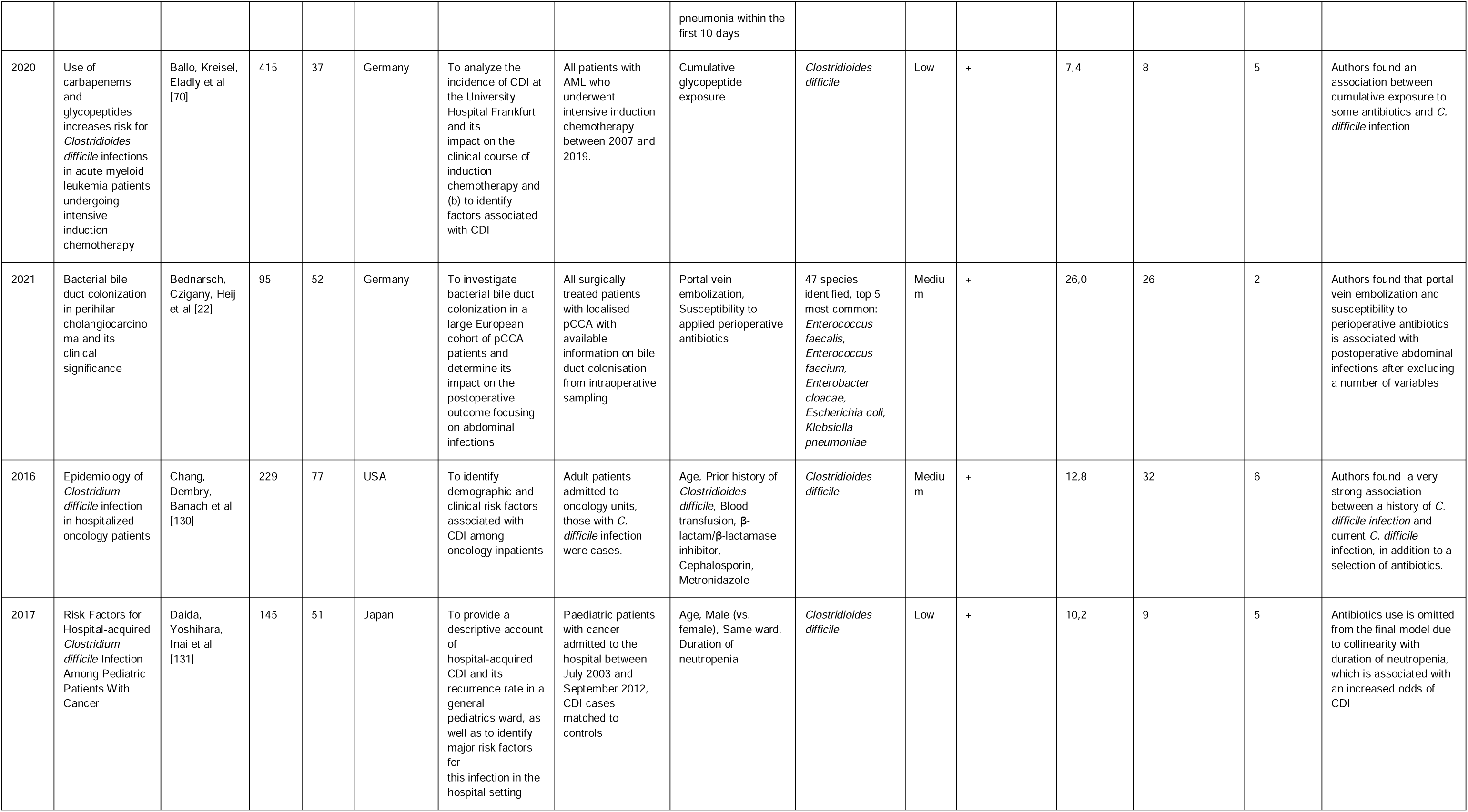
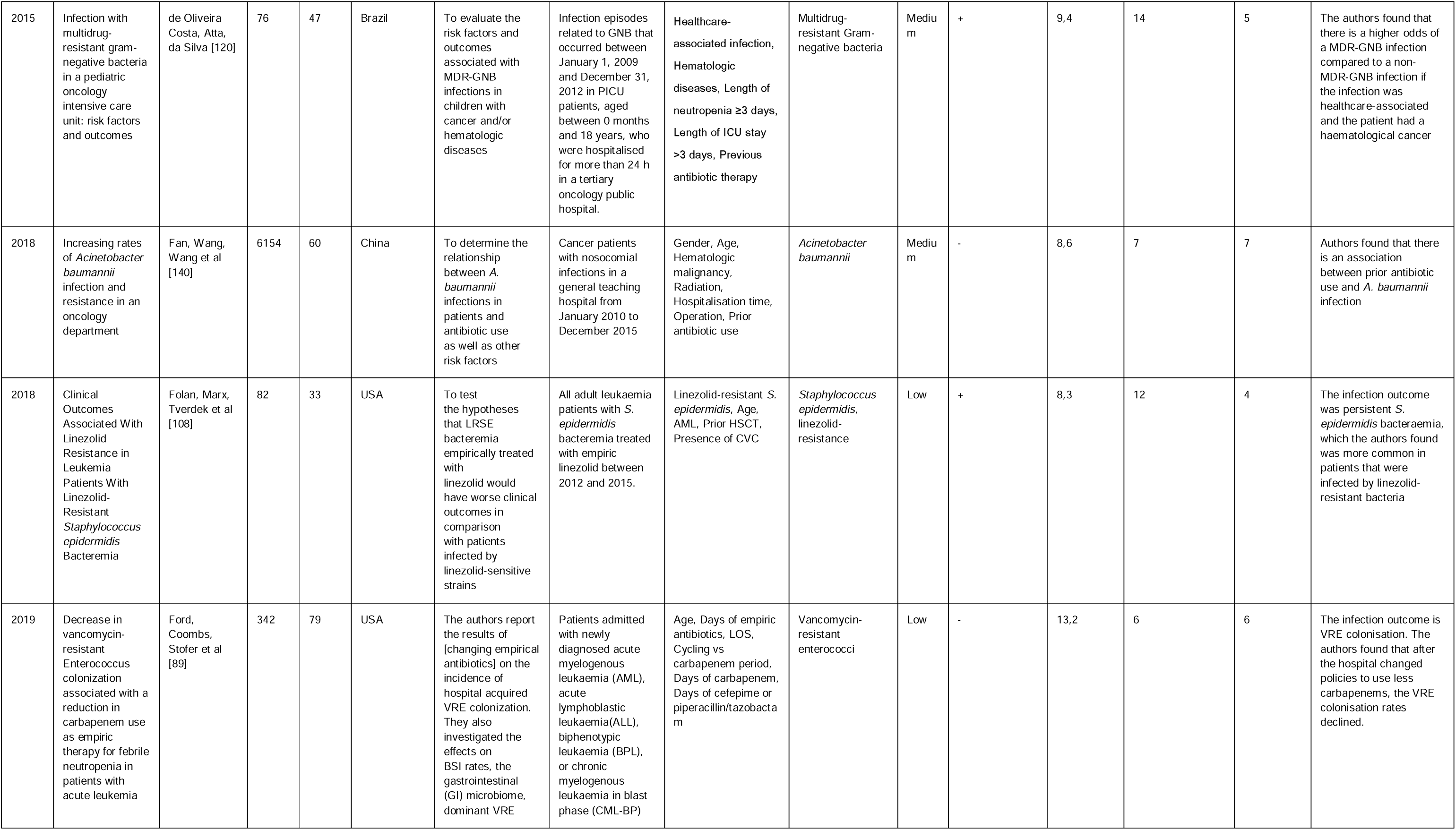
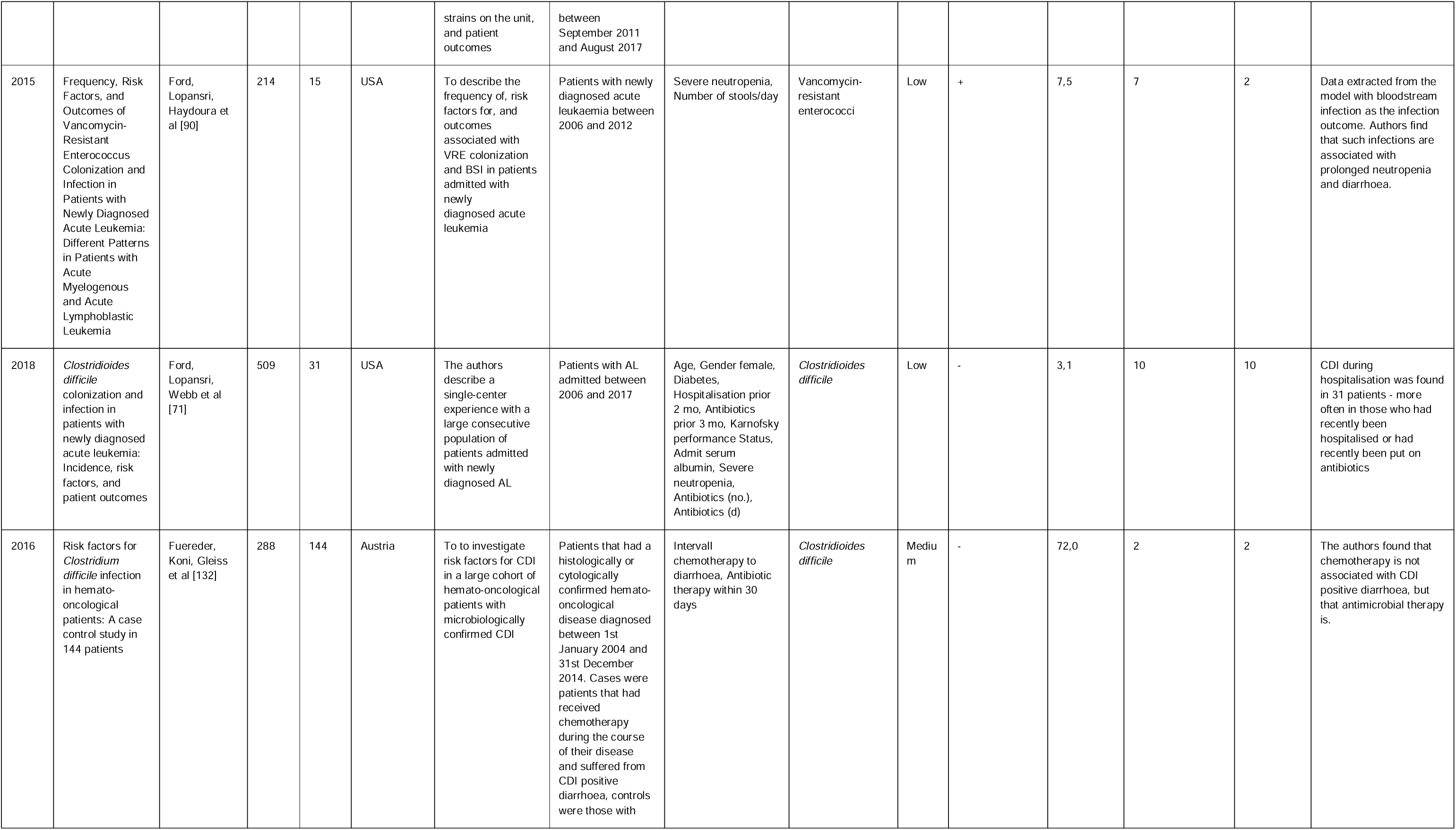
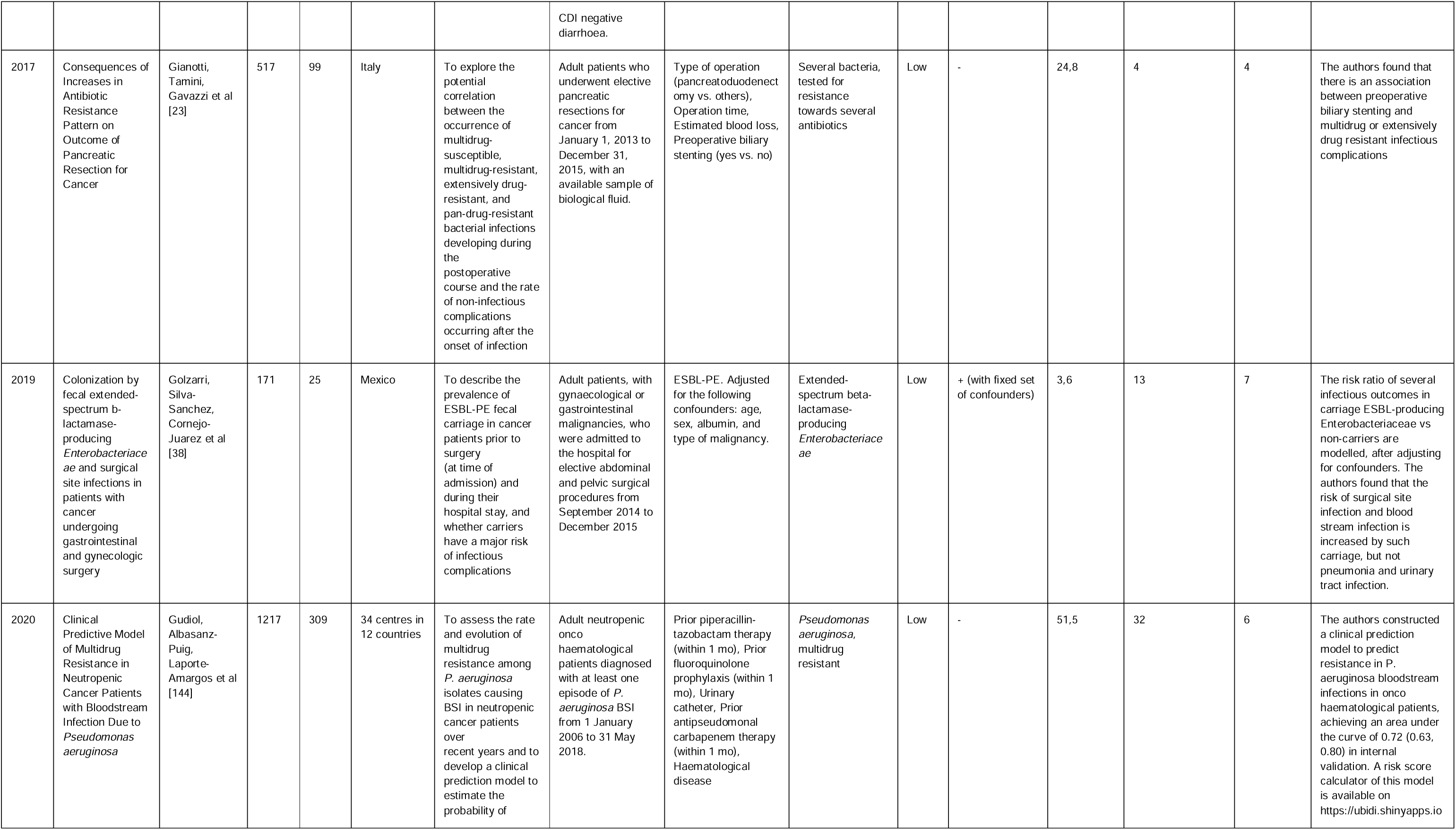
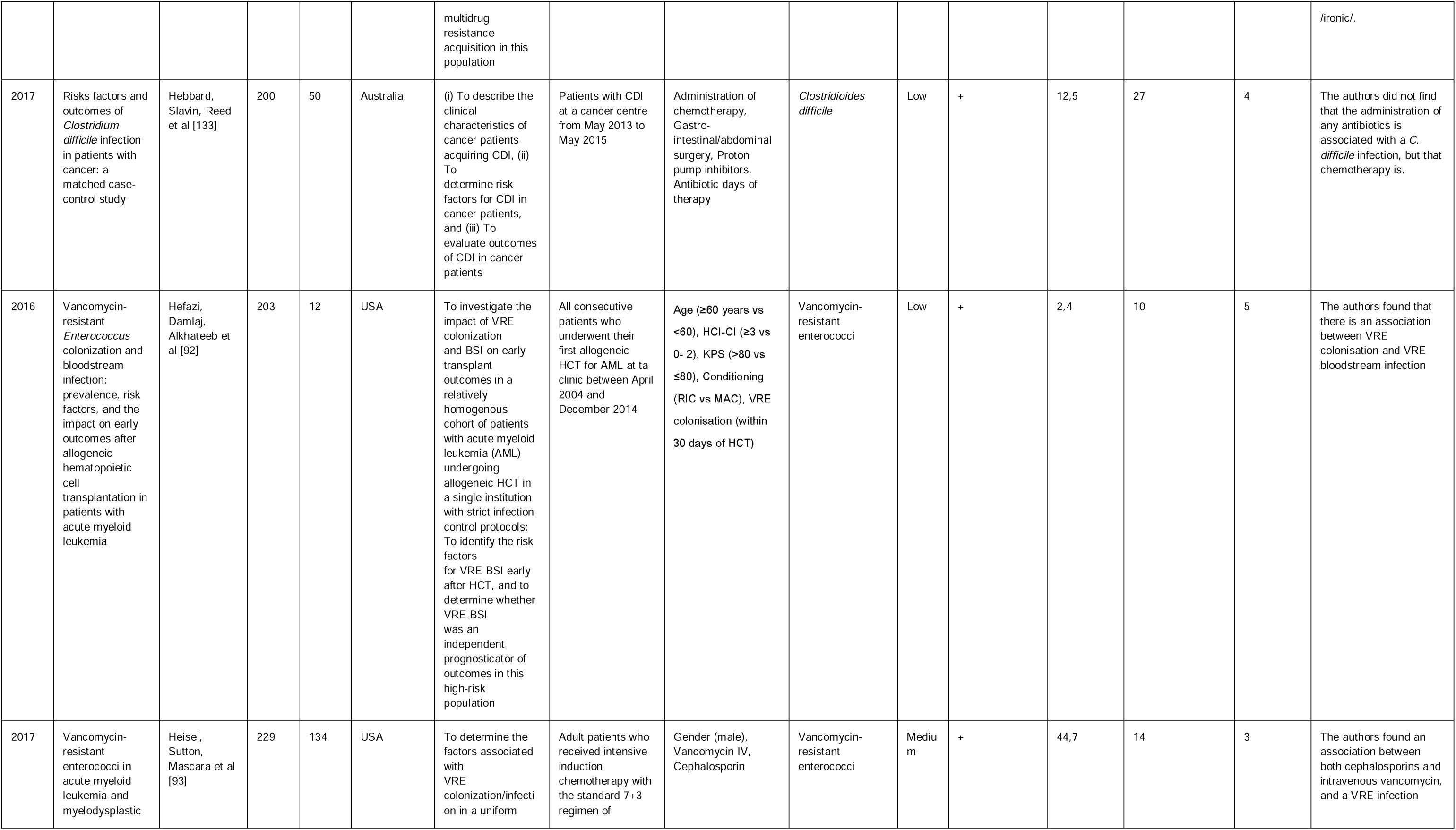
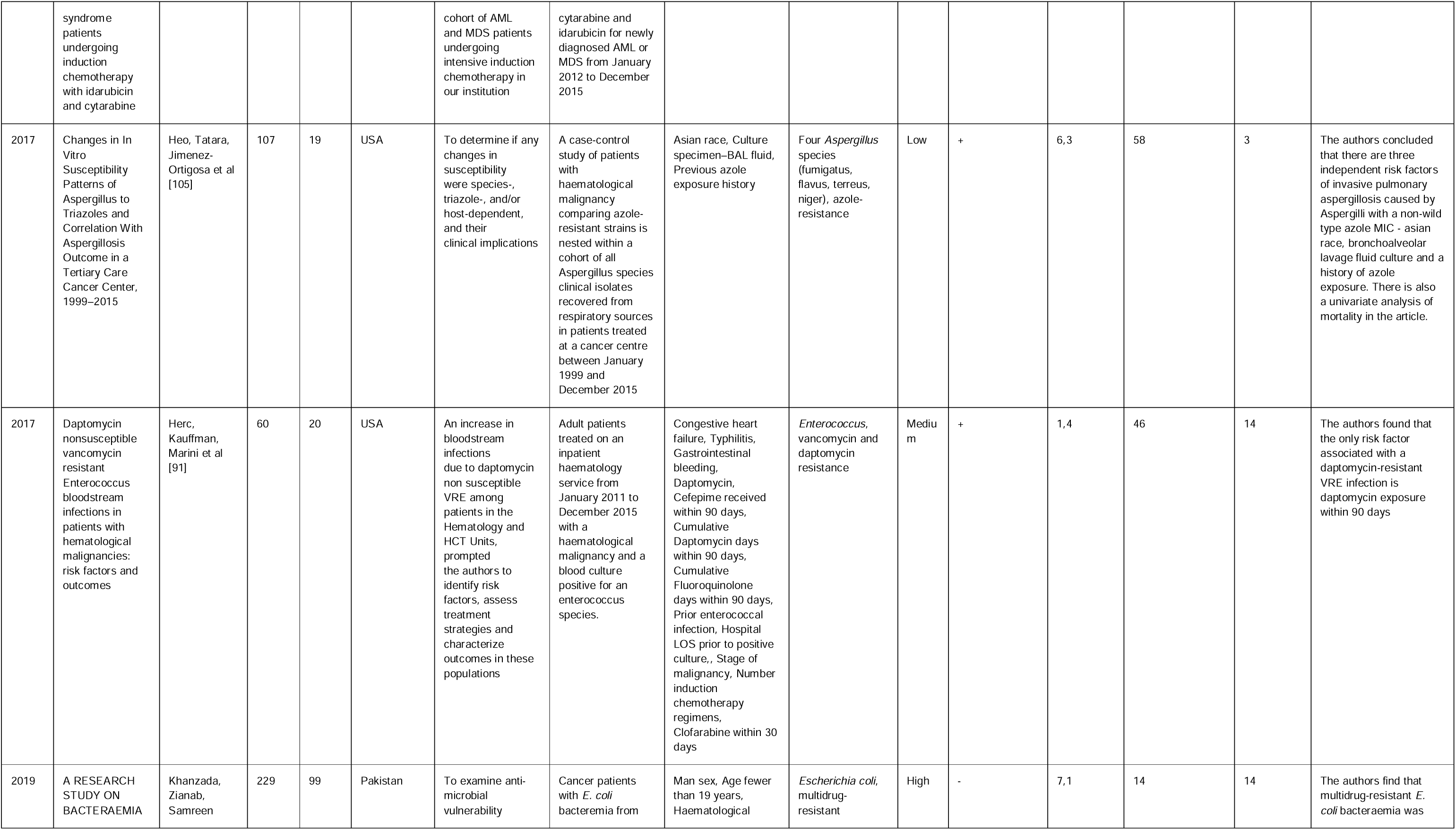
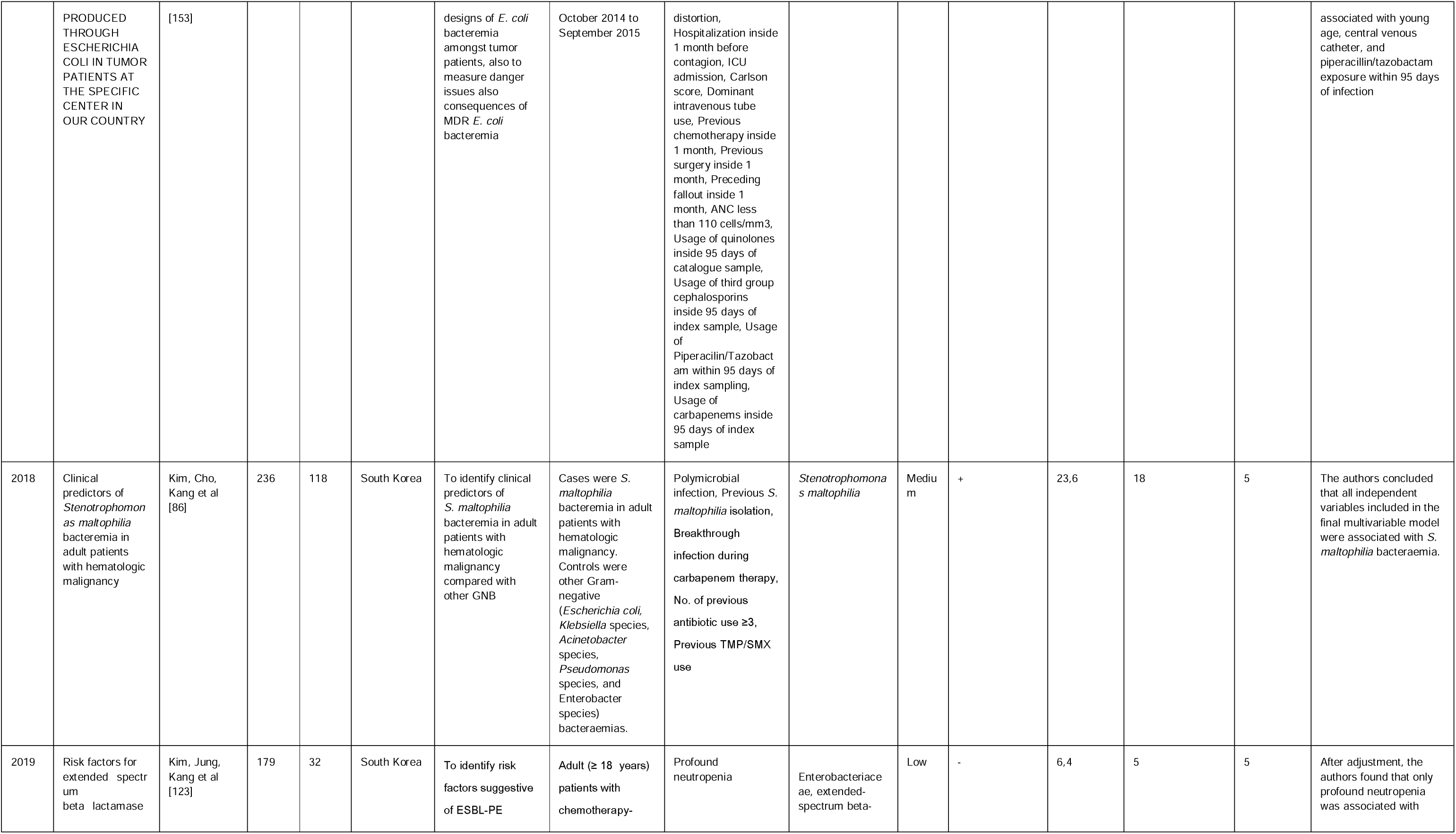
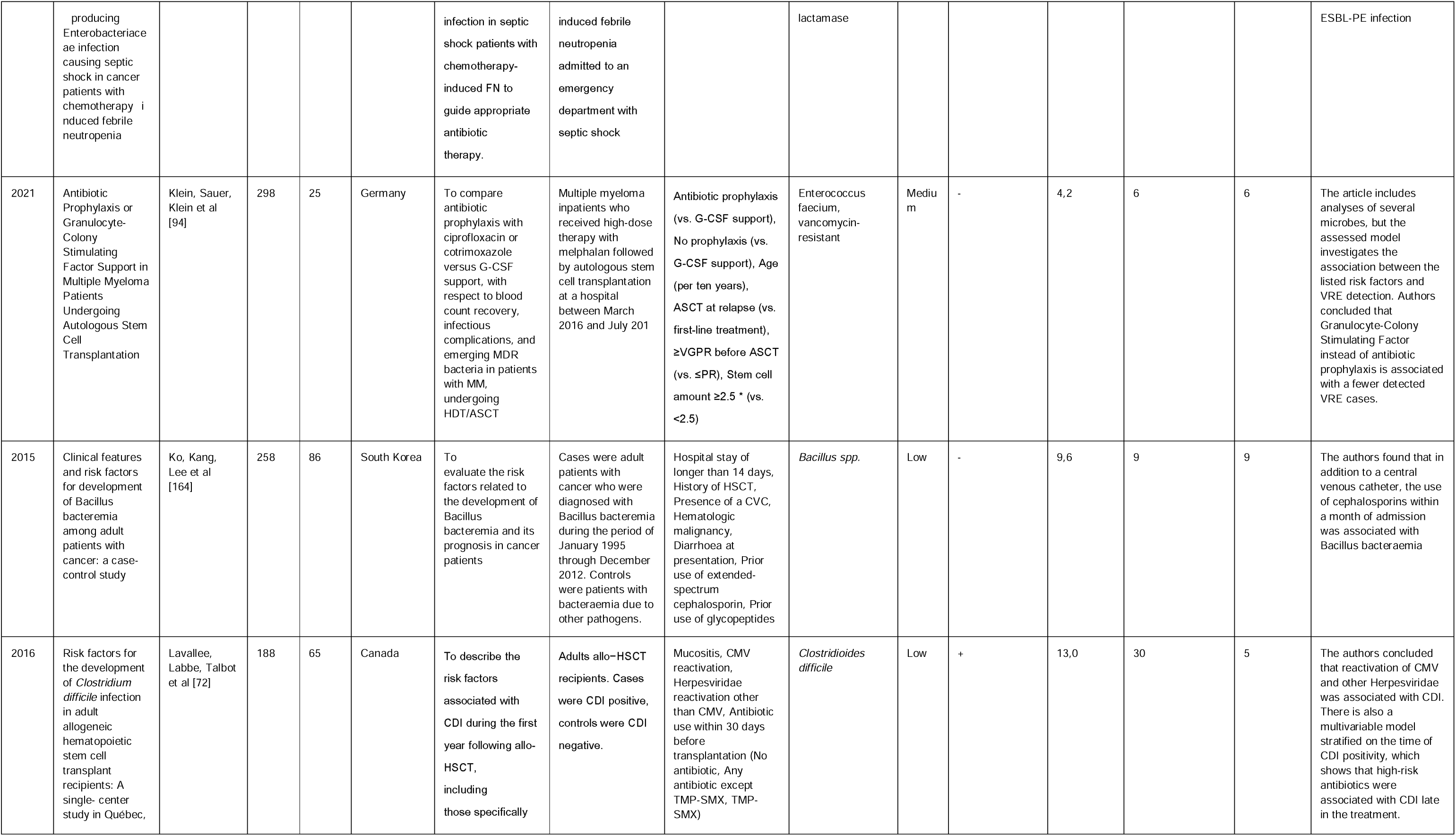
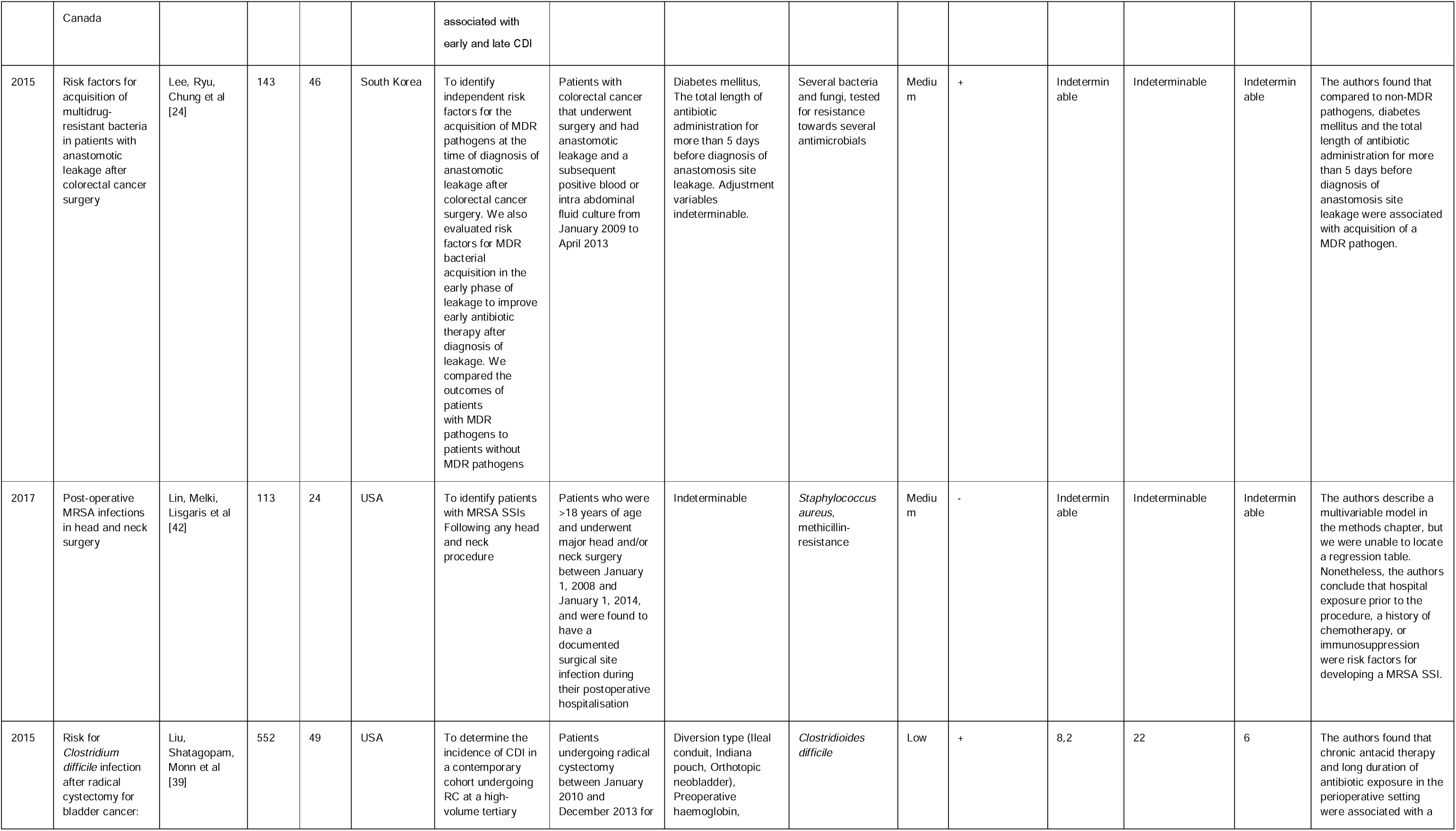
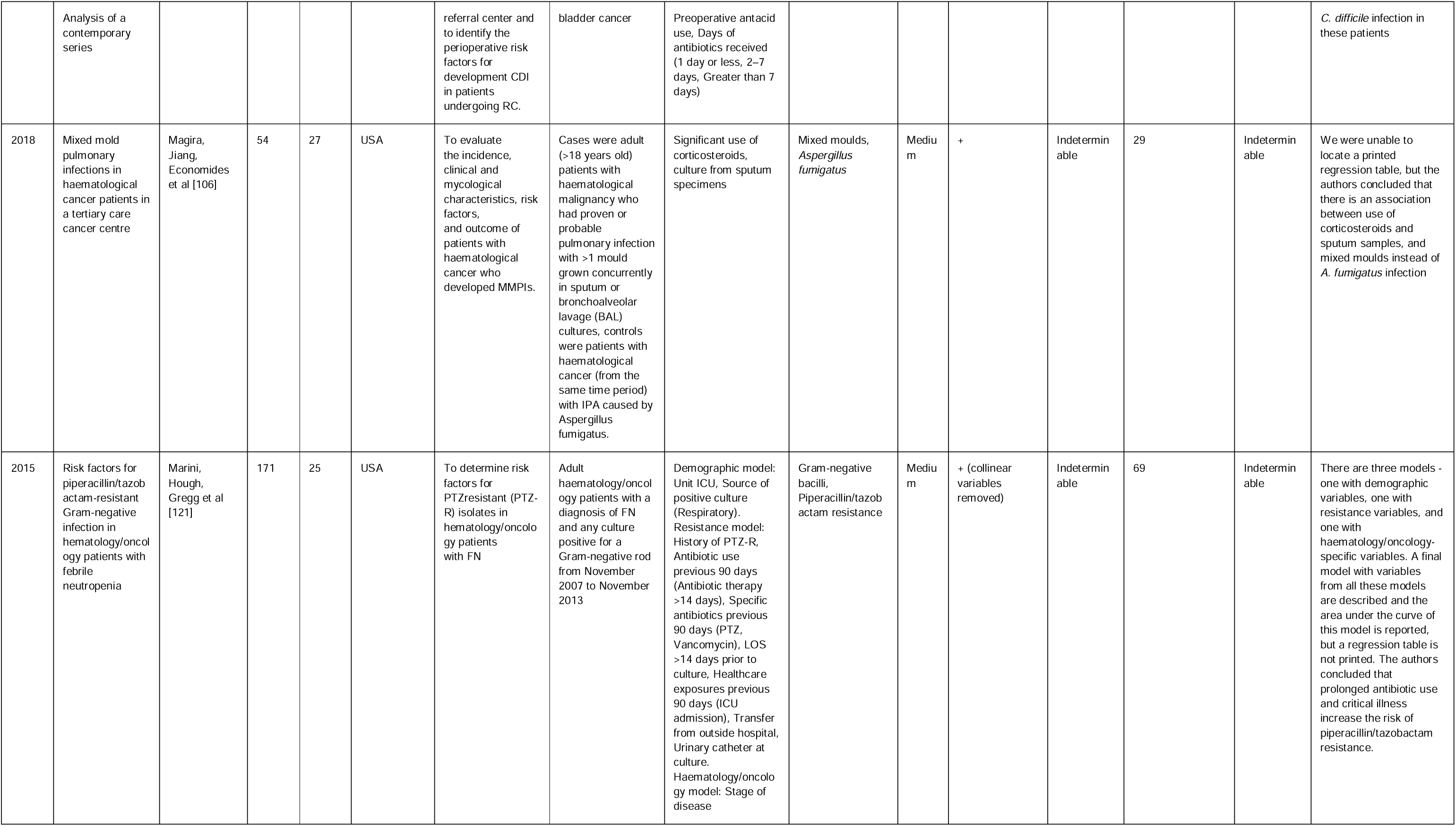
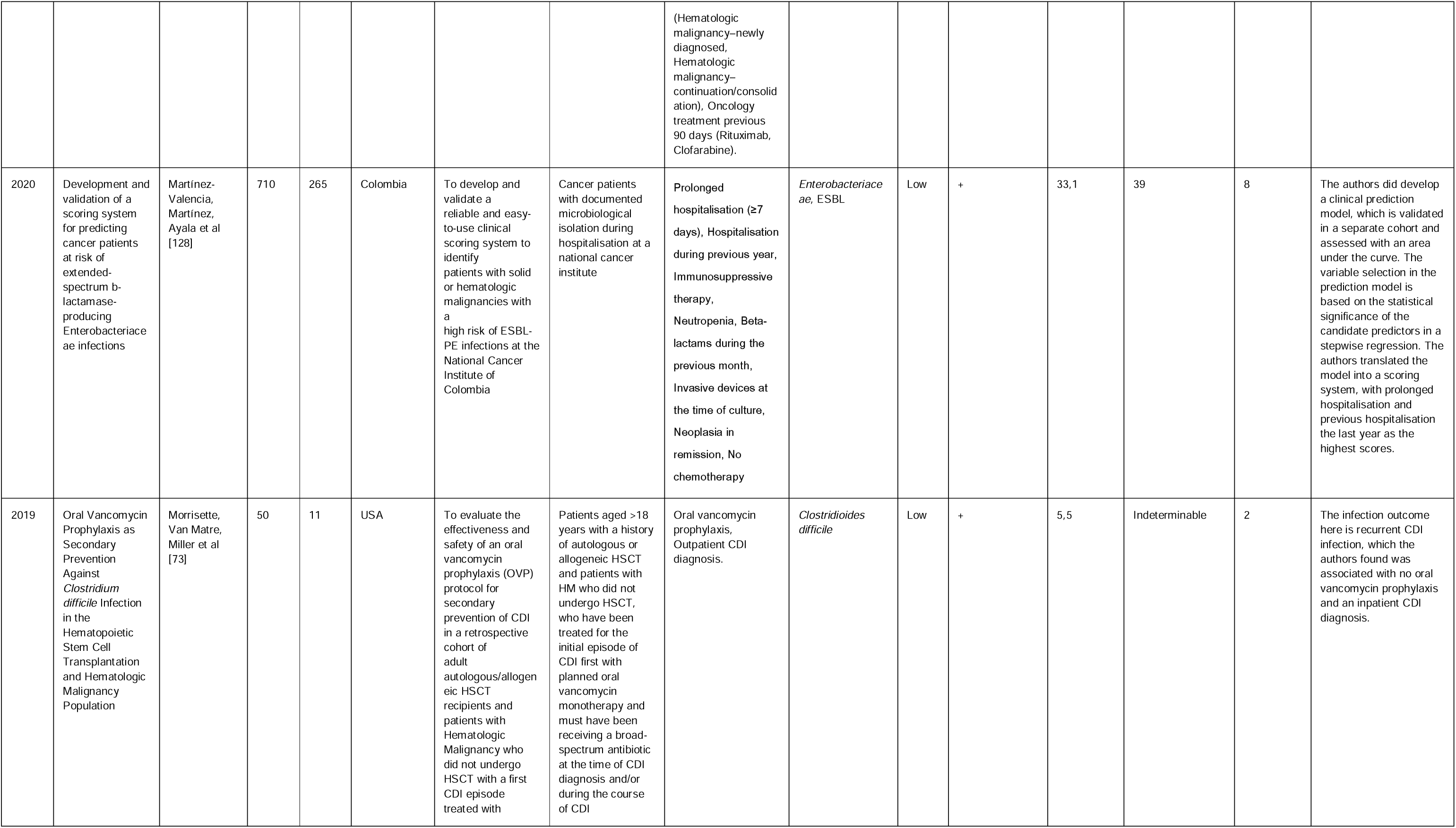
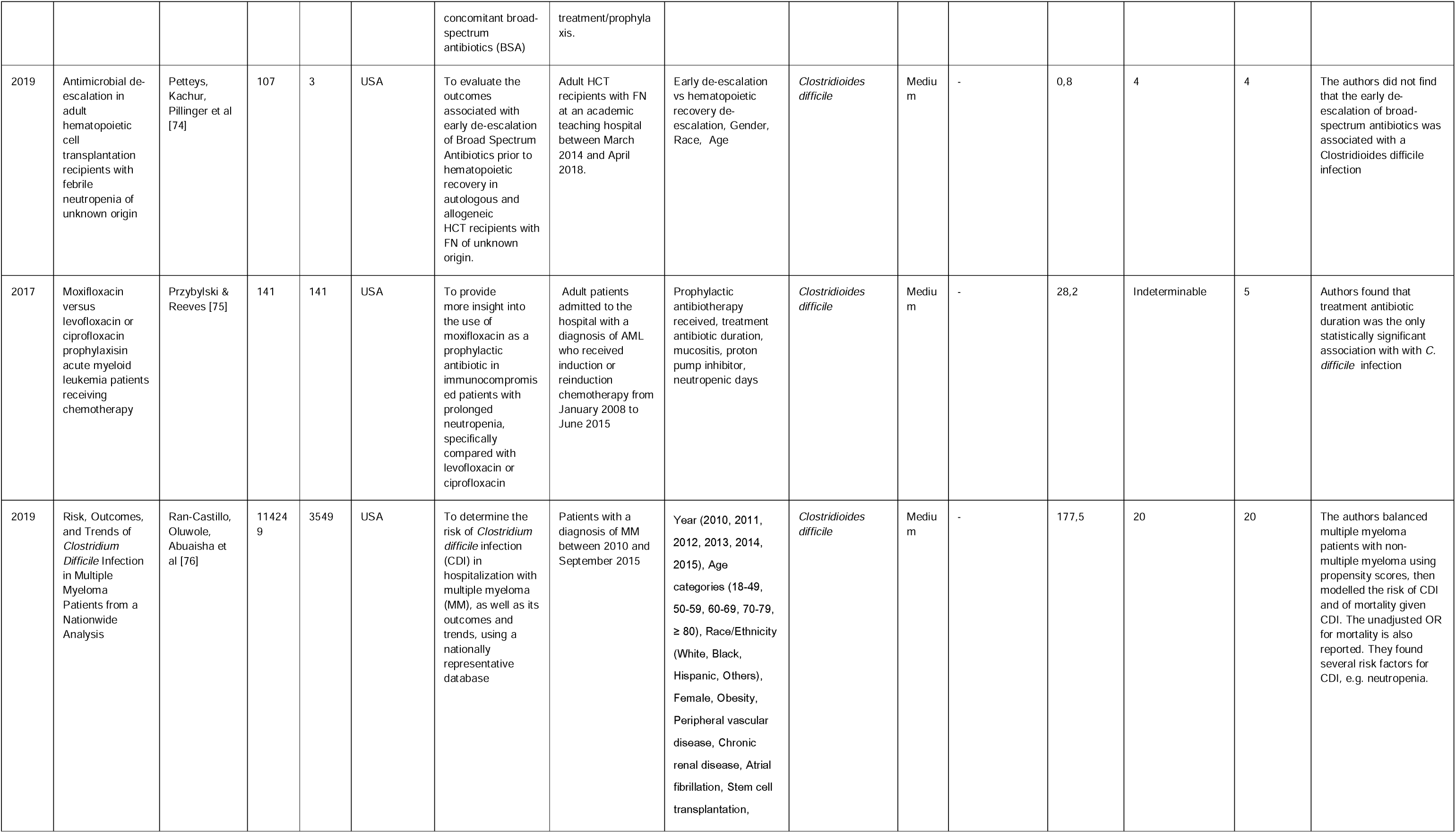
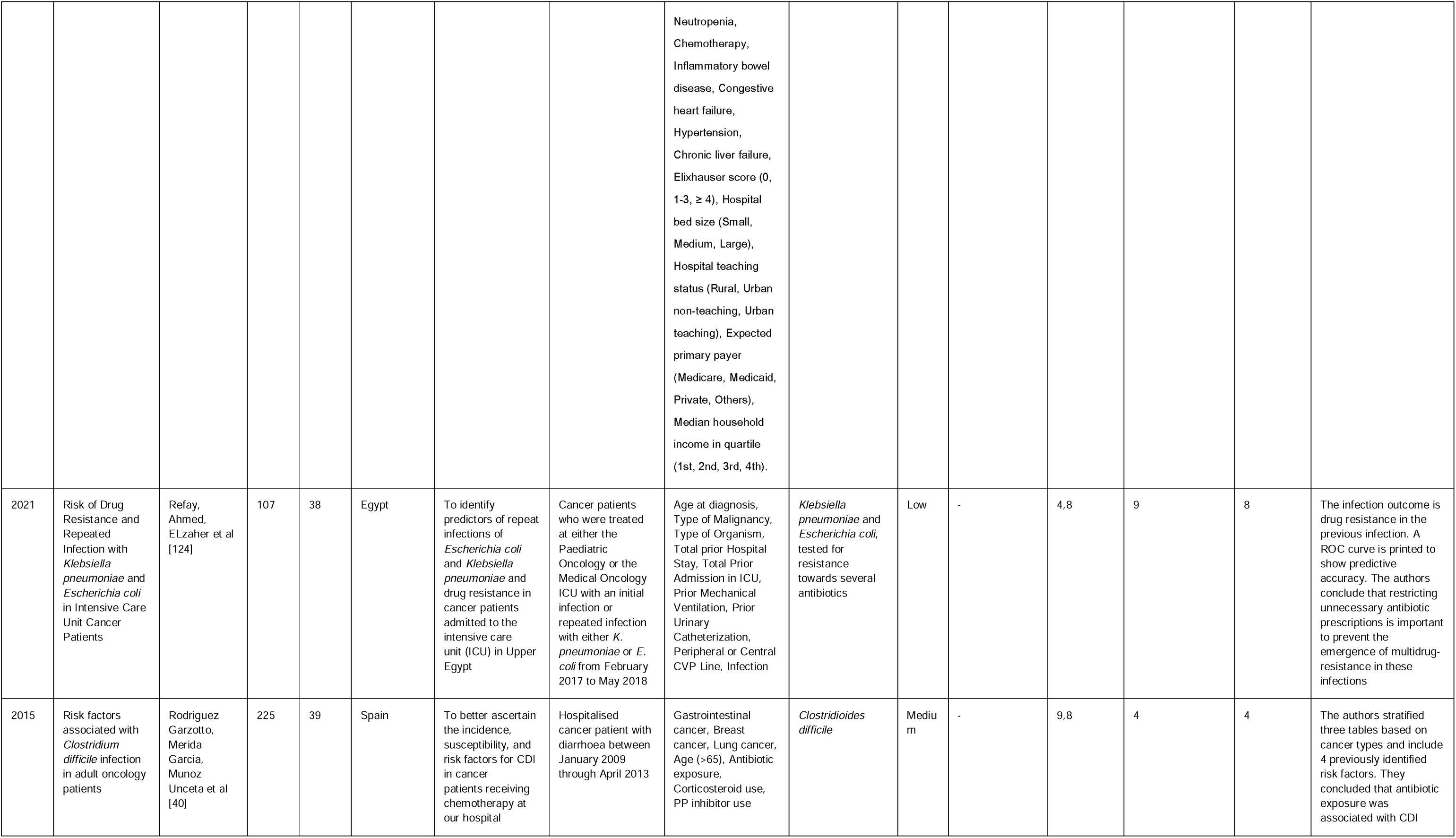
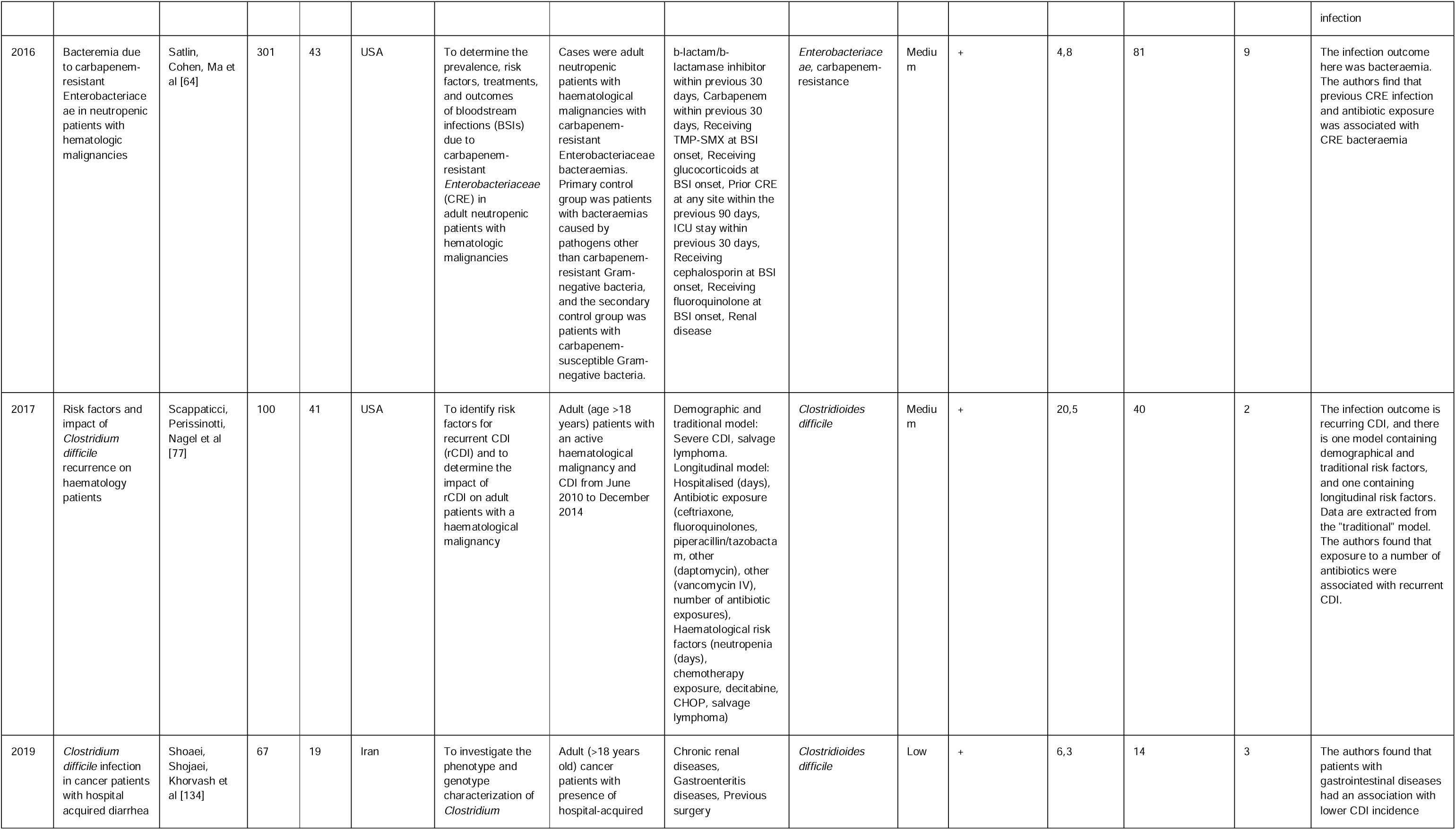
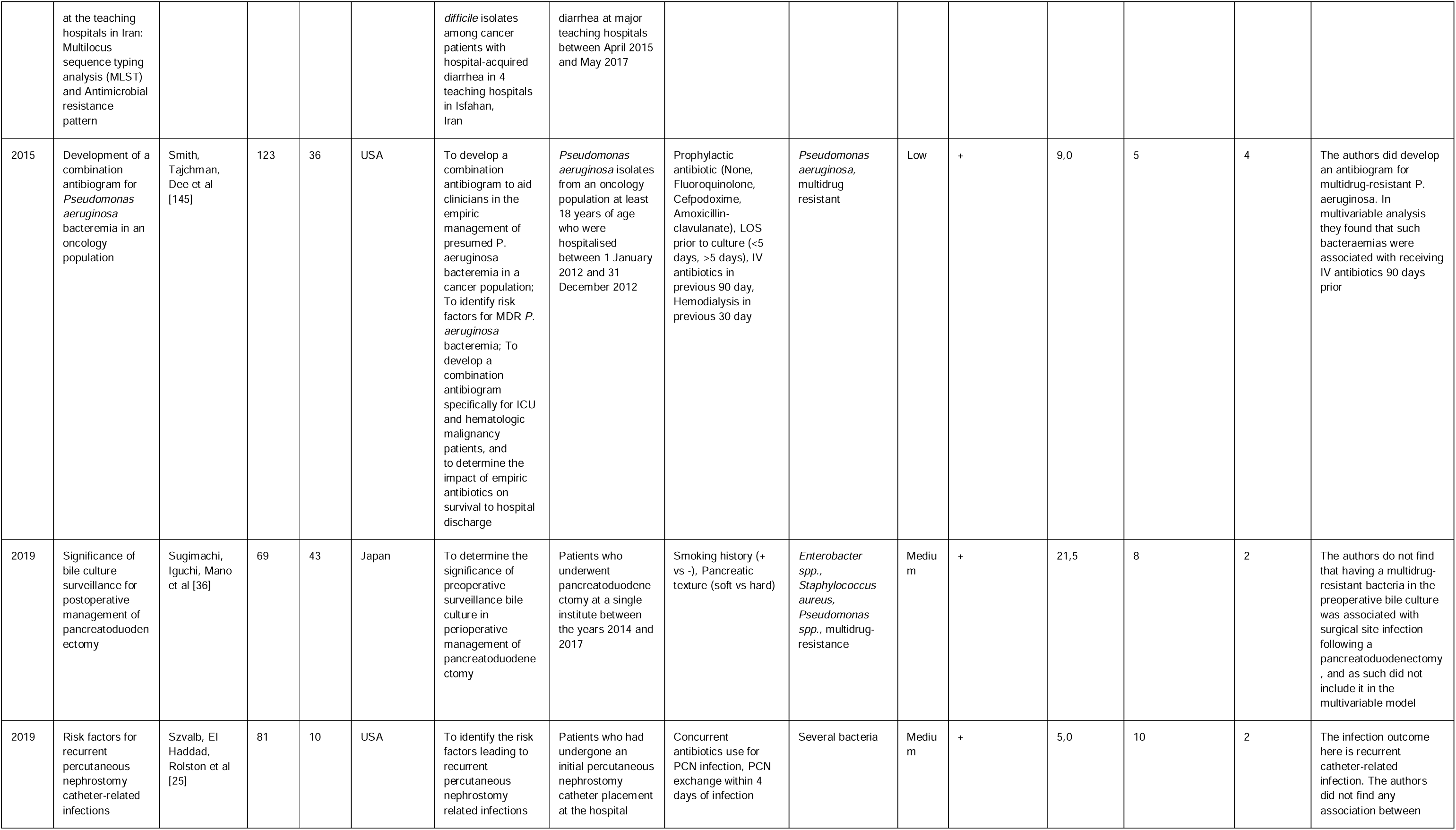
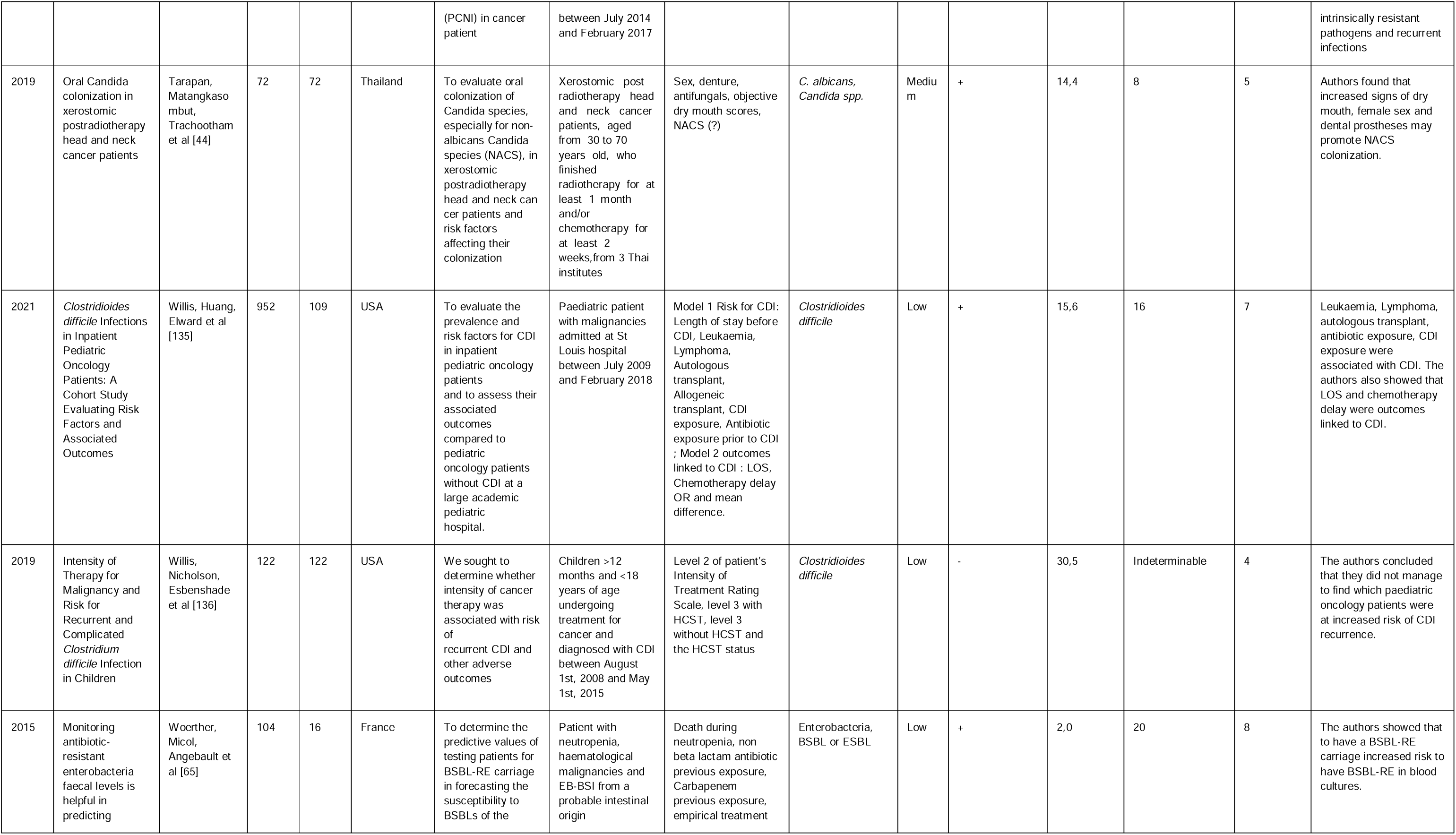
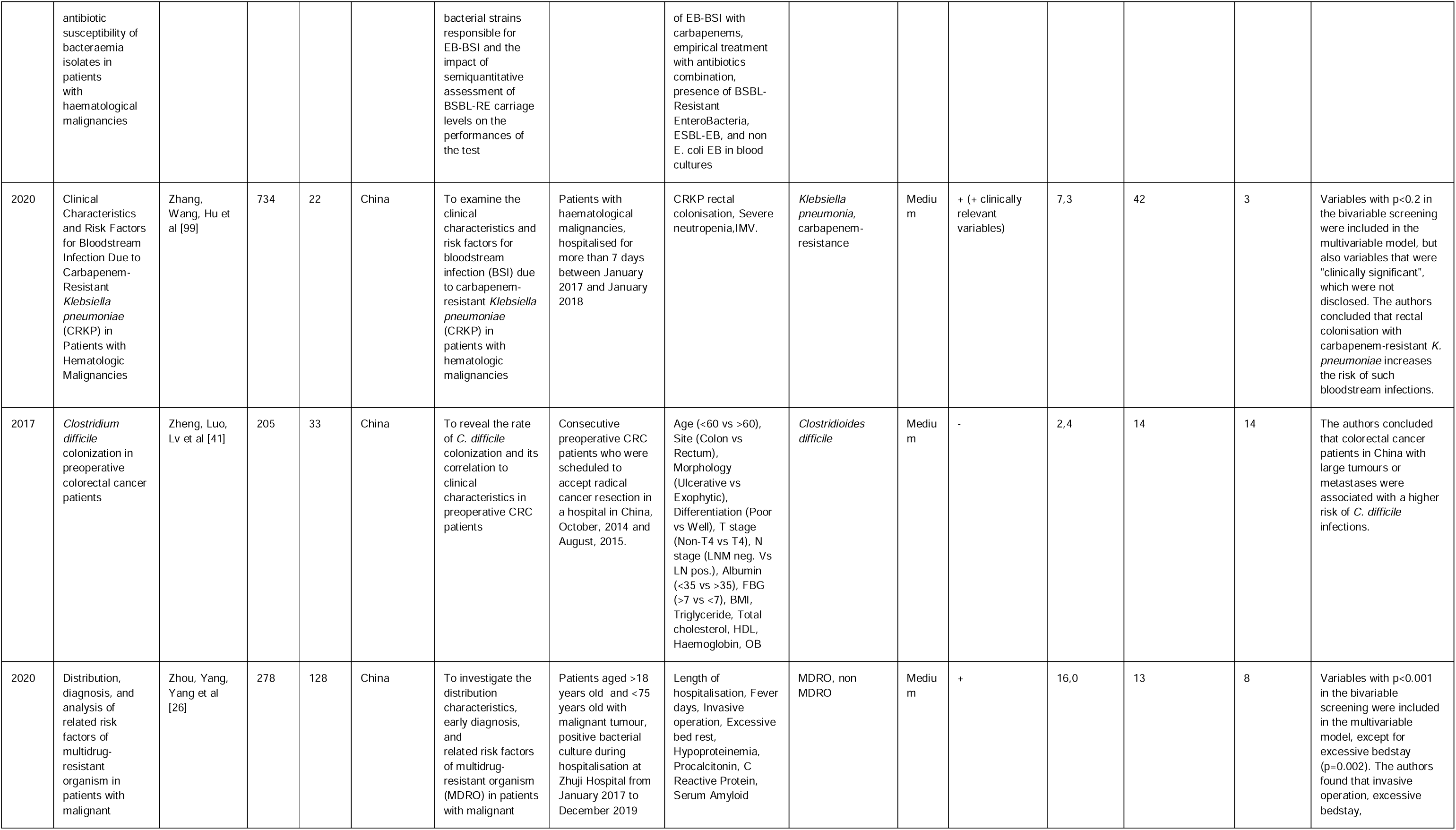
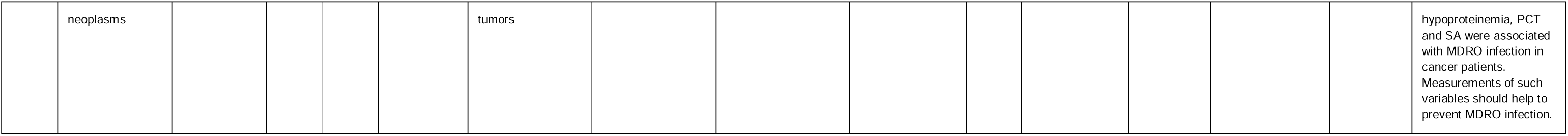
All included articles in the systematic review with an infection/colonisation outcome.

## Supplementary material 4

**Table S4.**
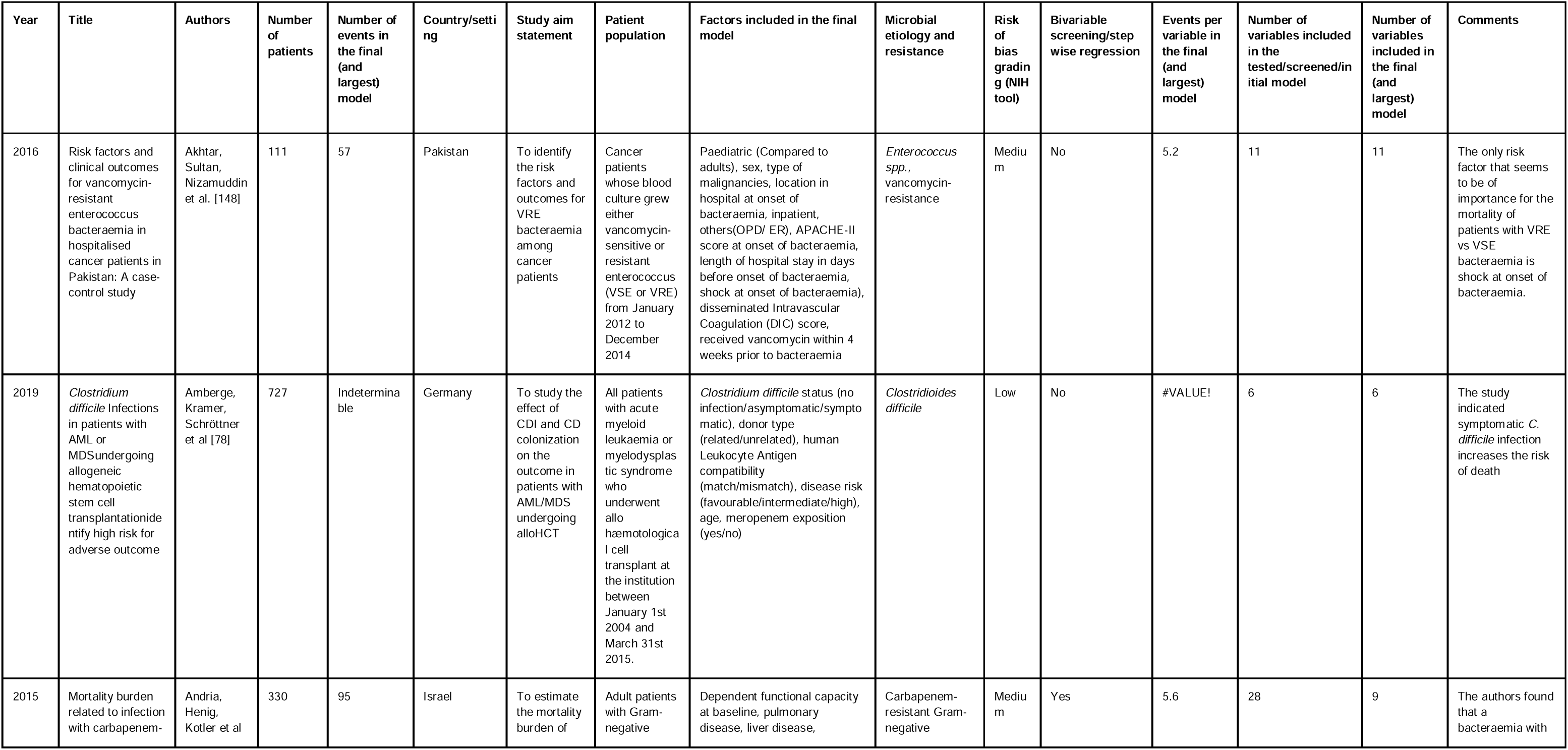
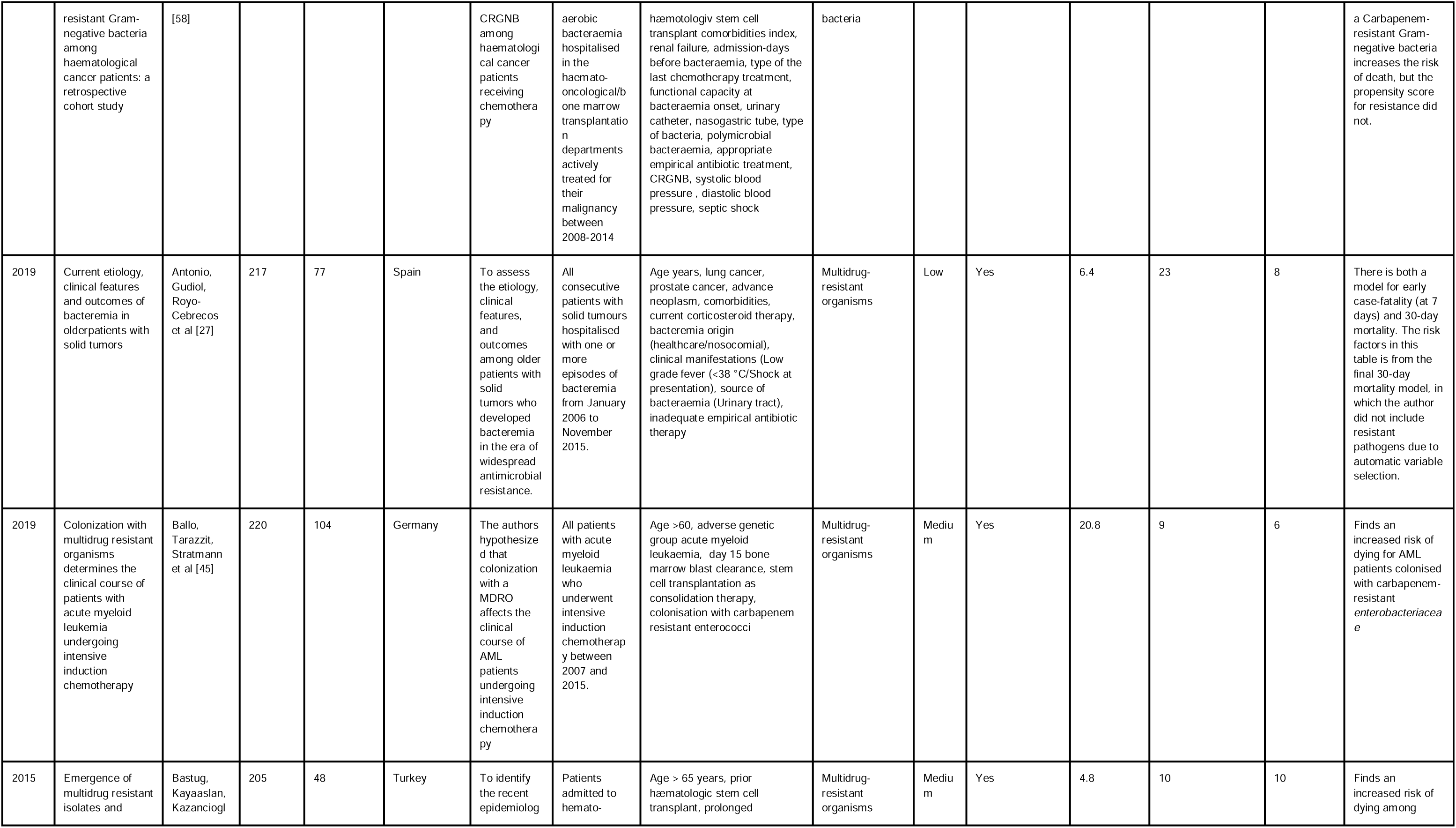
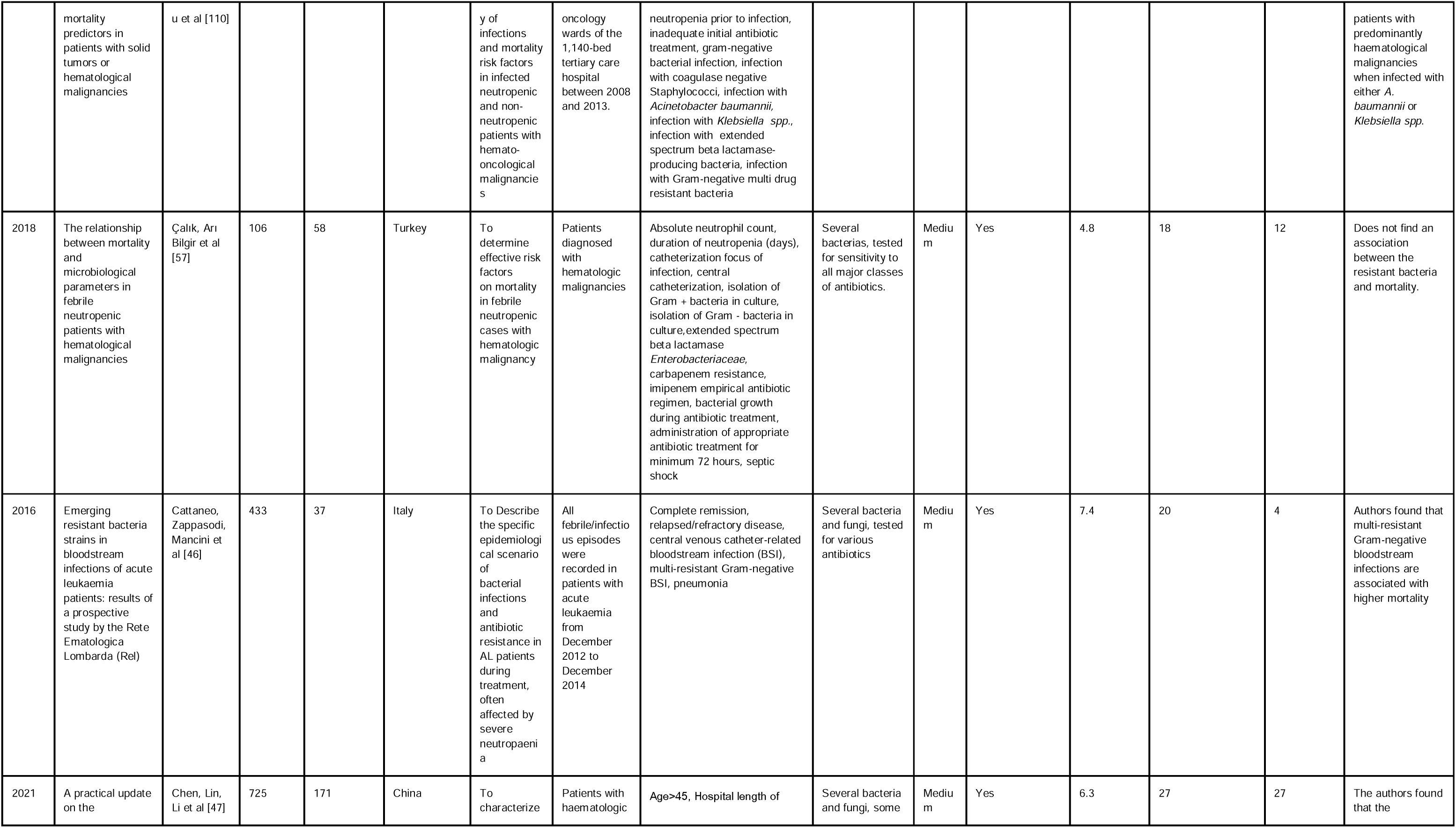
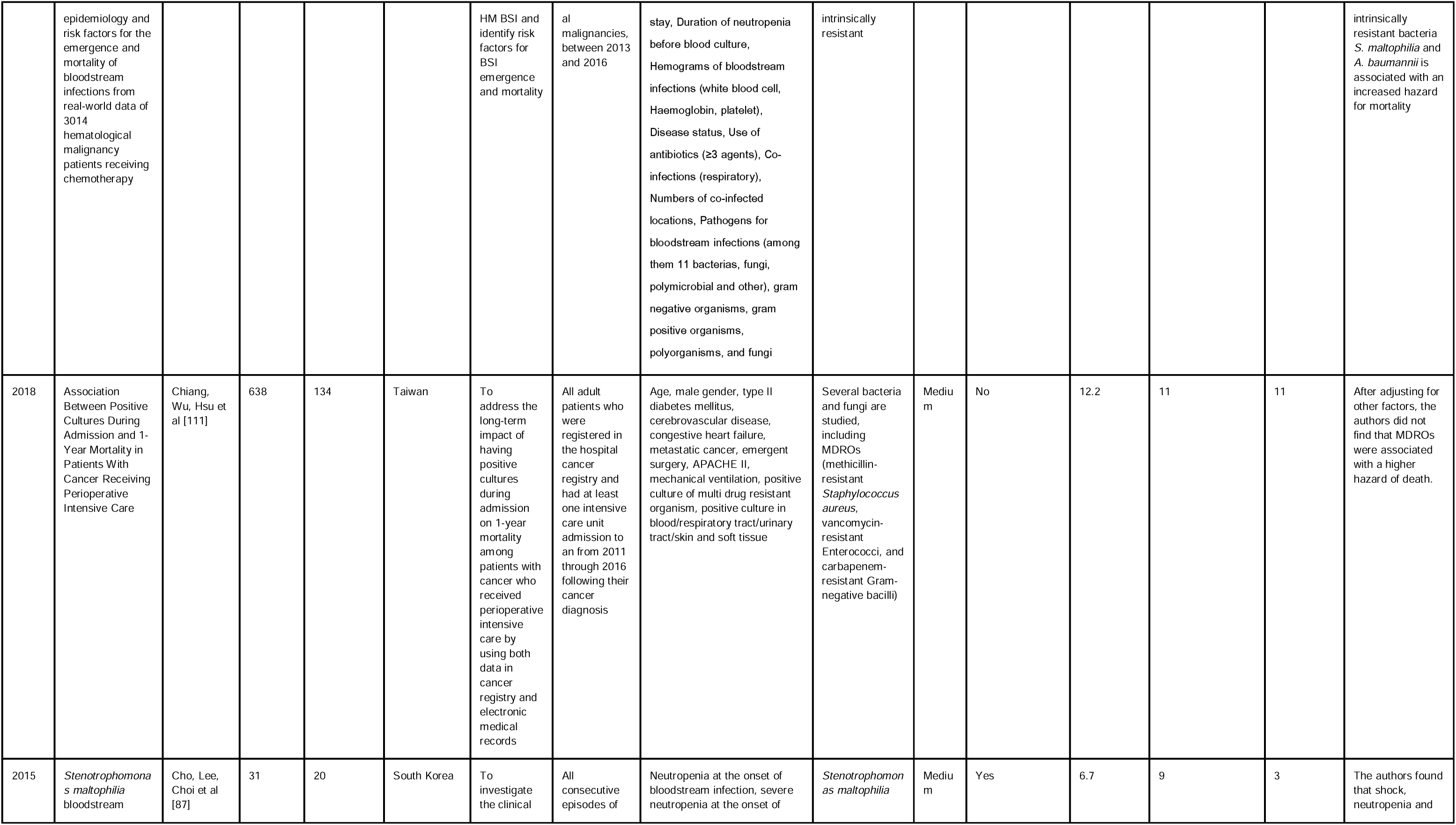
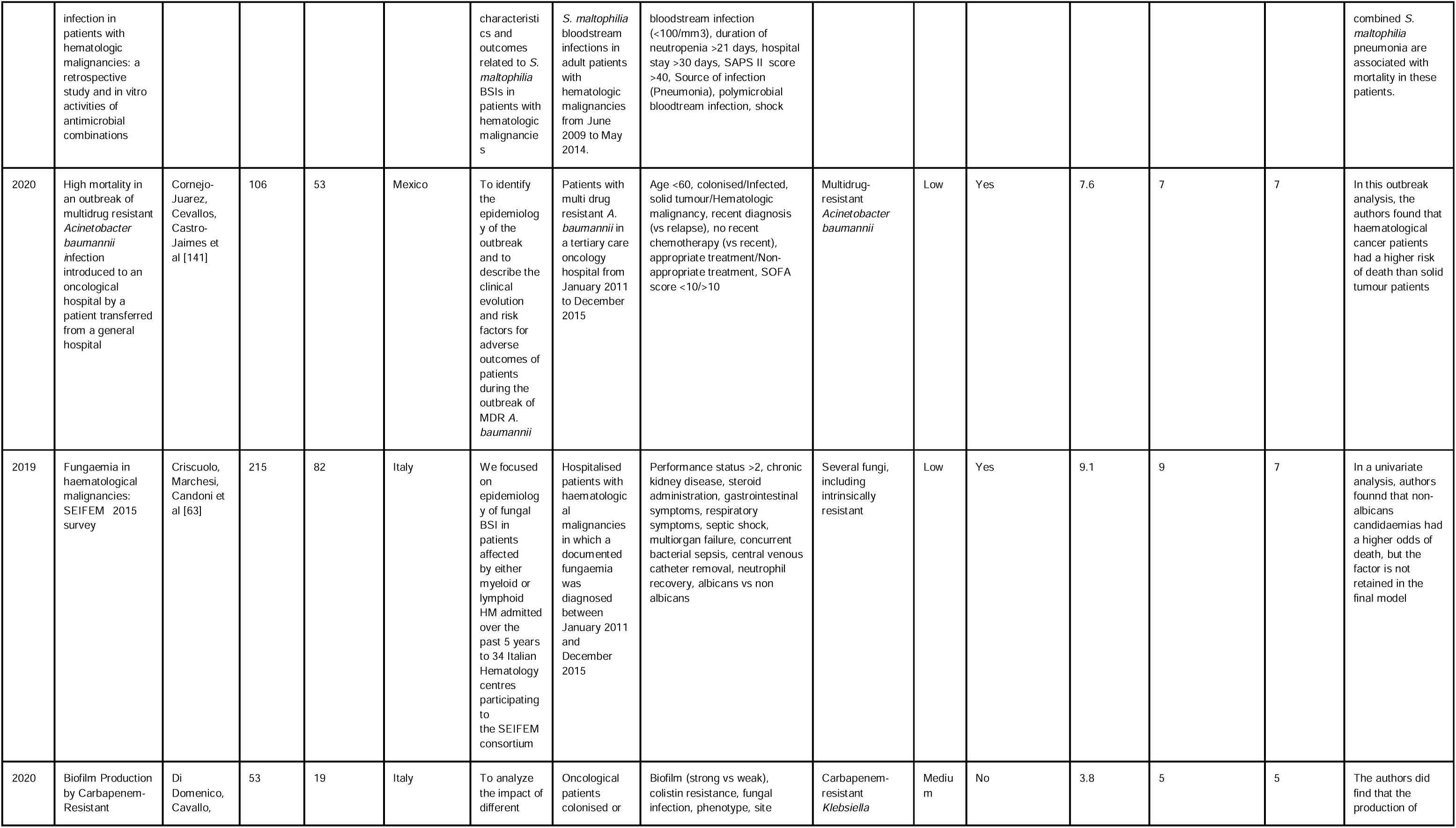
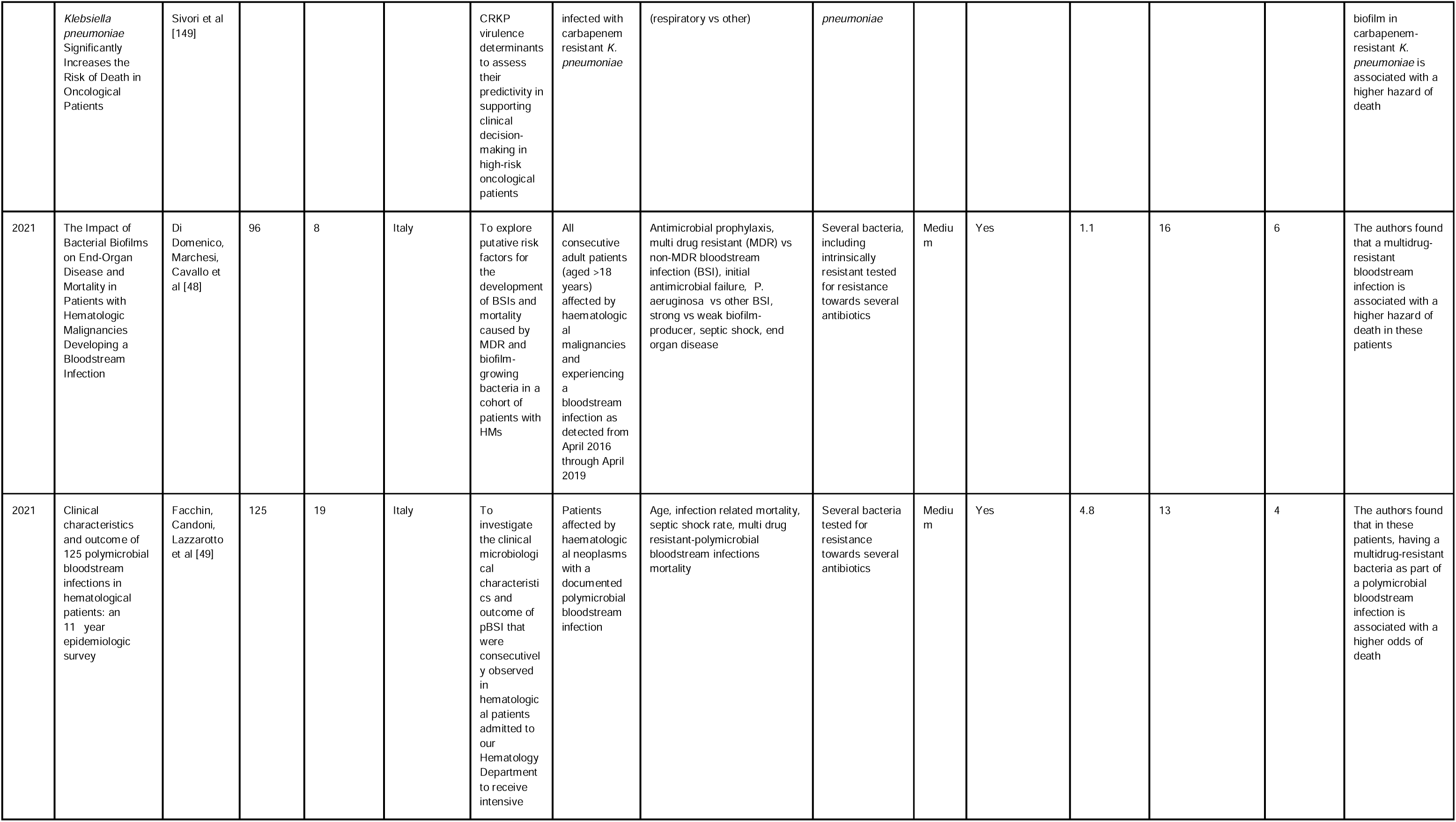
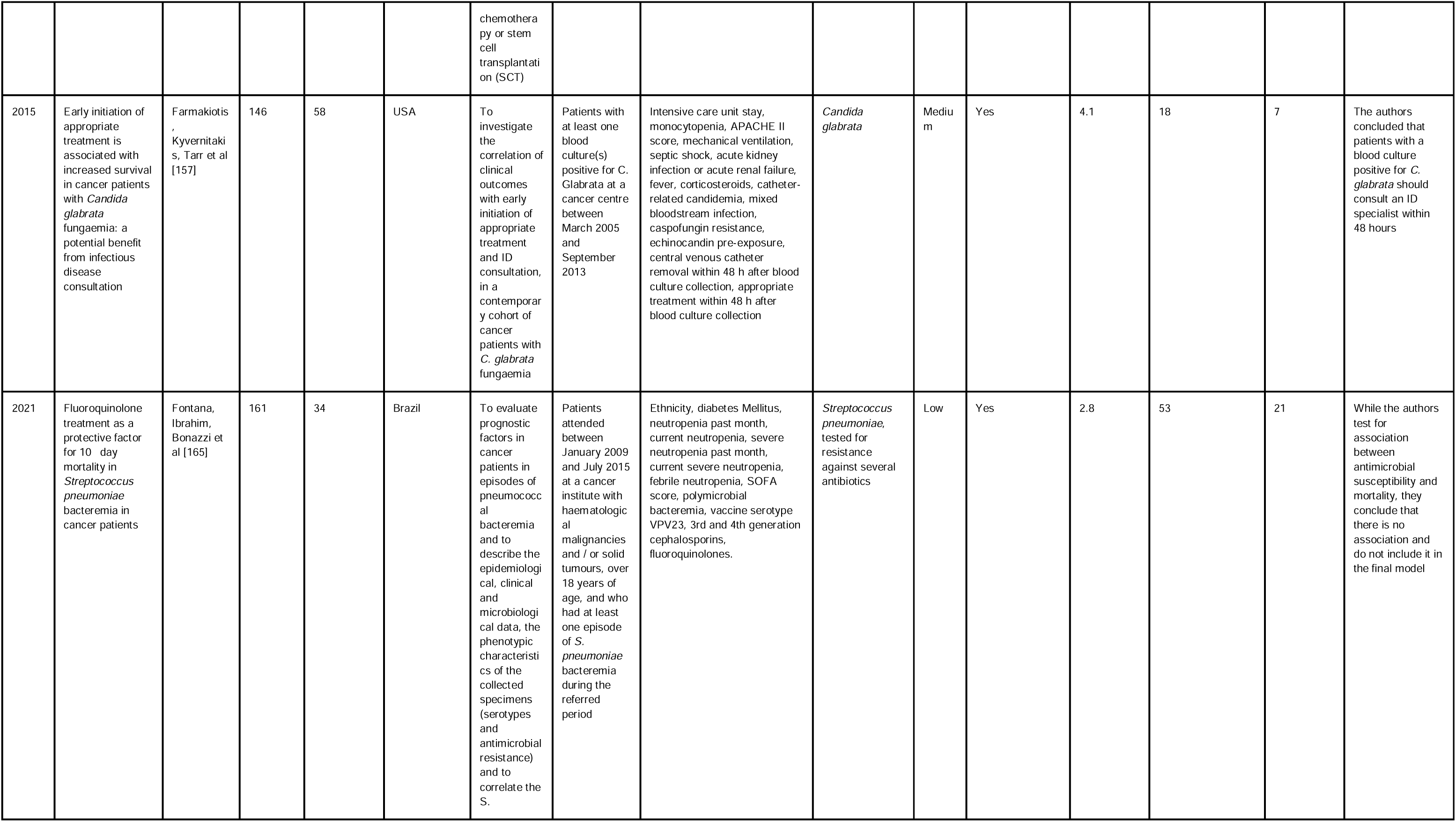
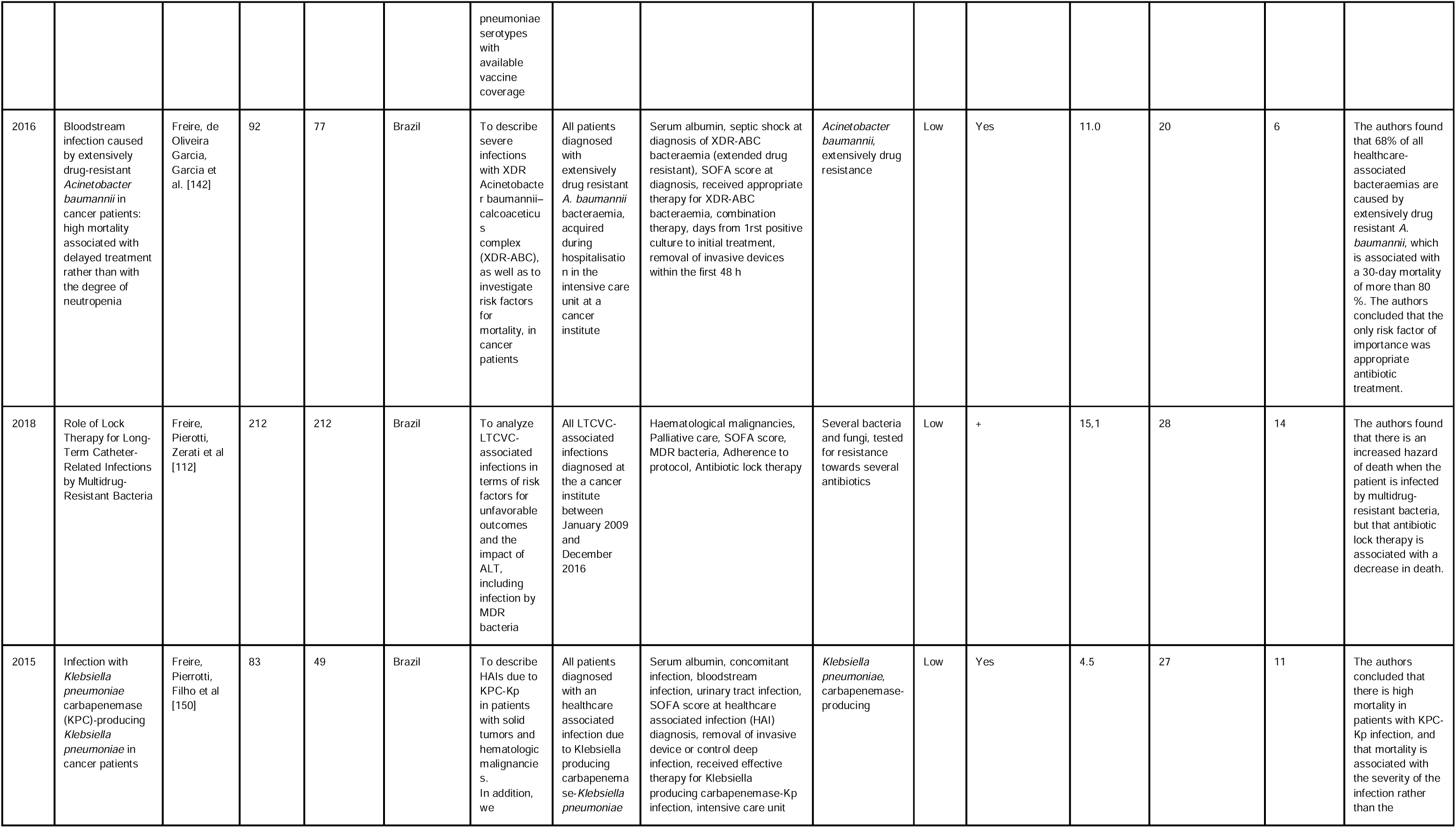
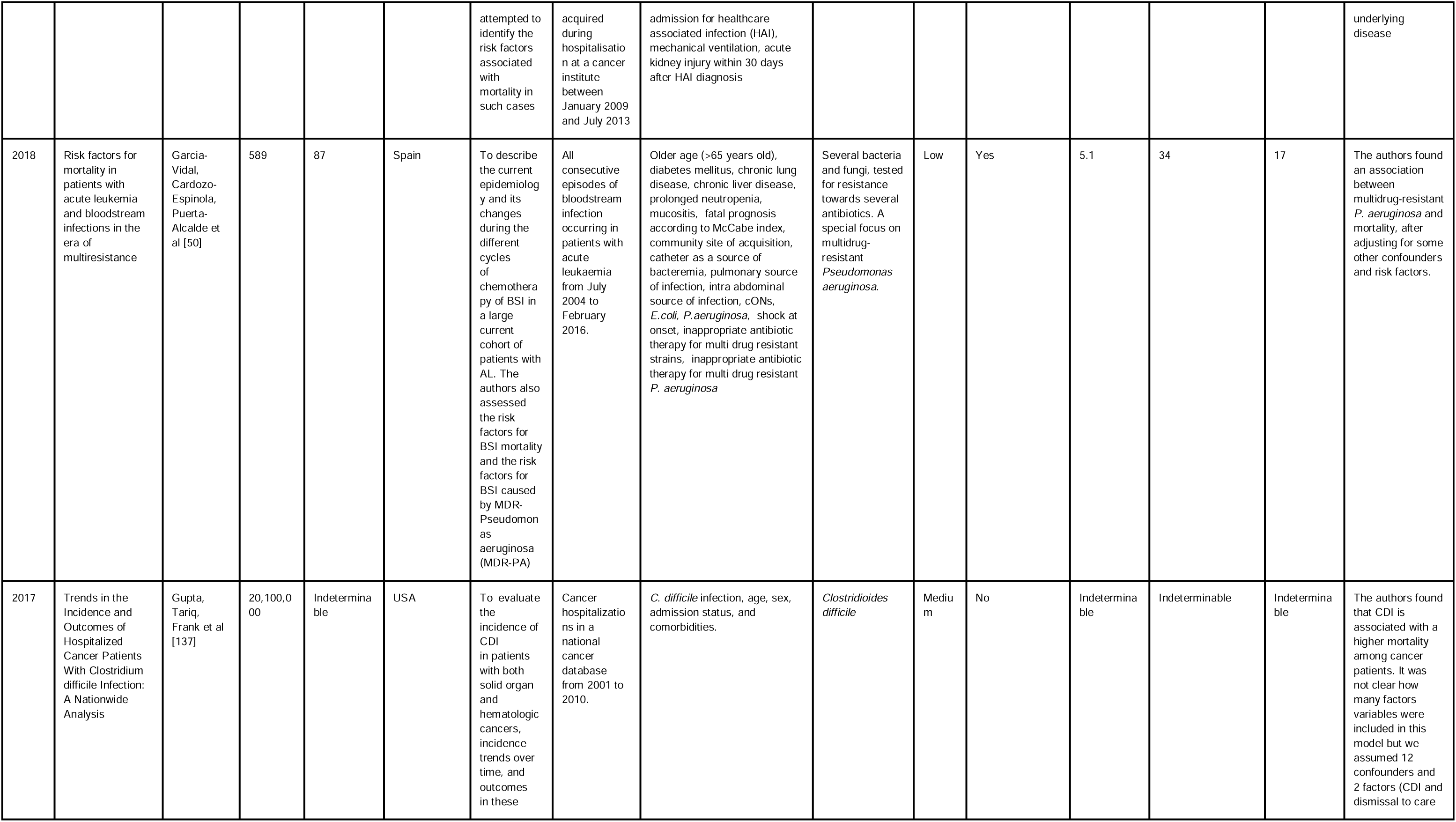
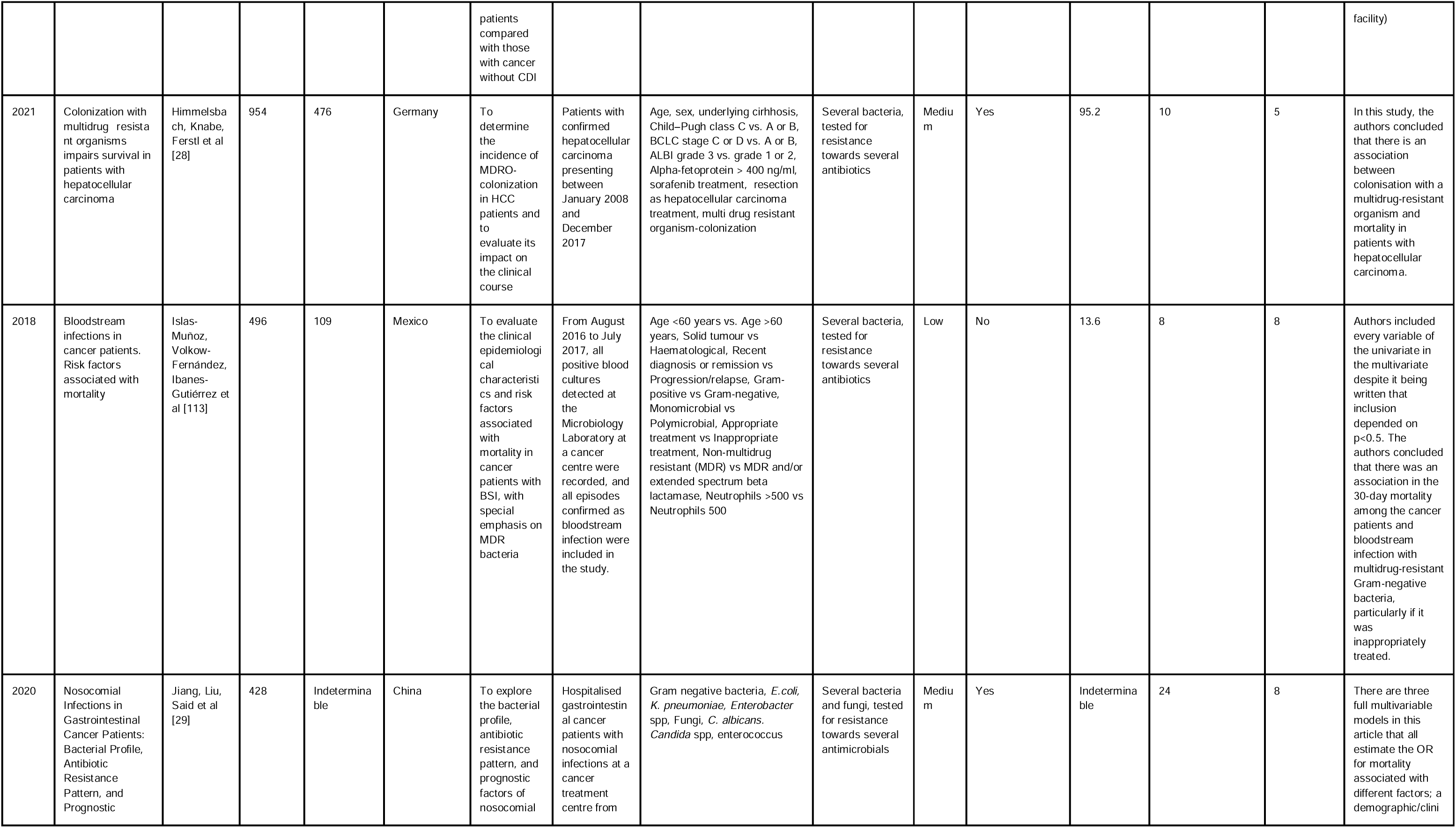
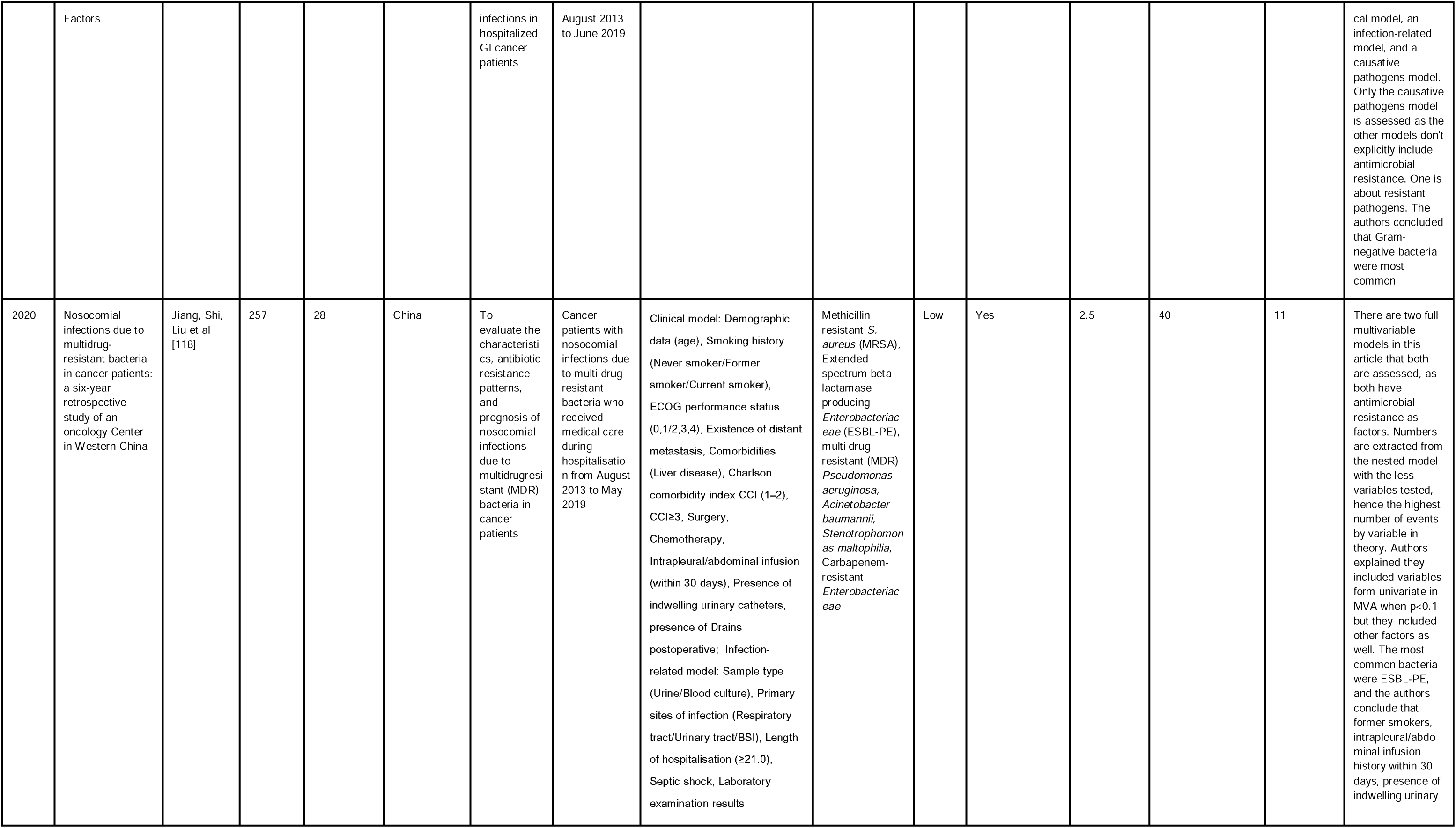
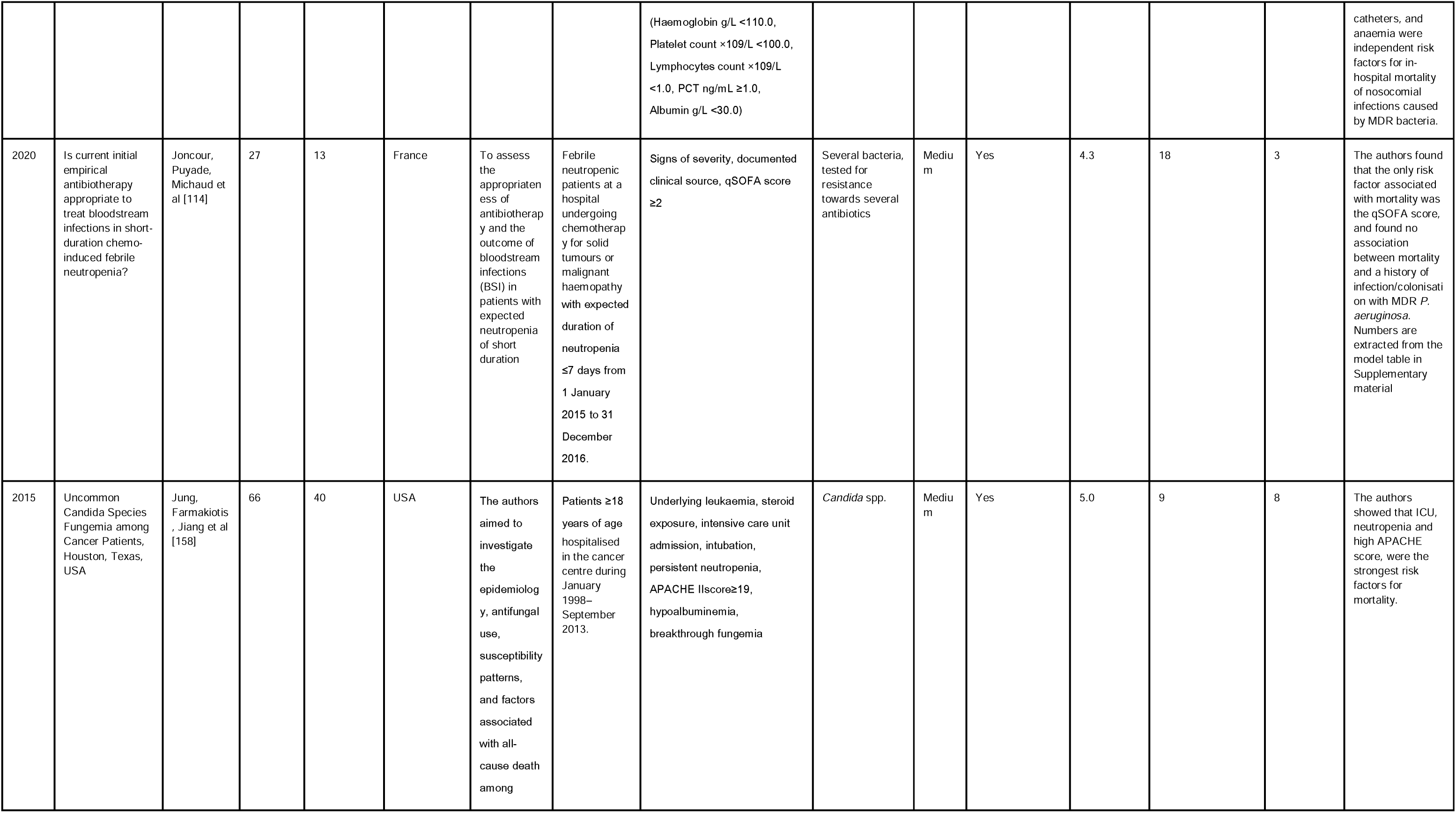
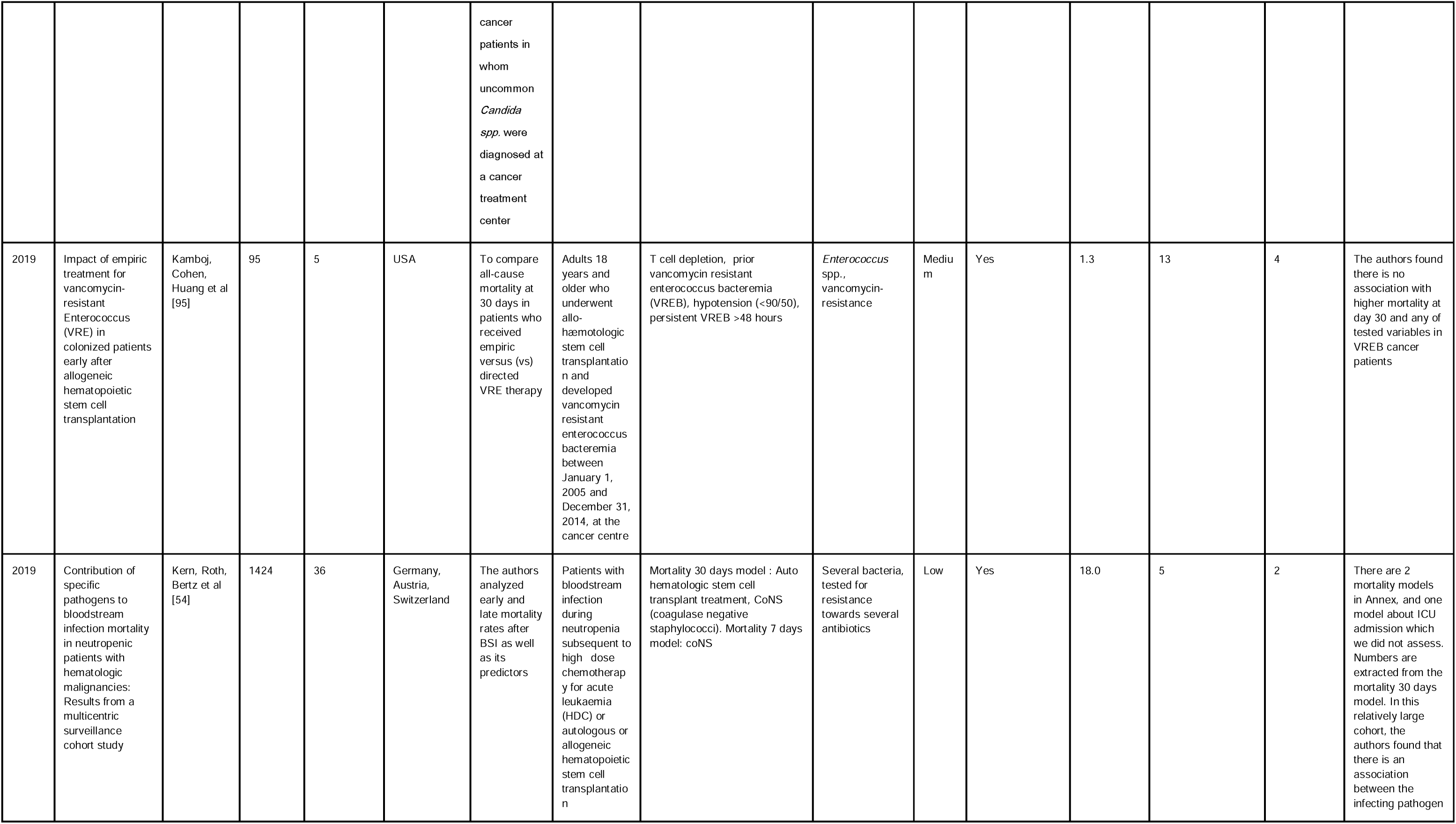
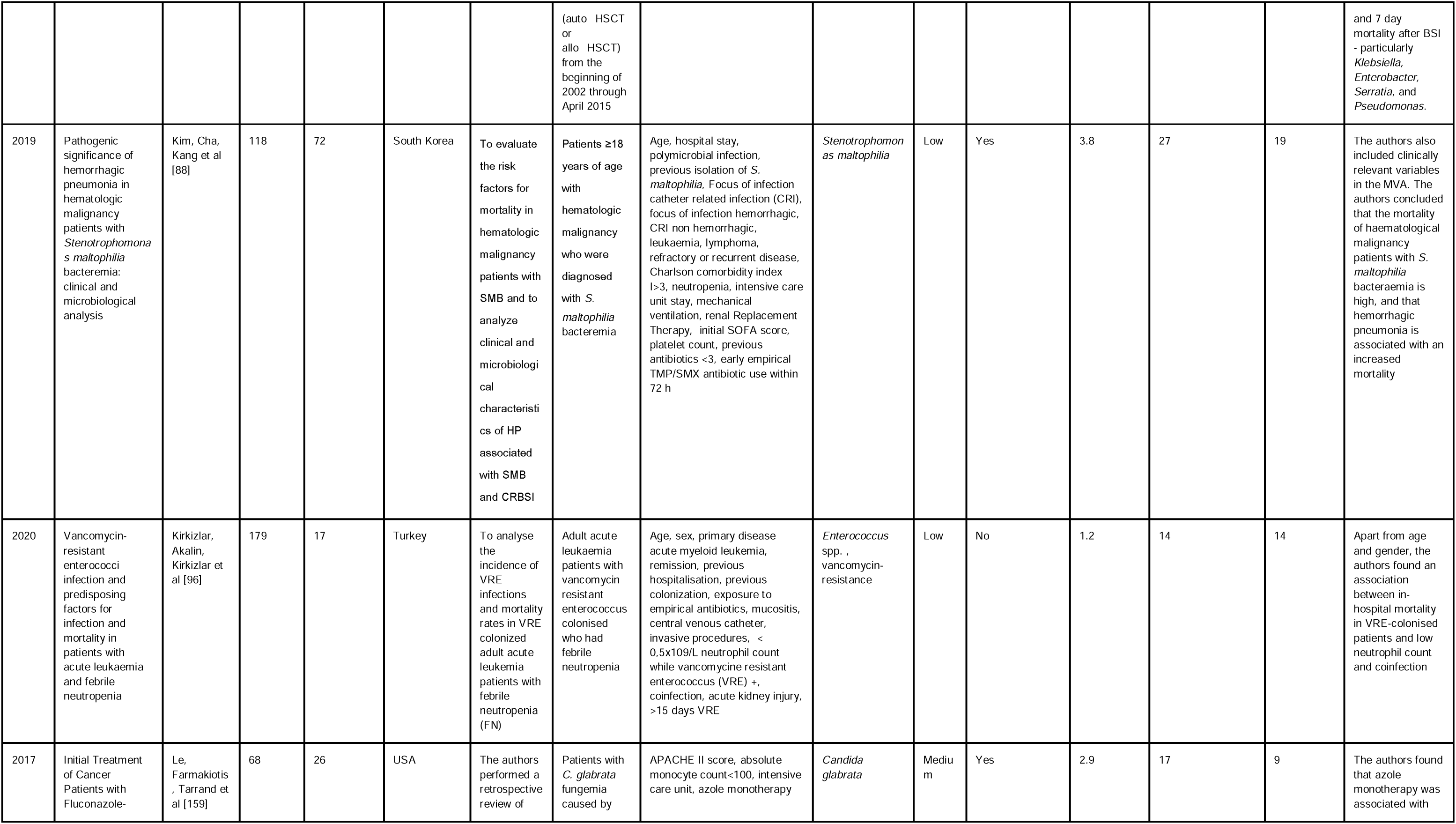
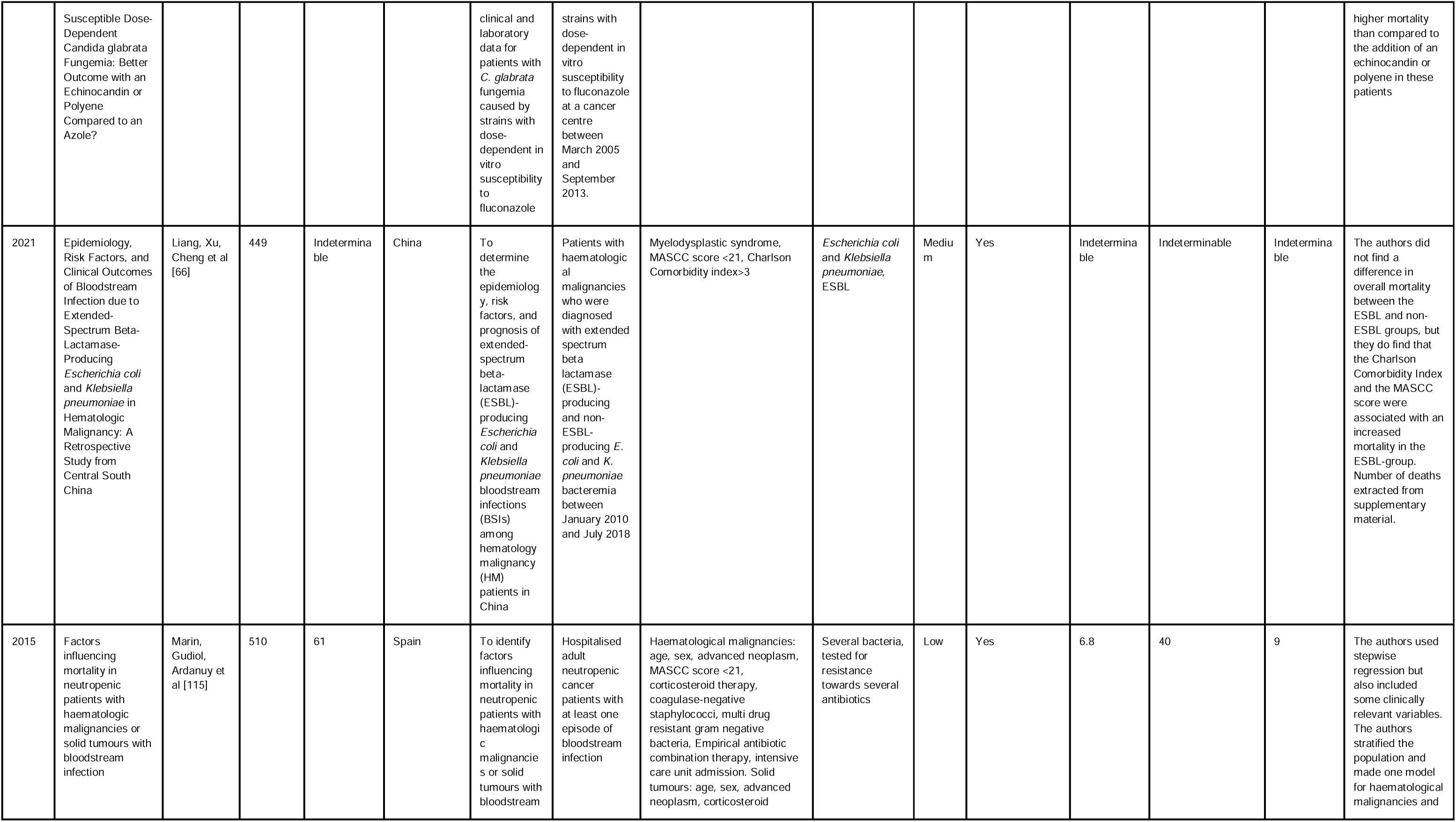
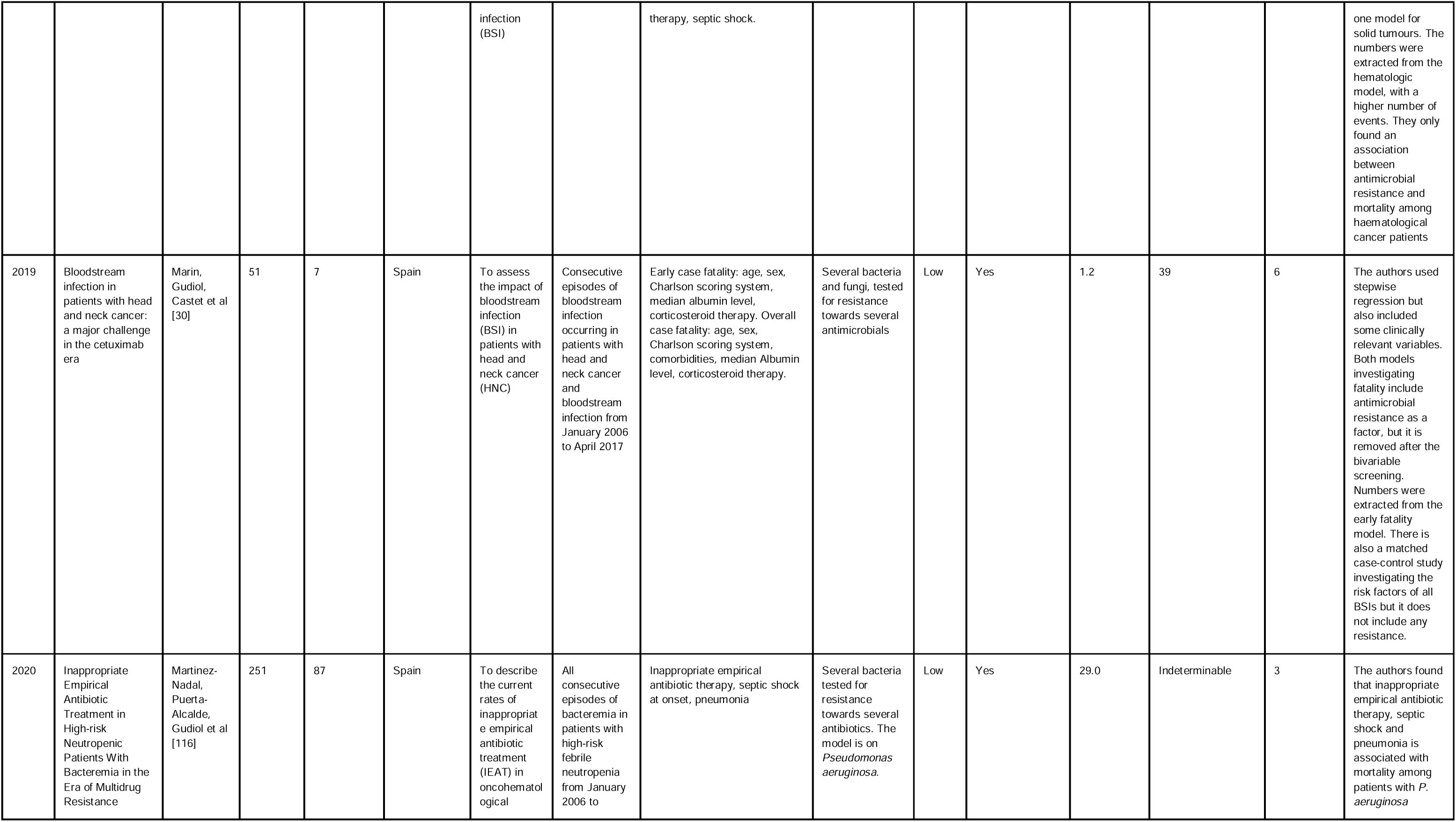
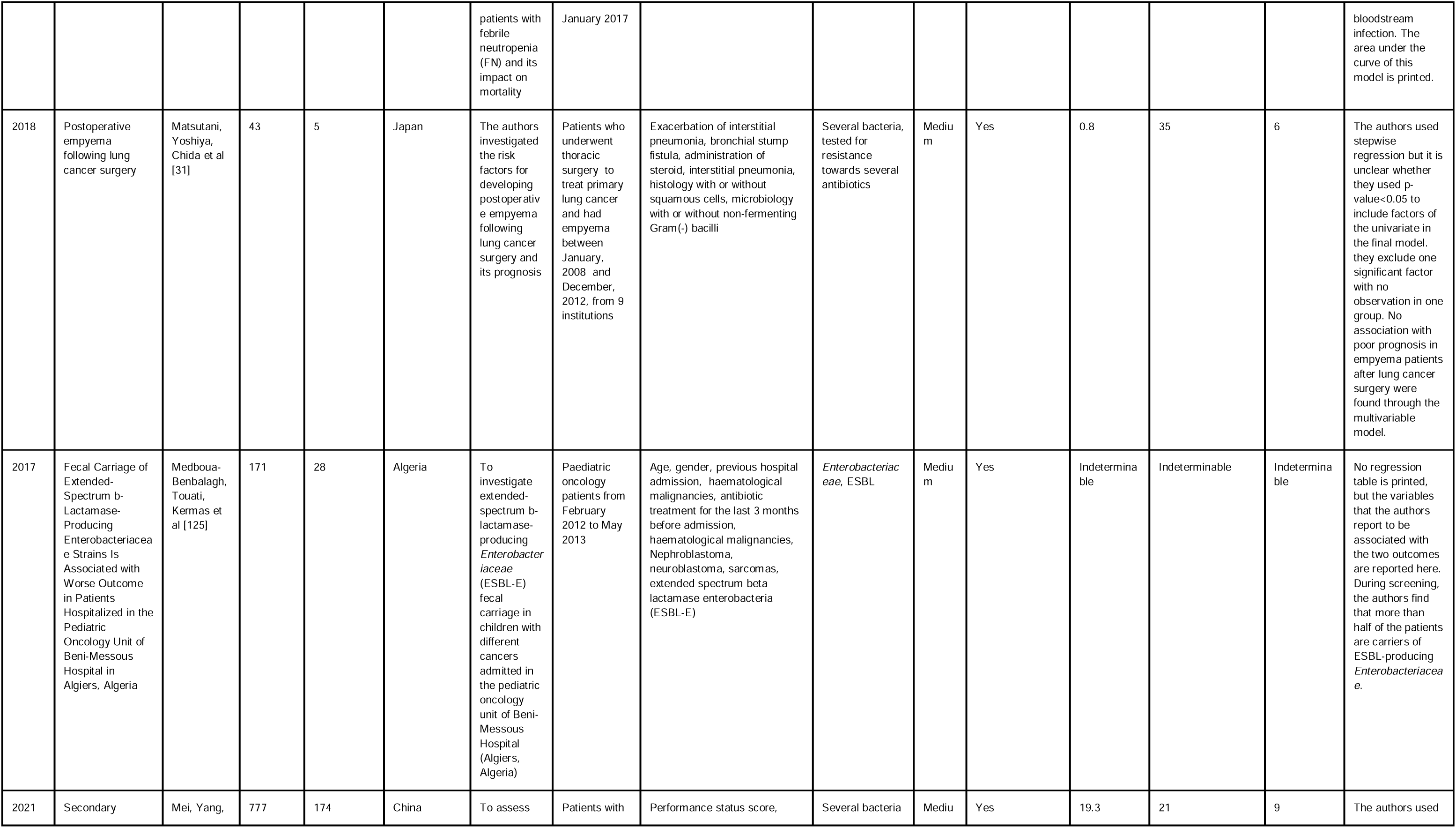
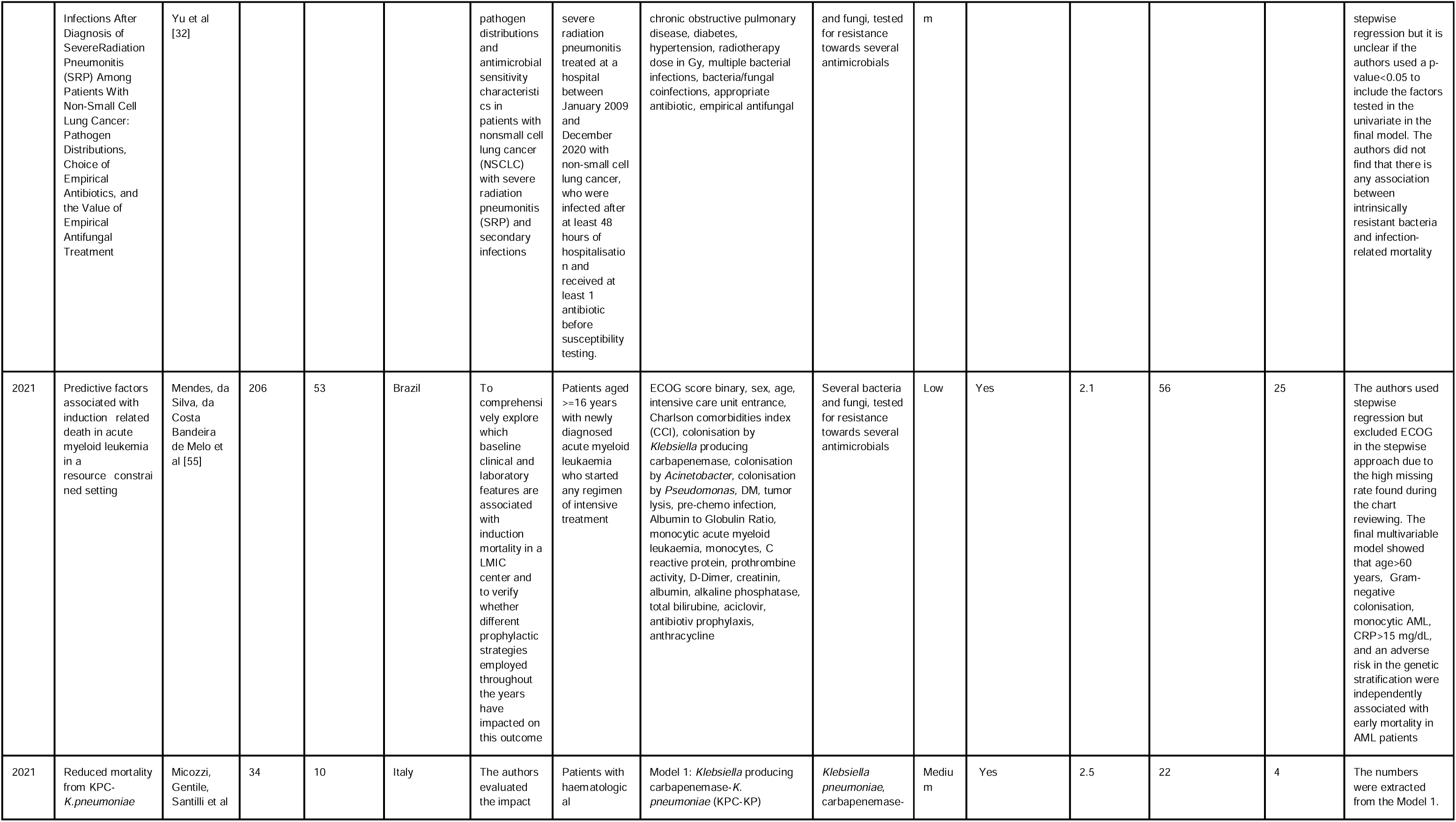
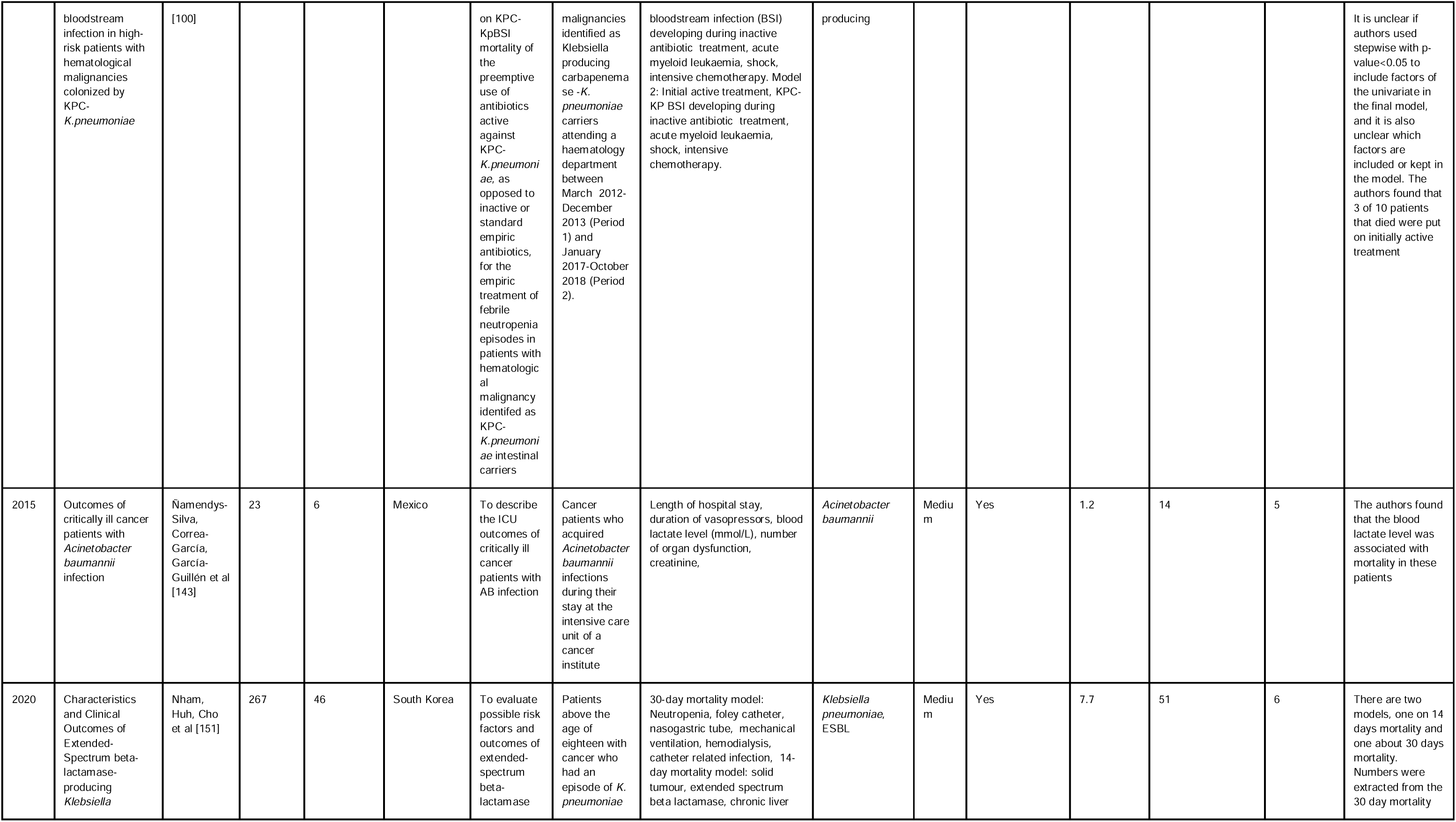
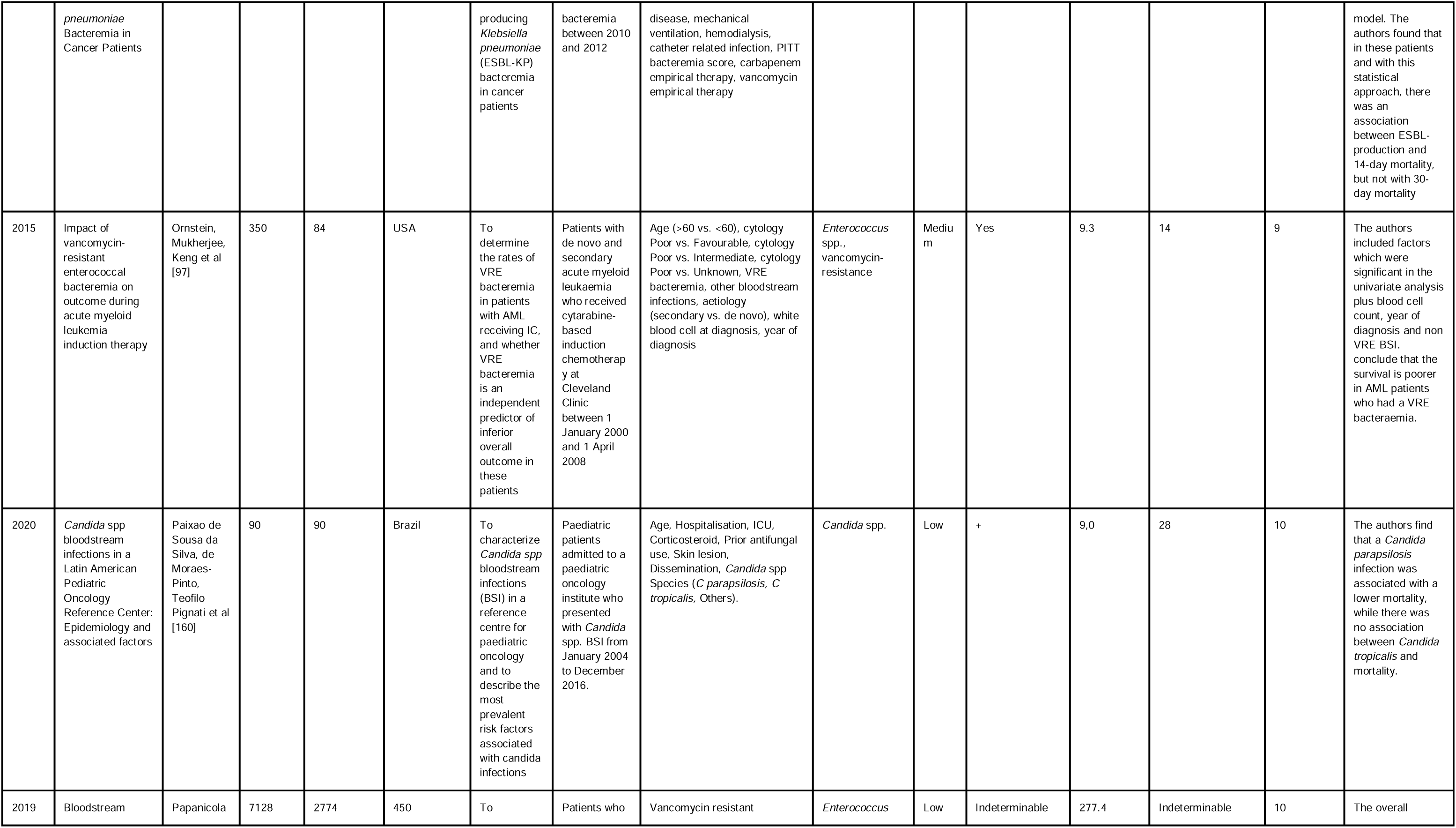
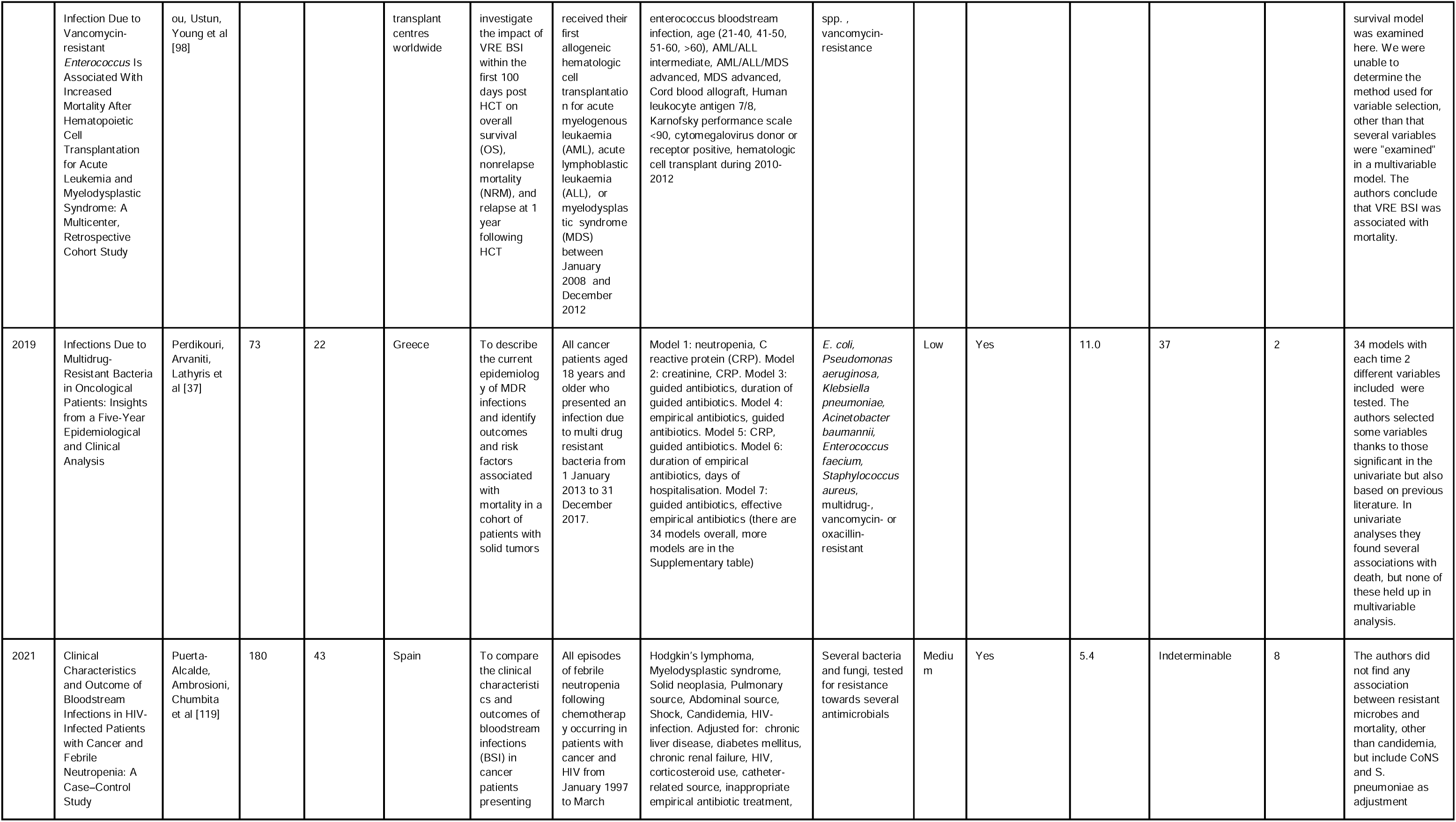
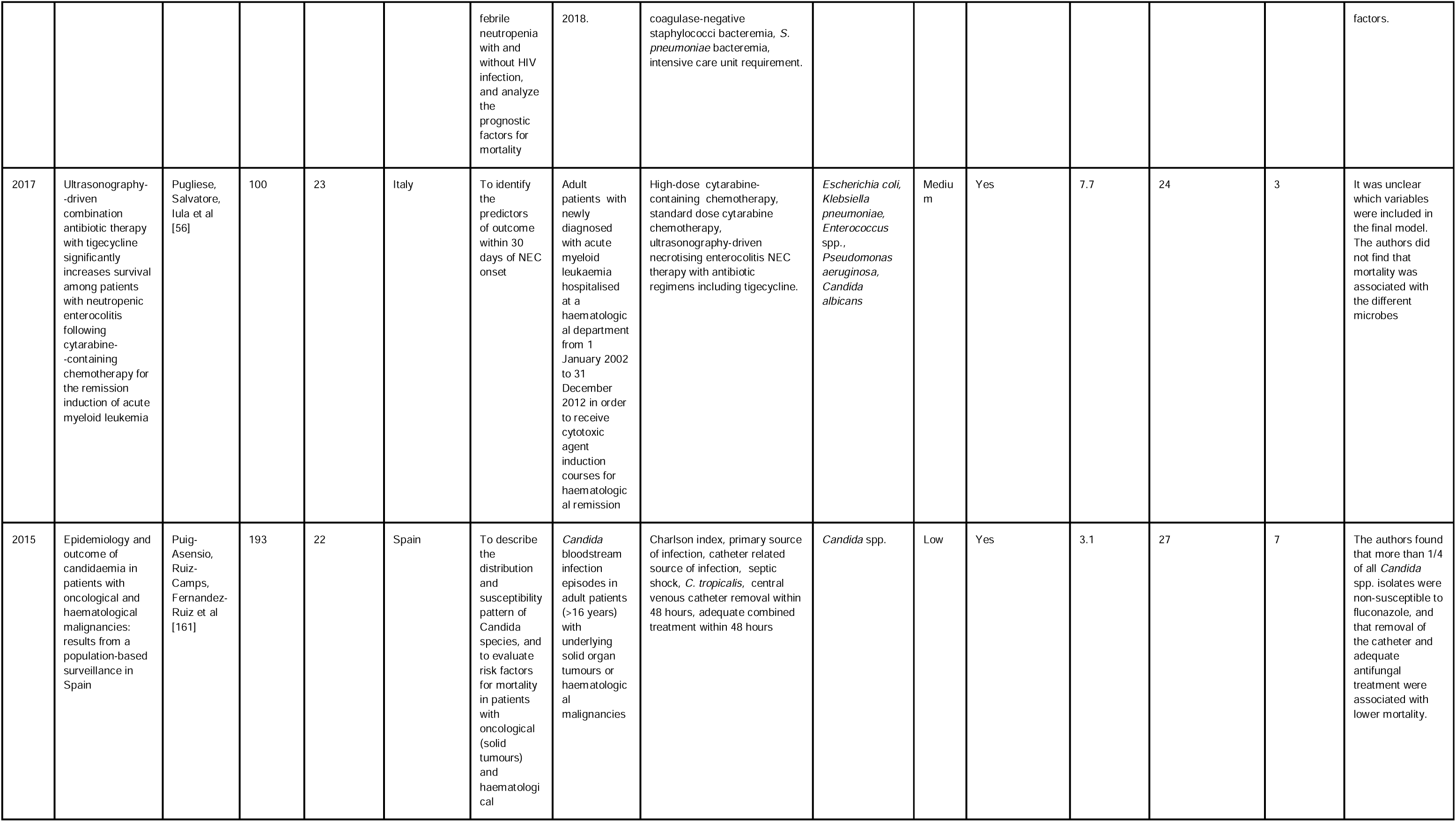
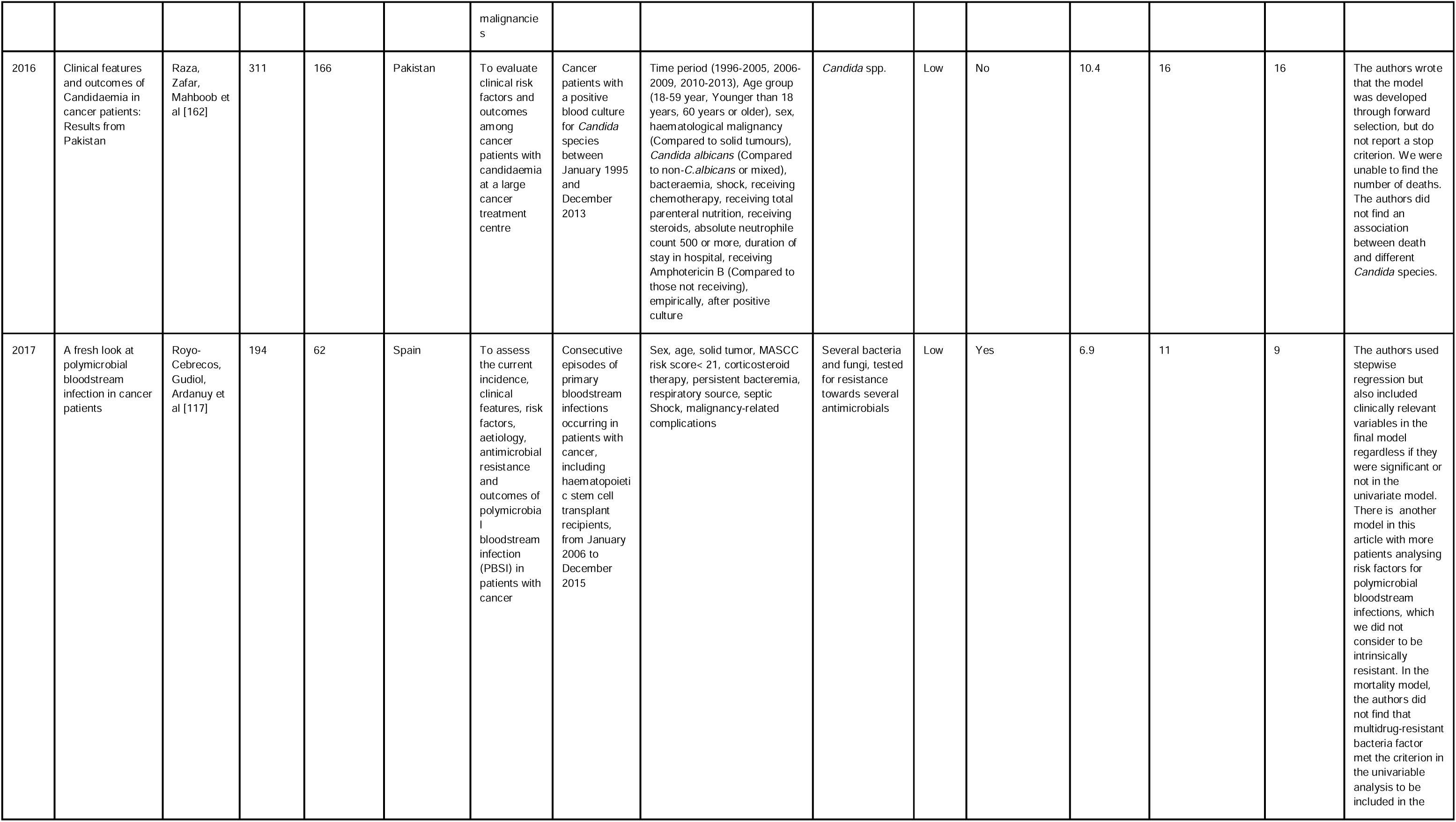
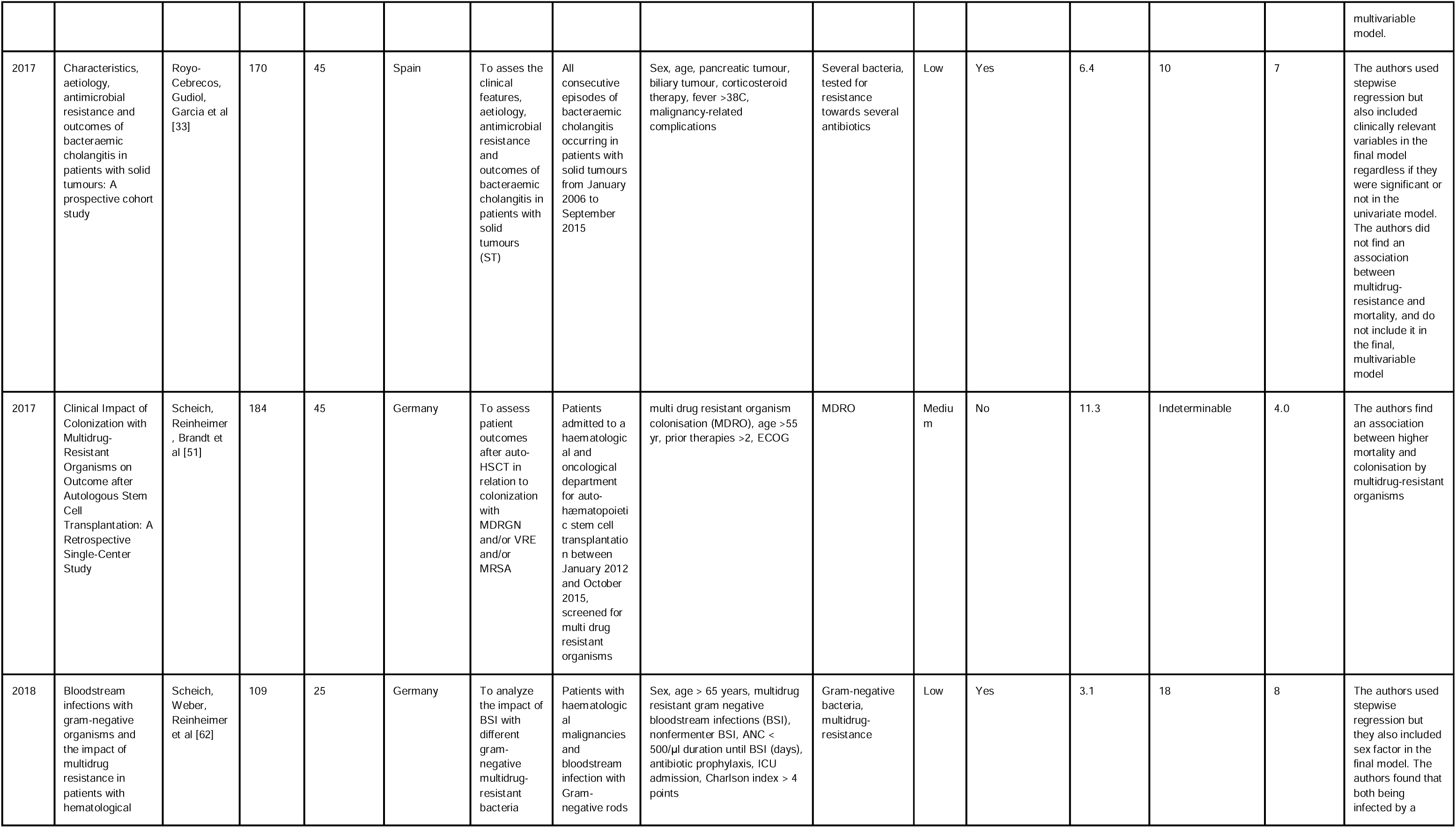
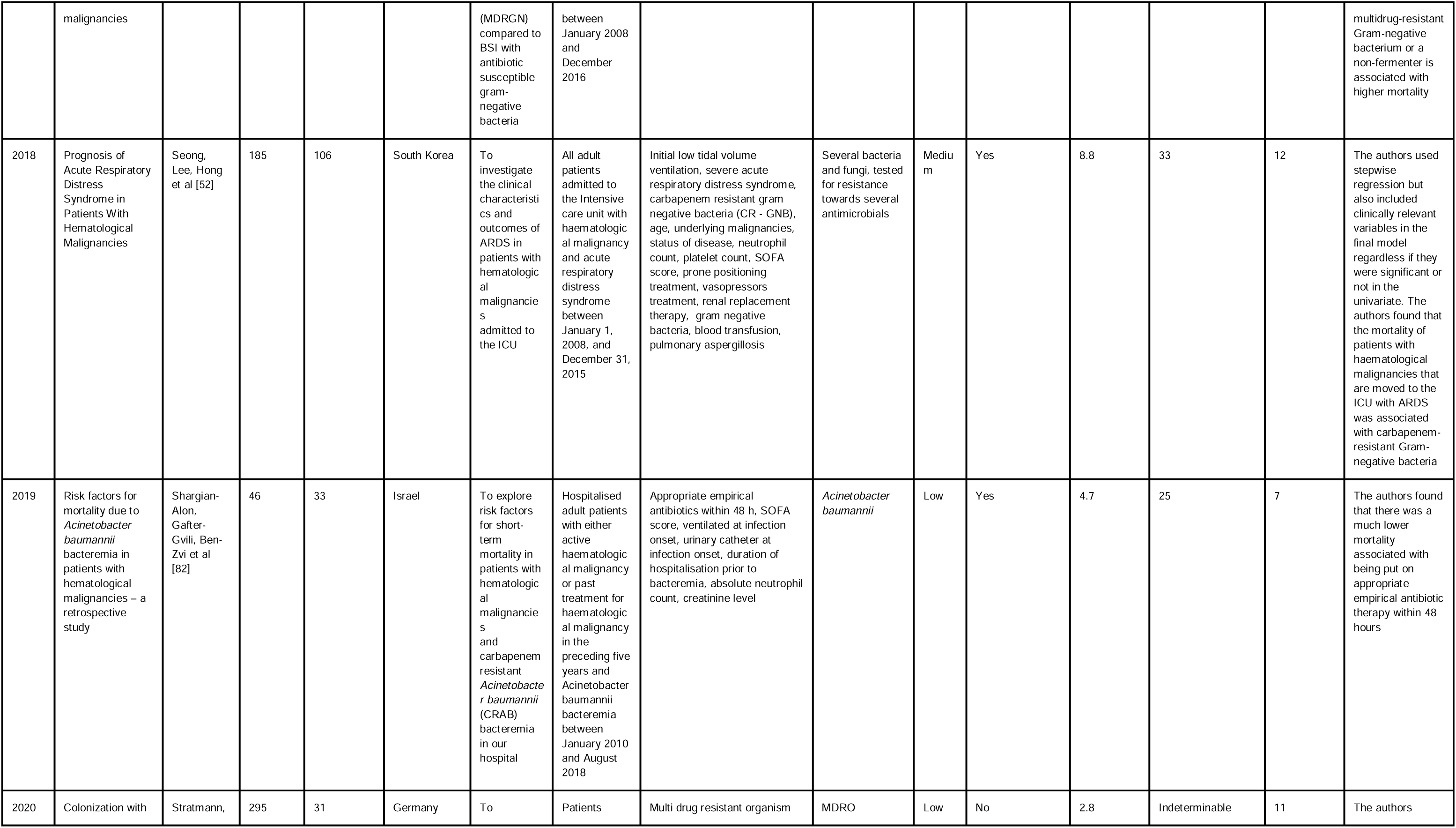
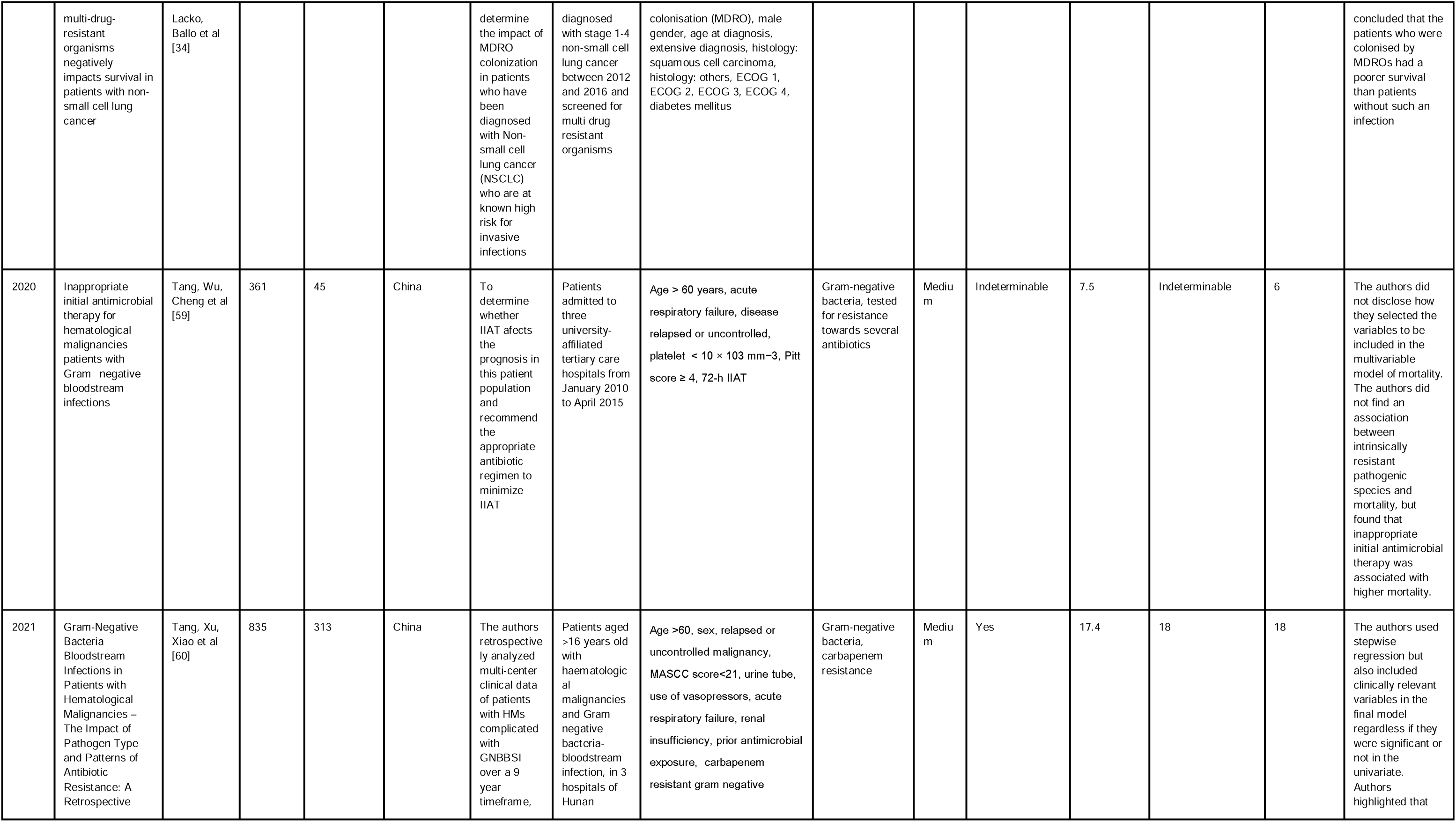
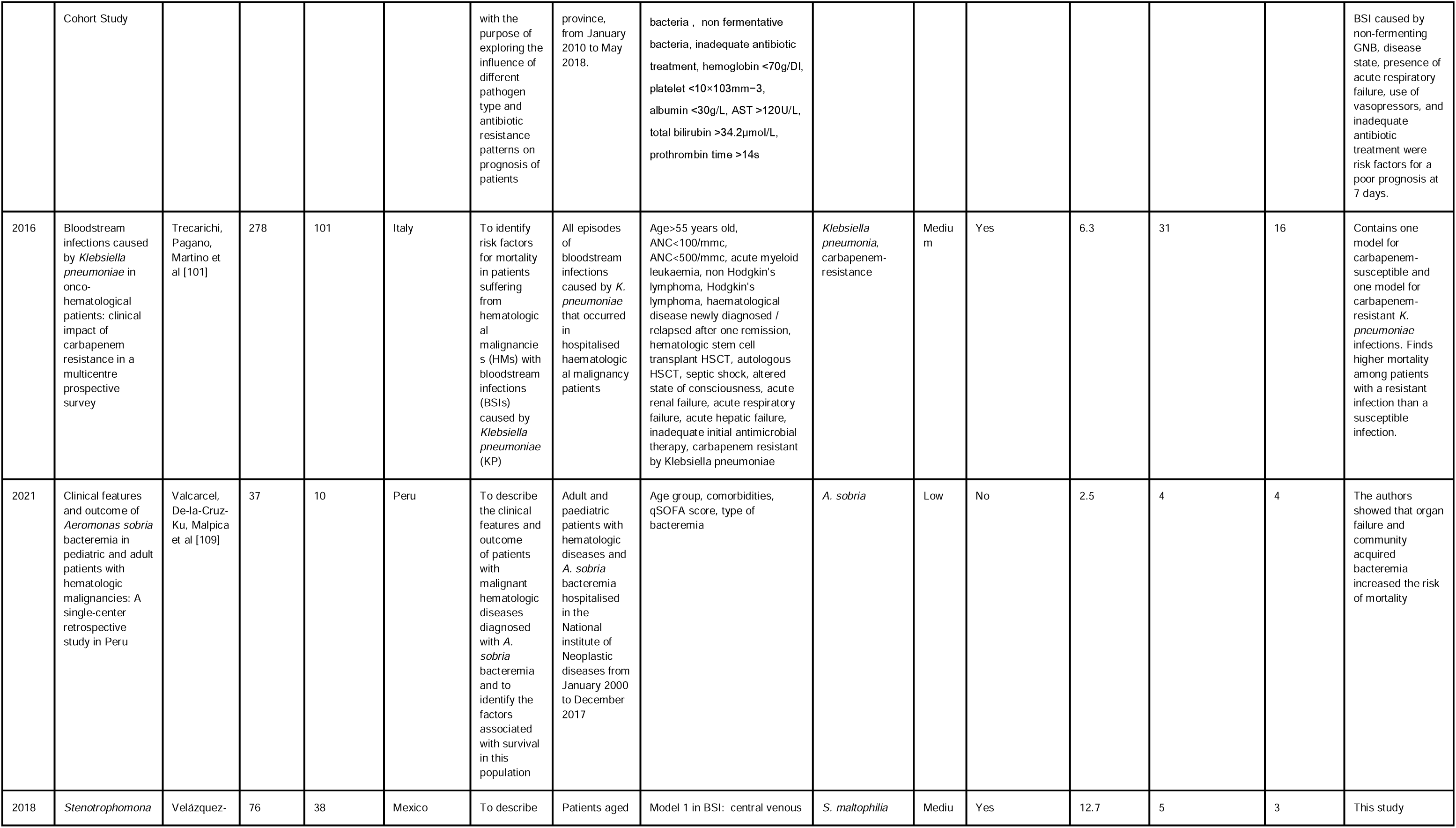
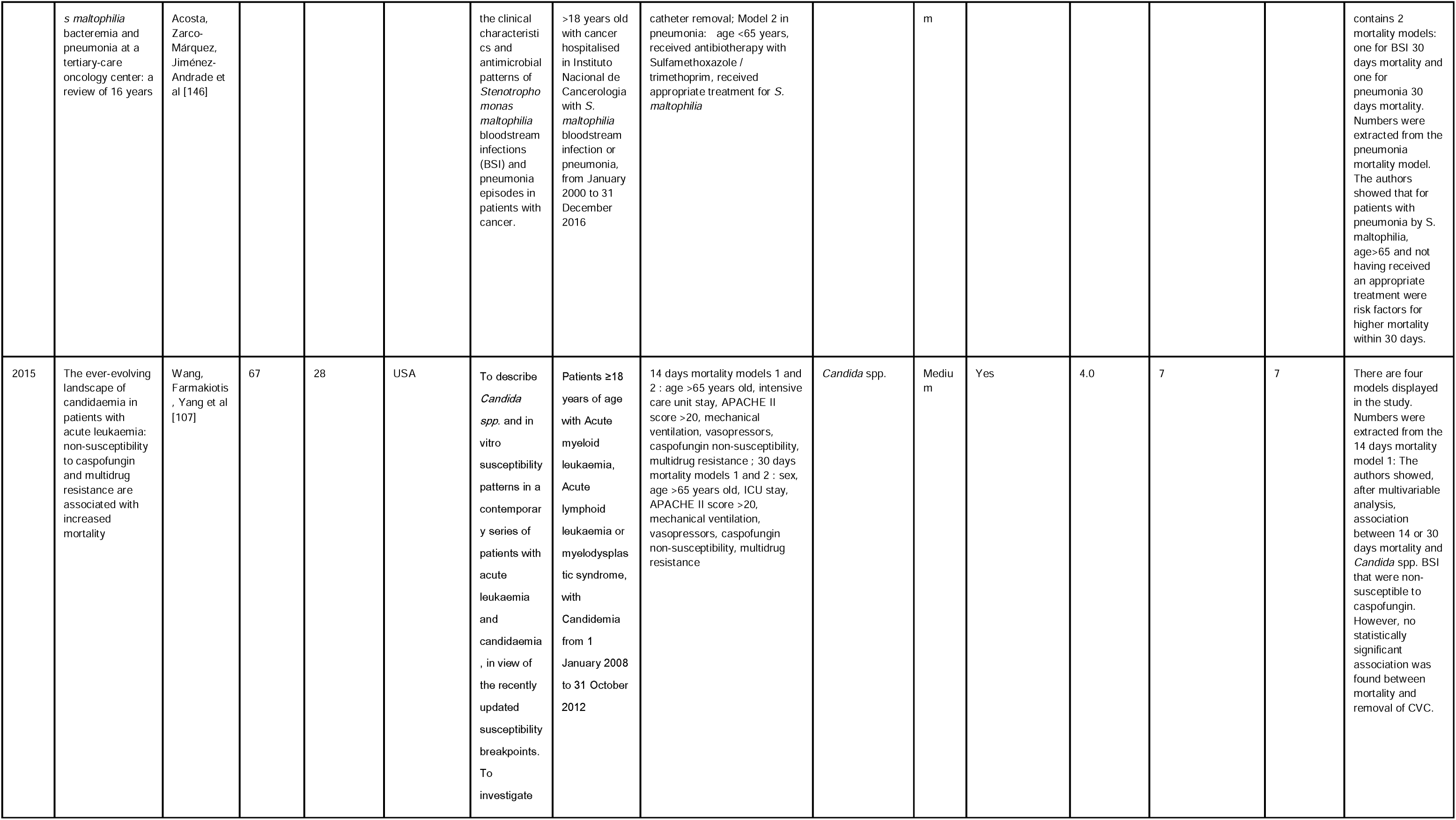
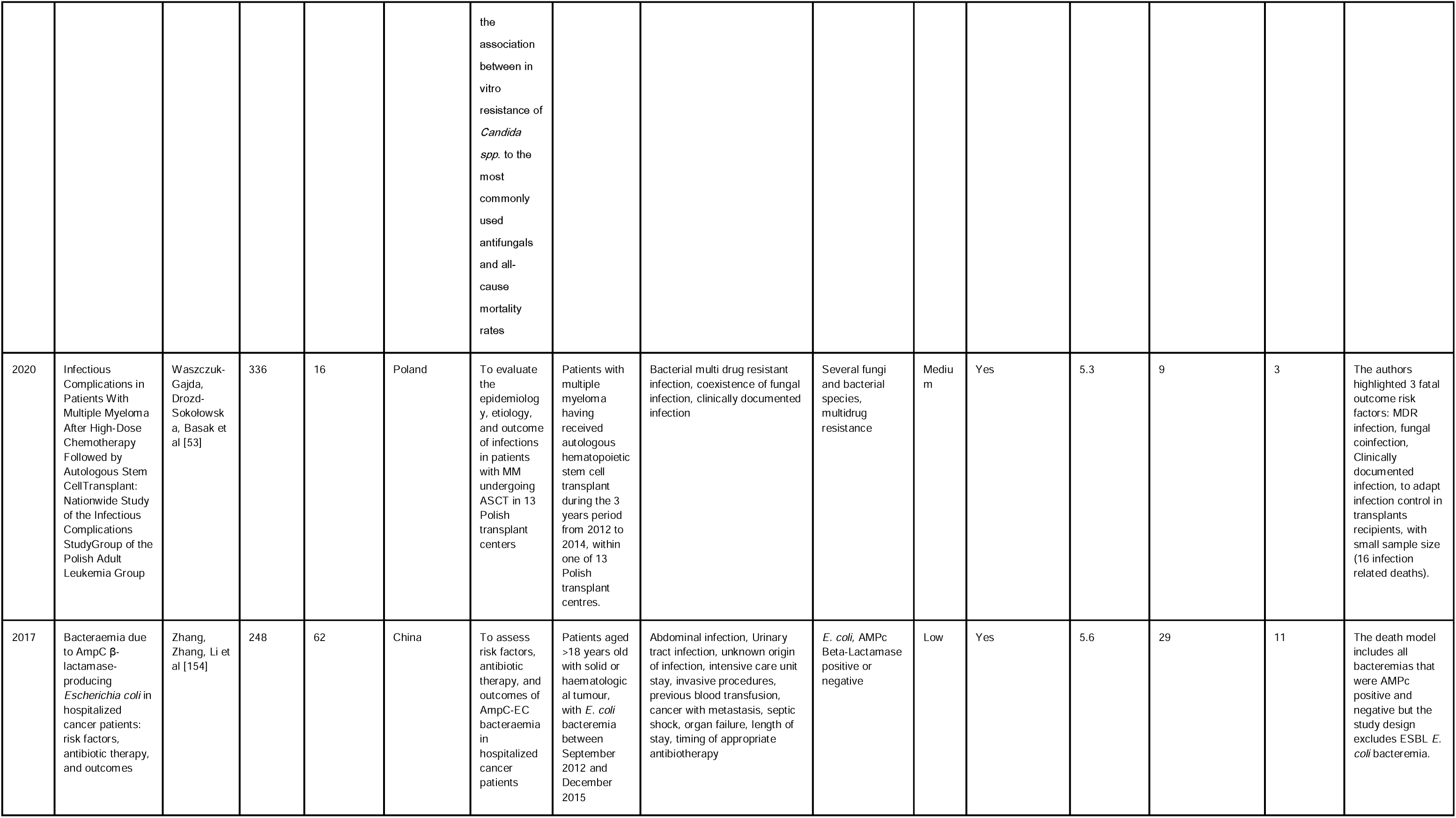
All included articles in the systematic review with a mortality outcome.

## Supplementary material 5

**Table S5.**
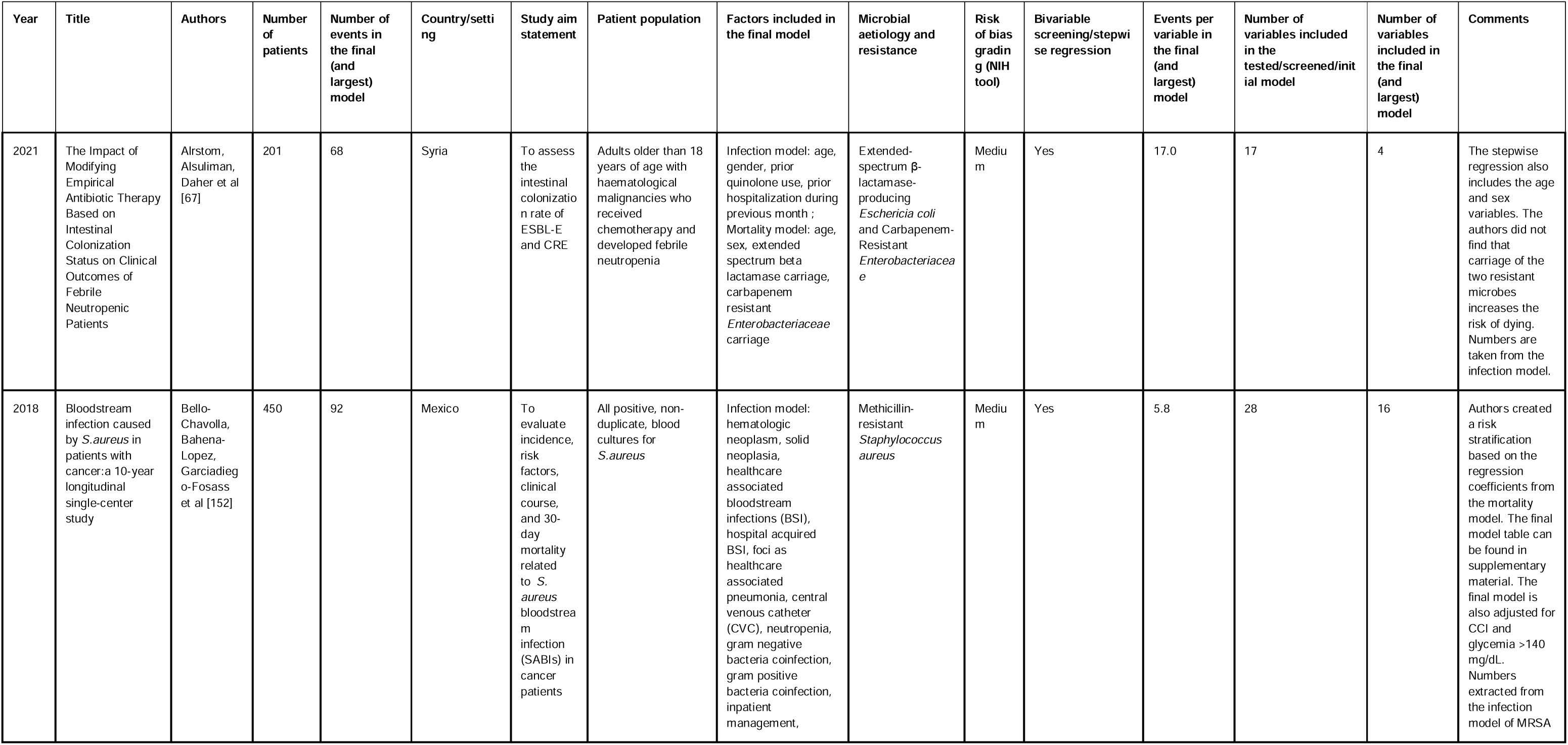
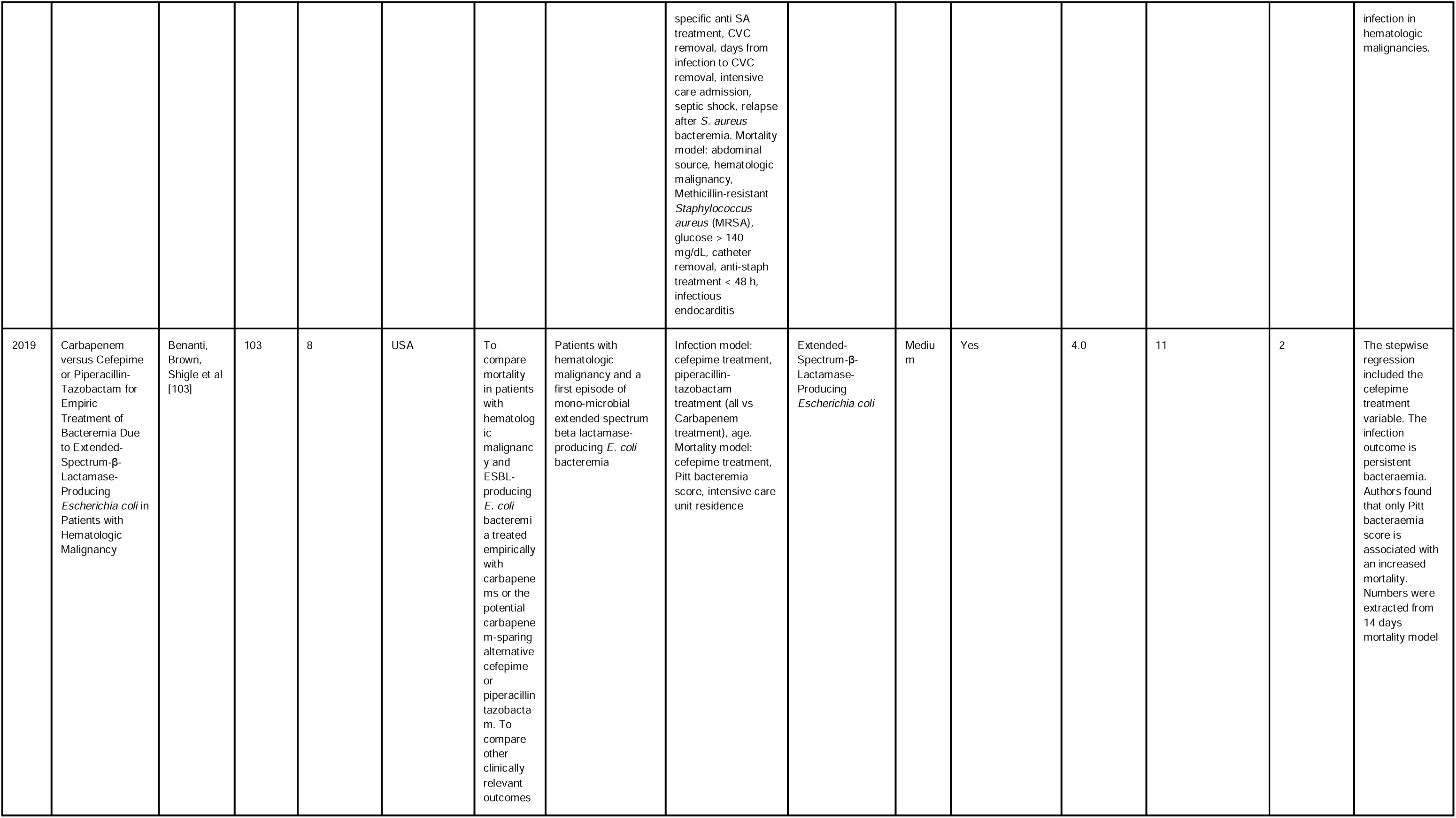
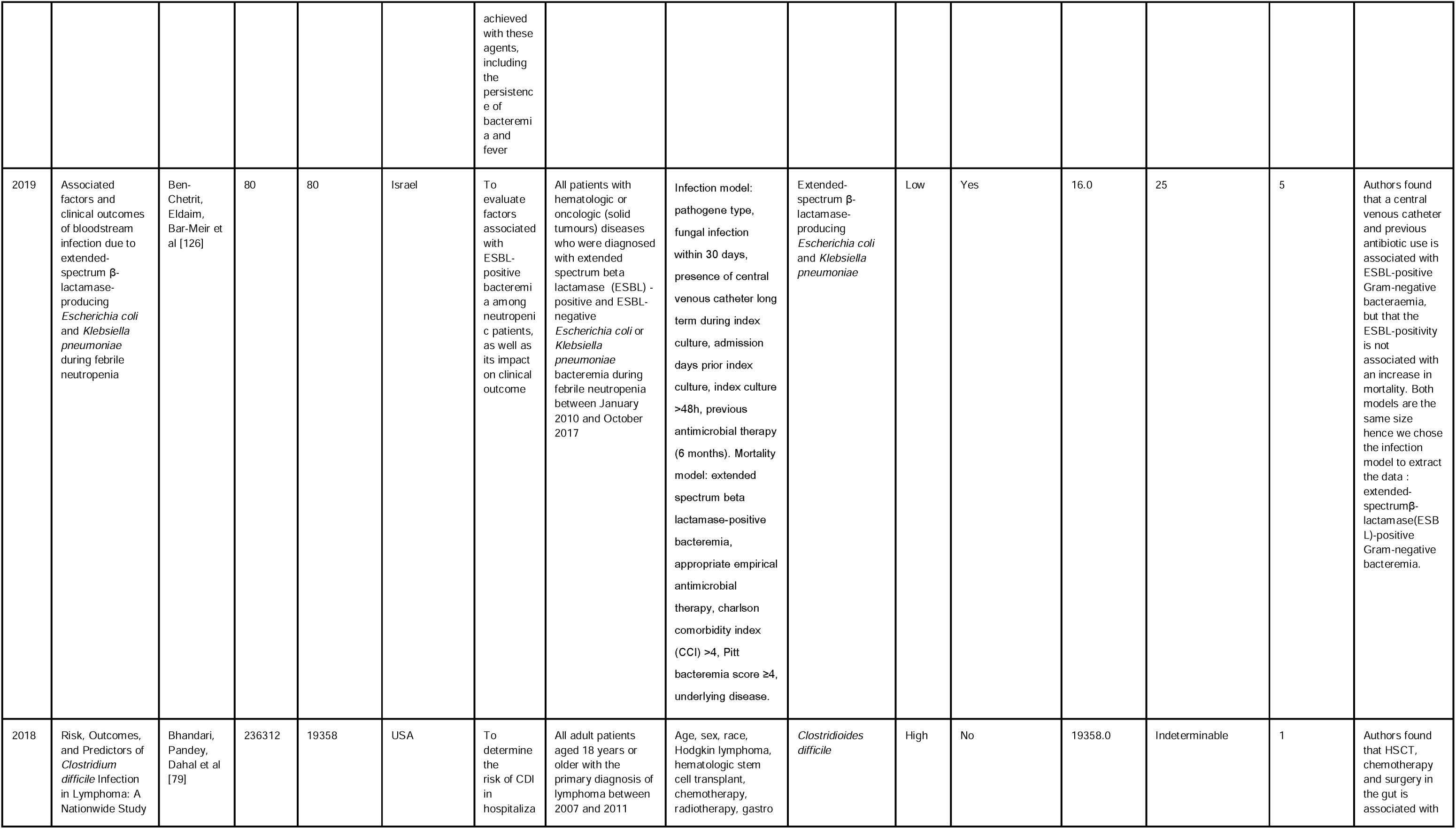
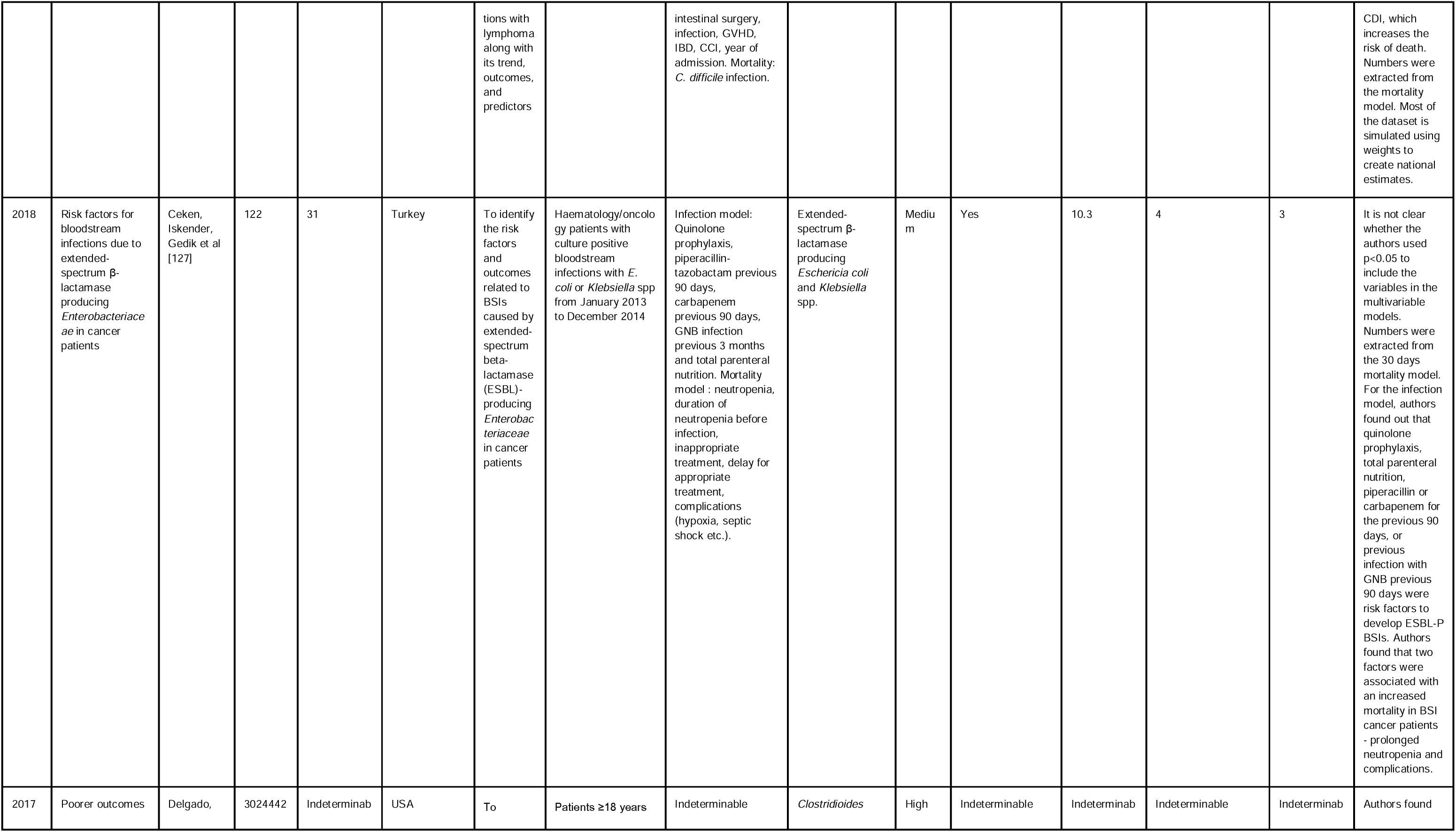
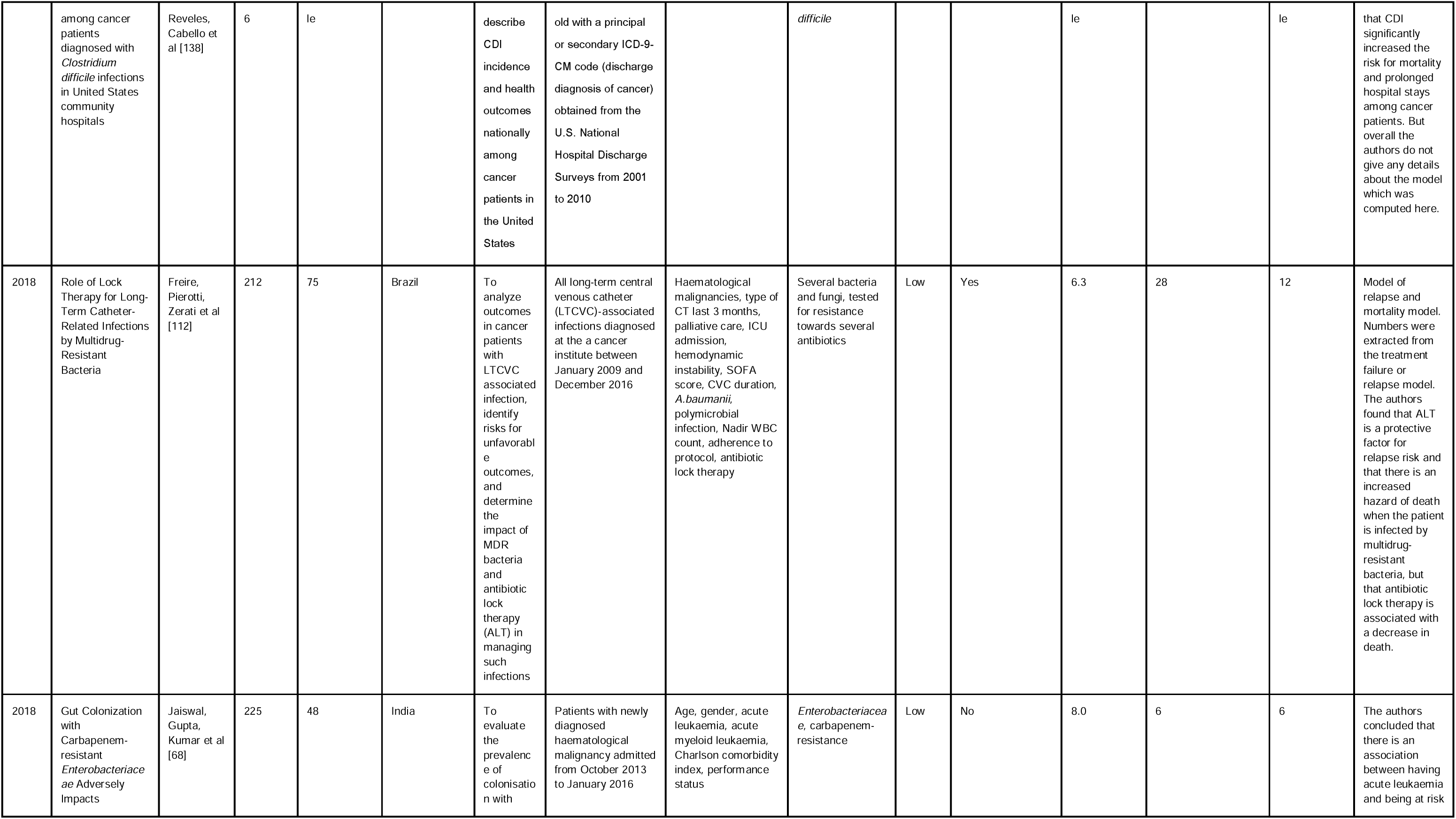
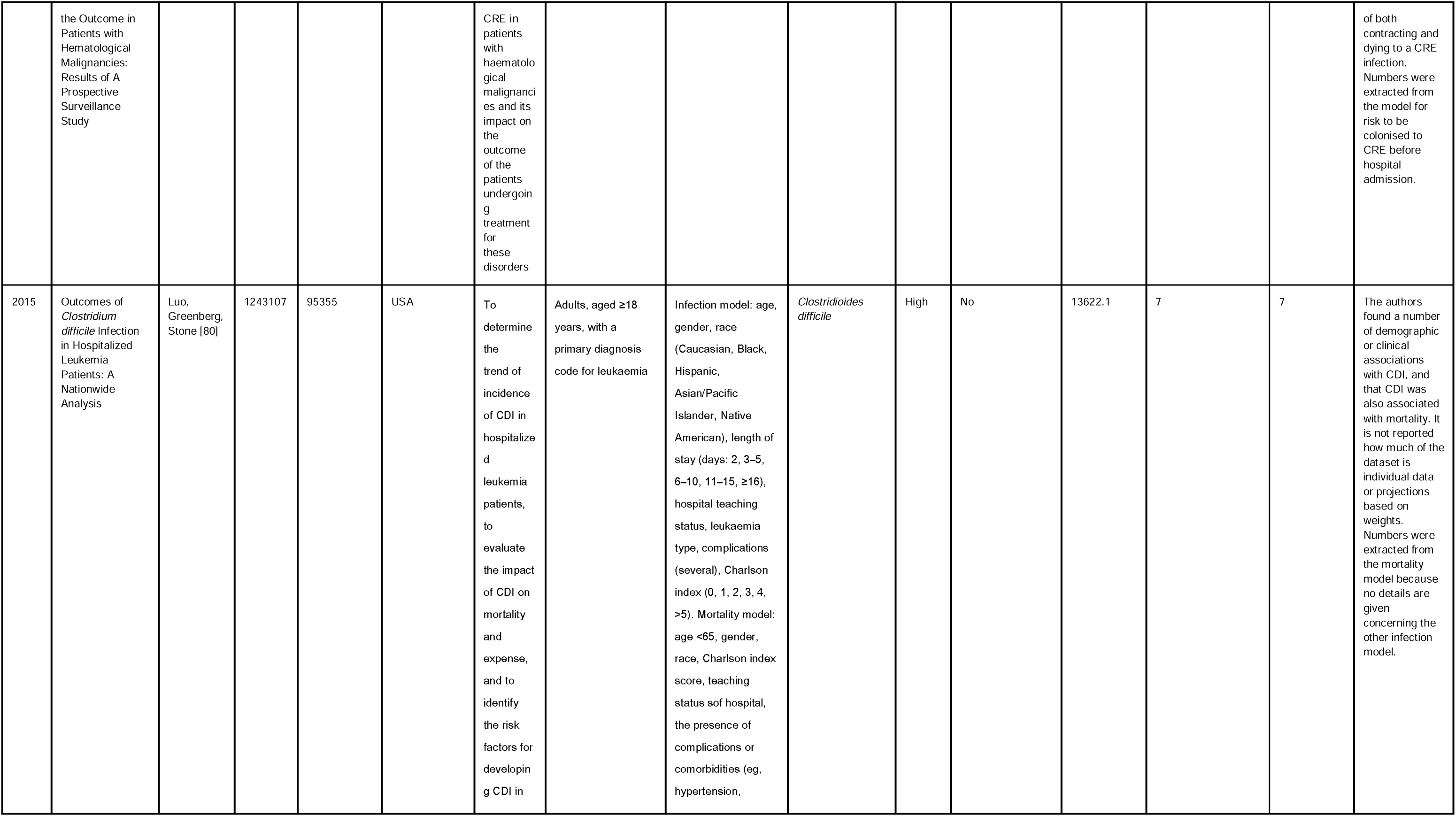
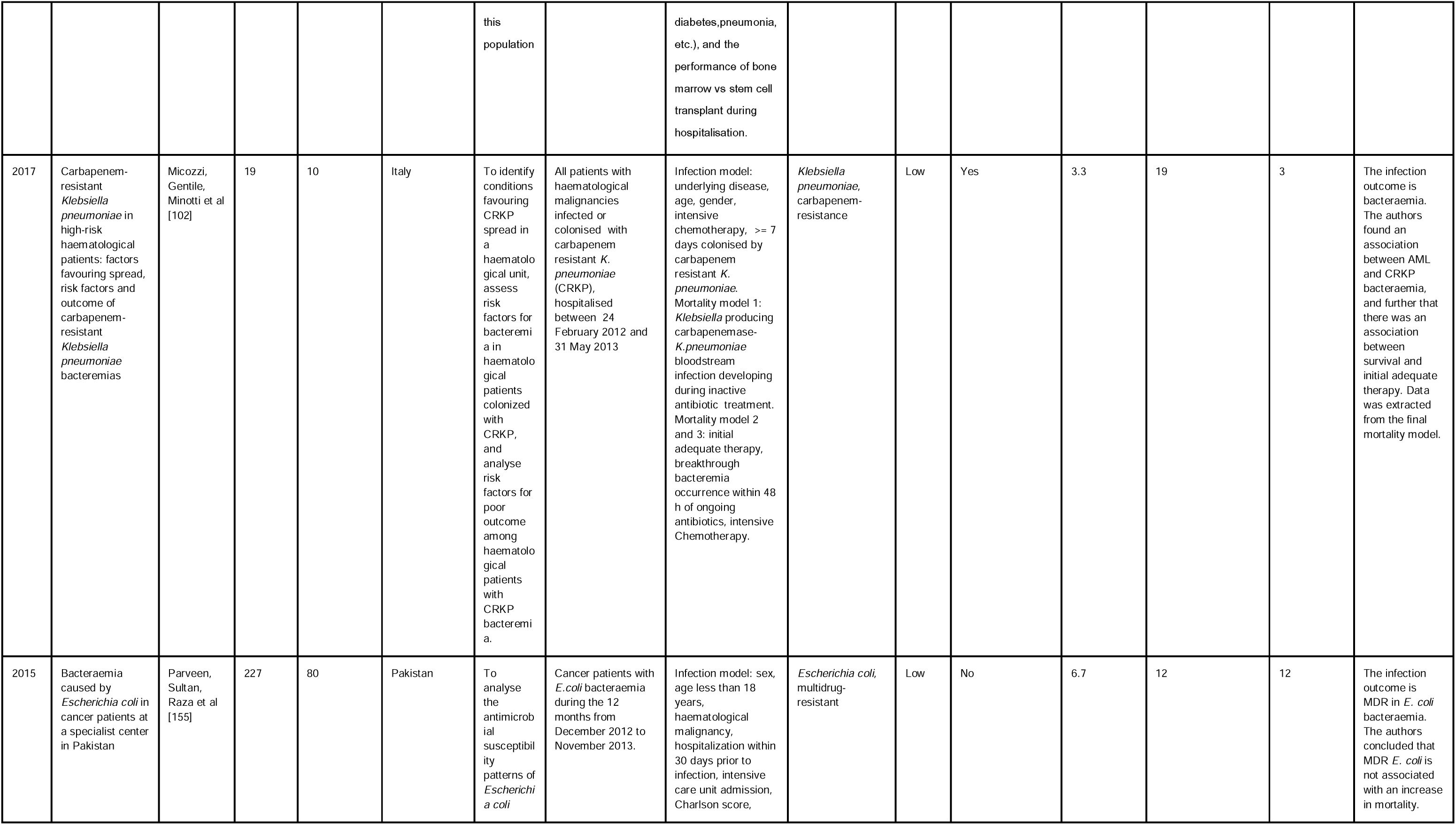
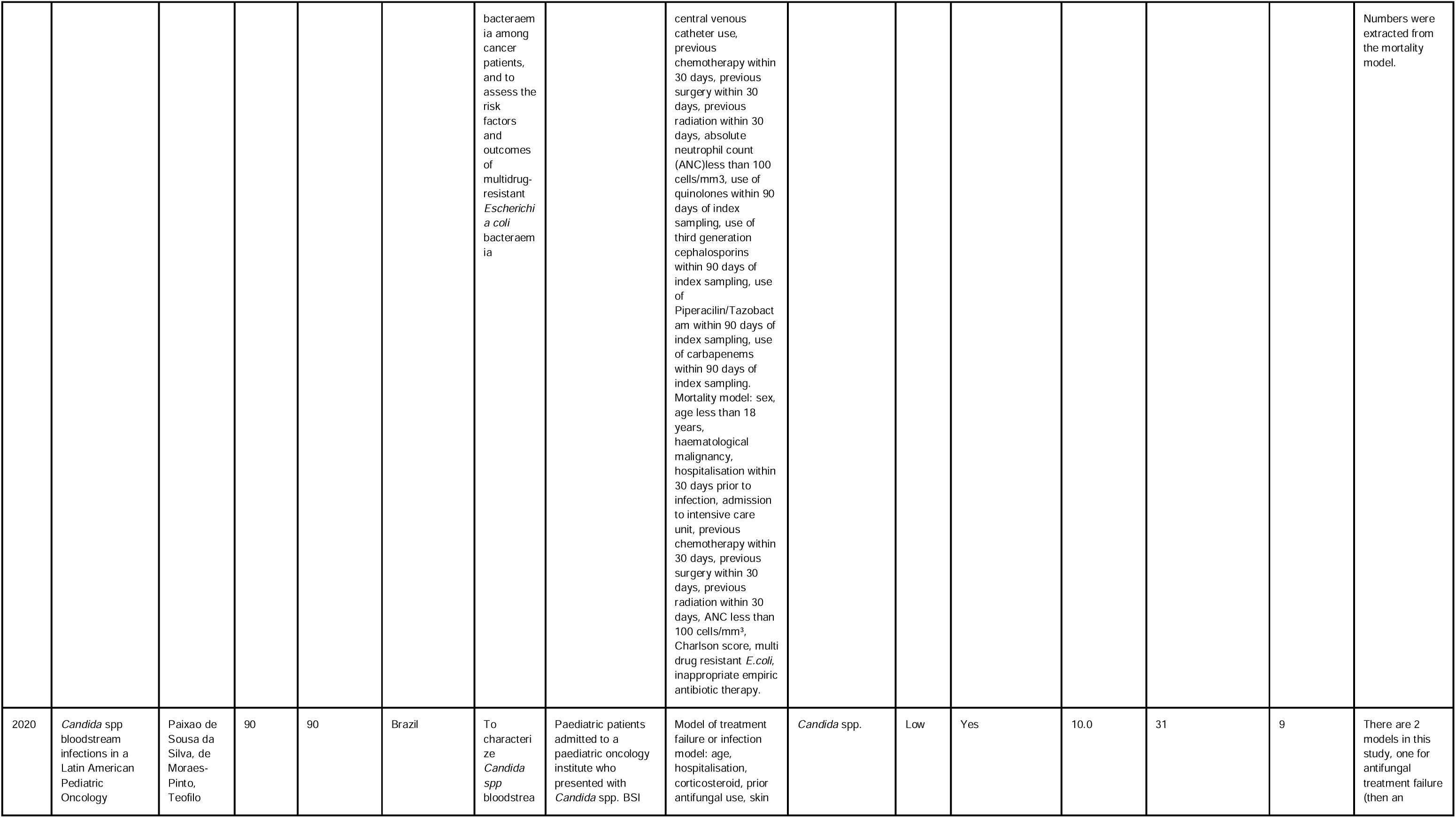
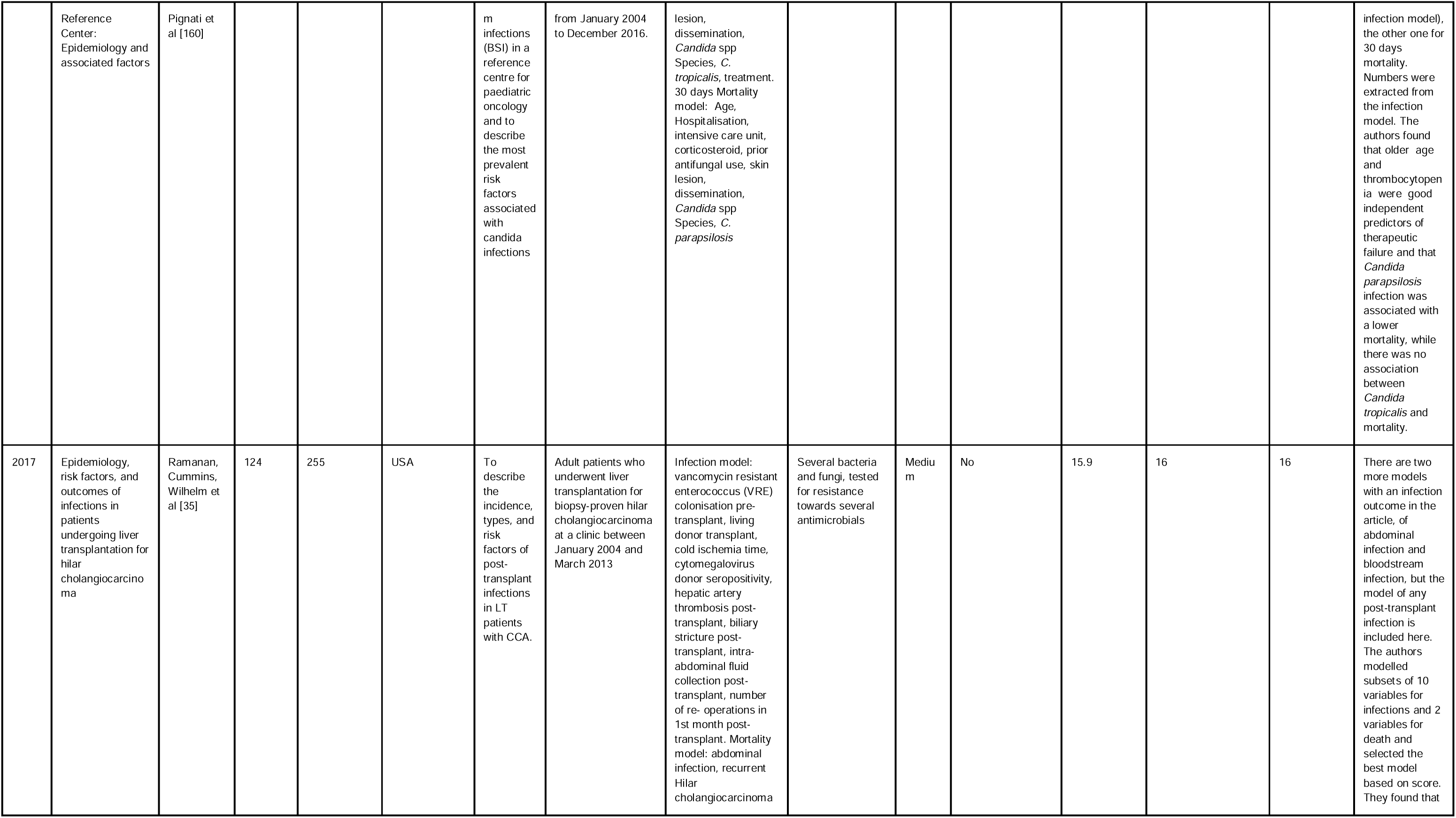
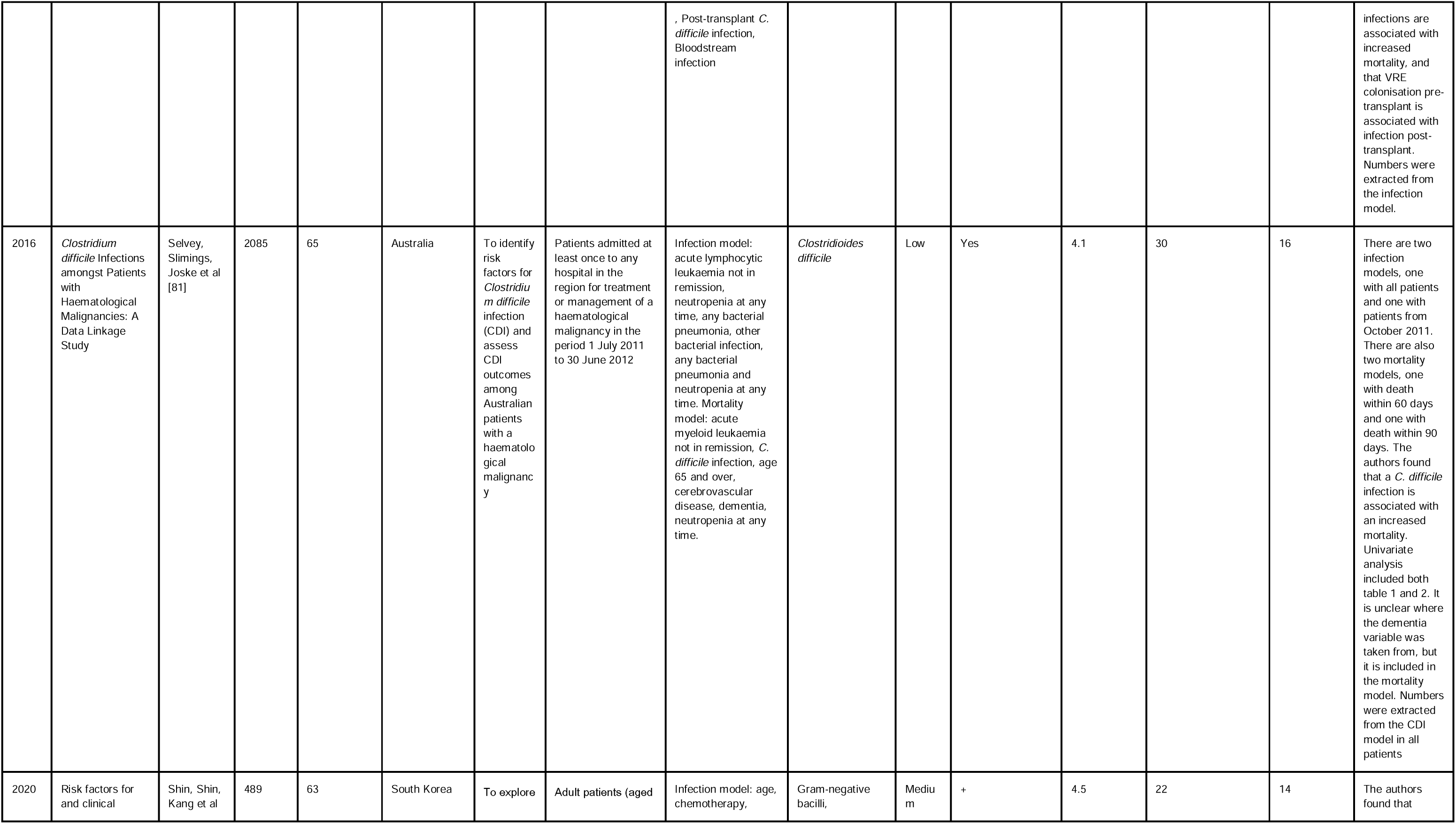
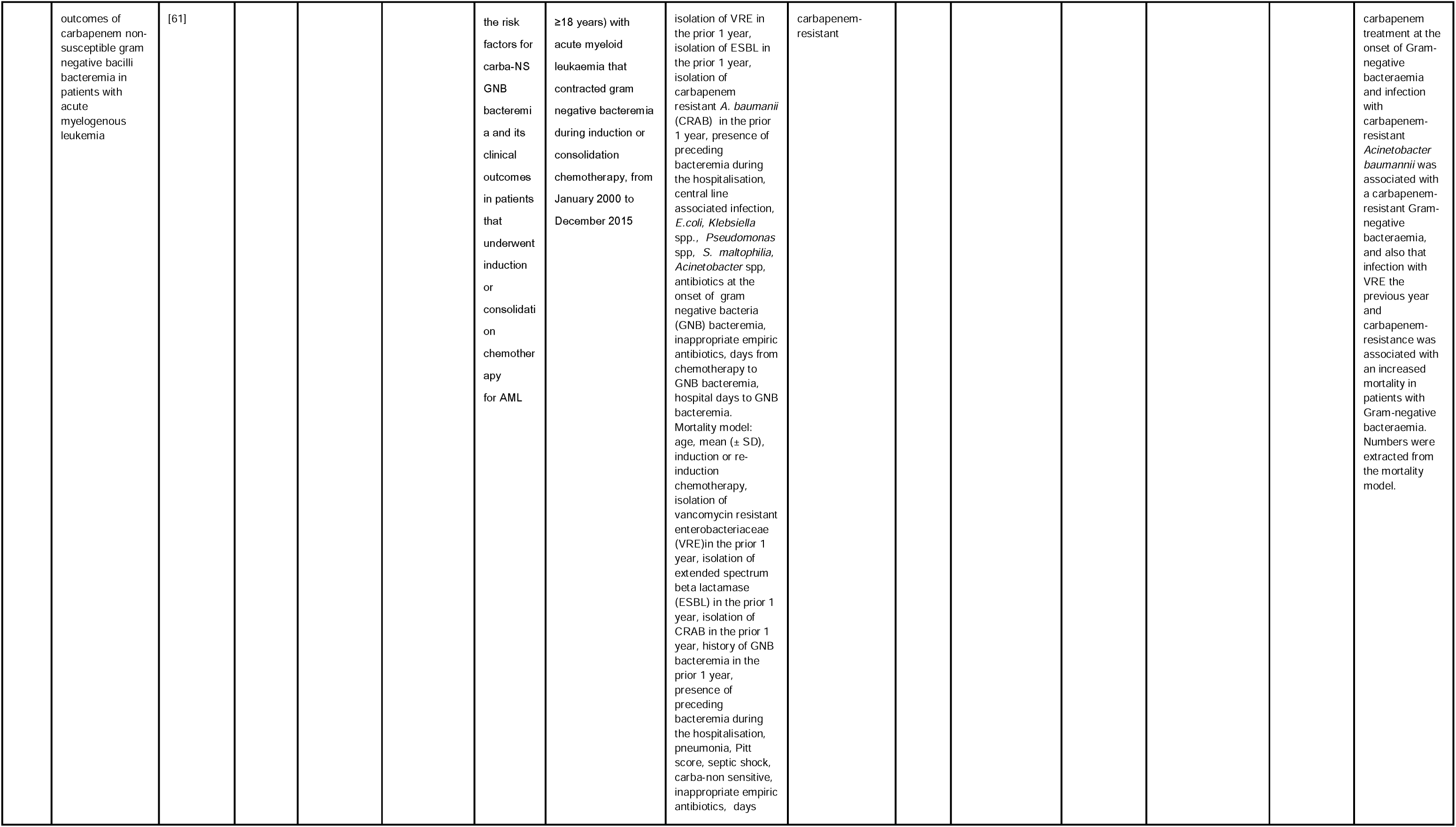
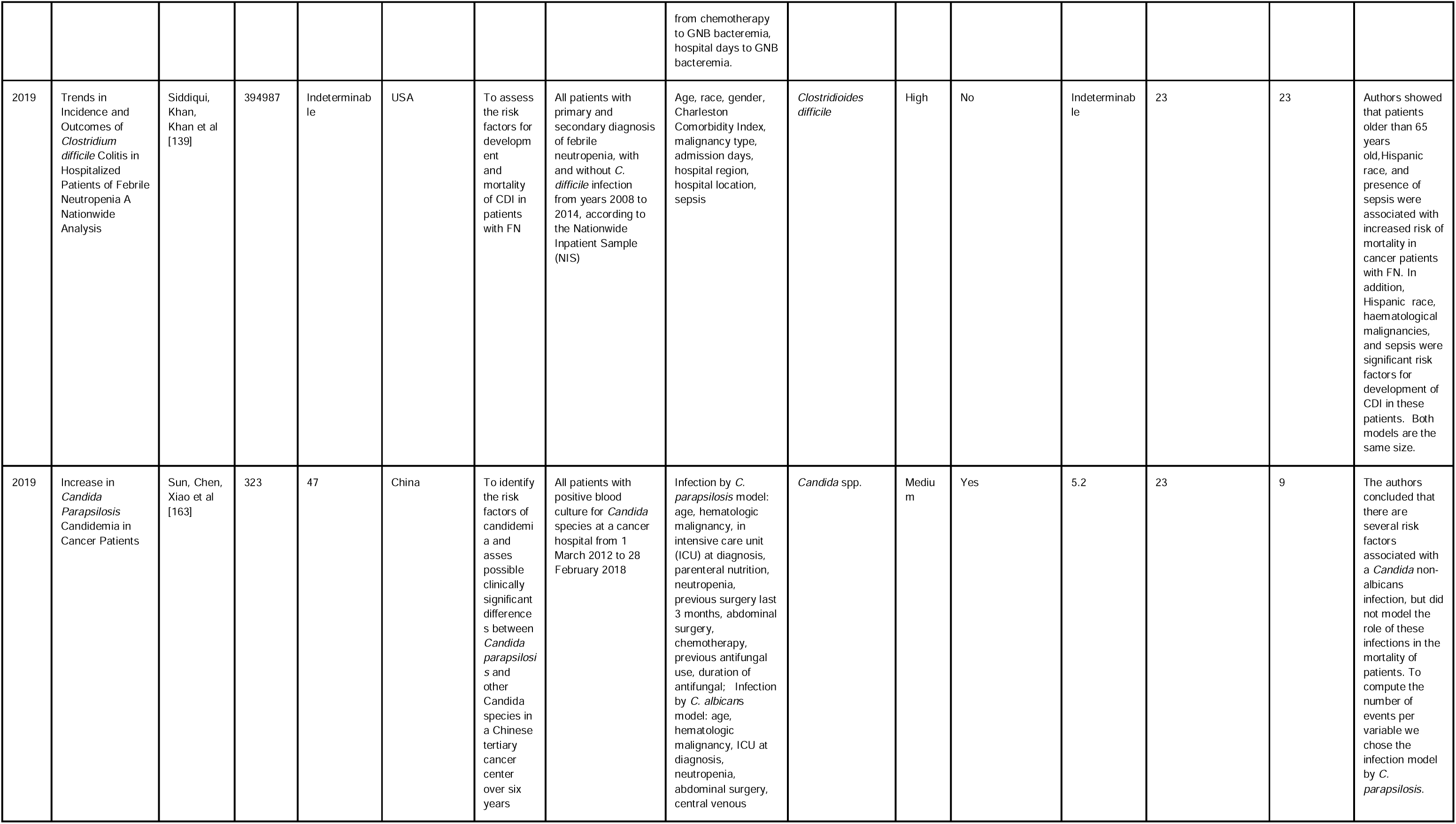
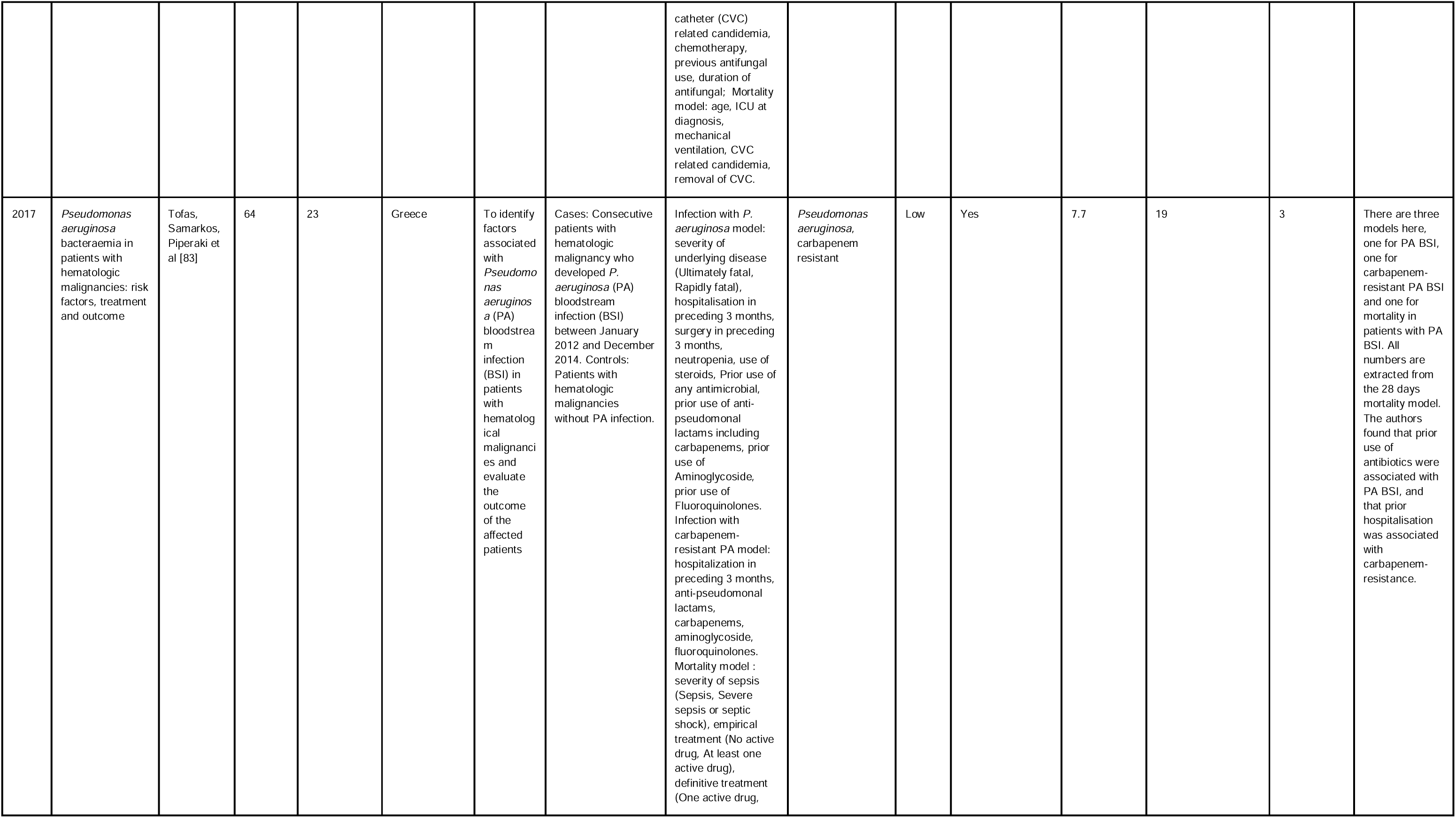
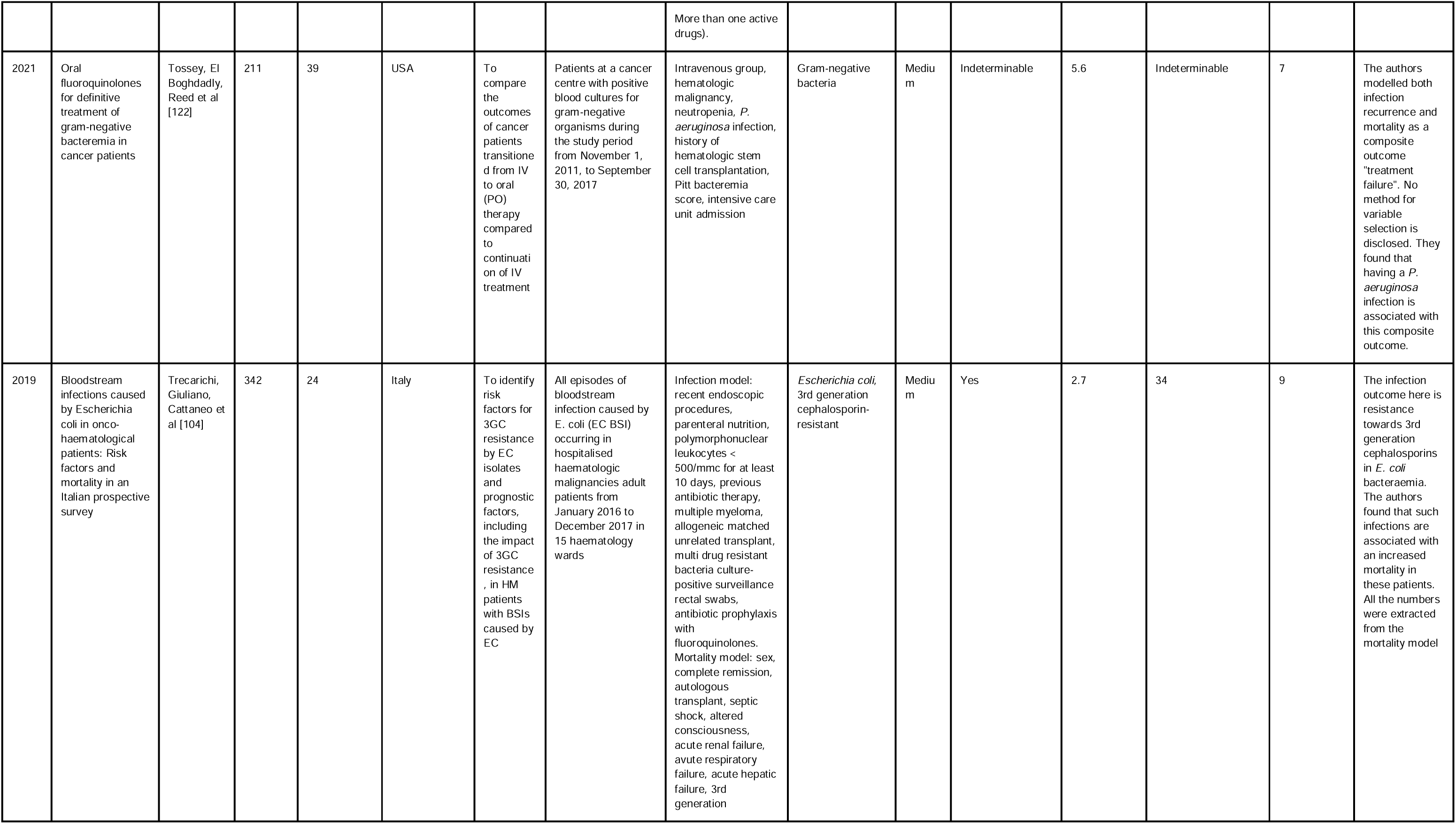
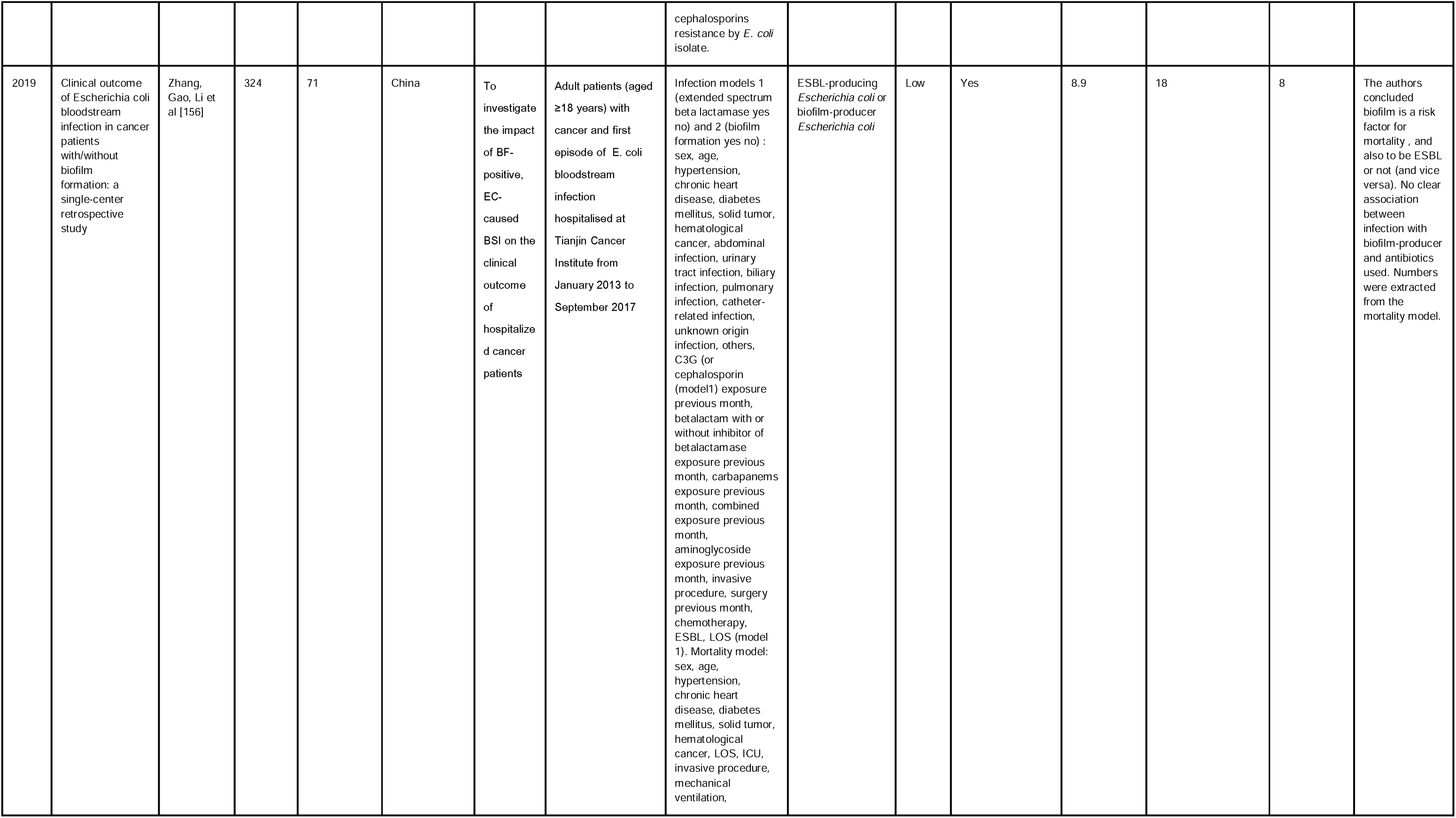
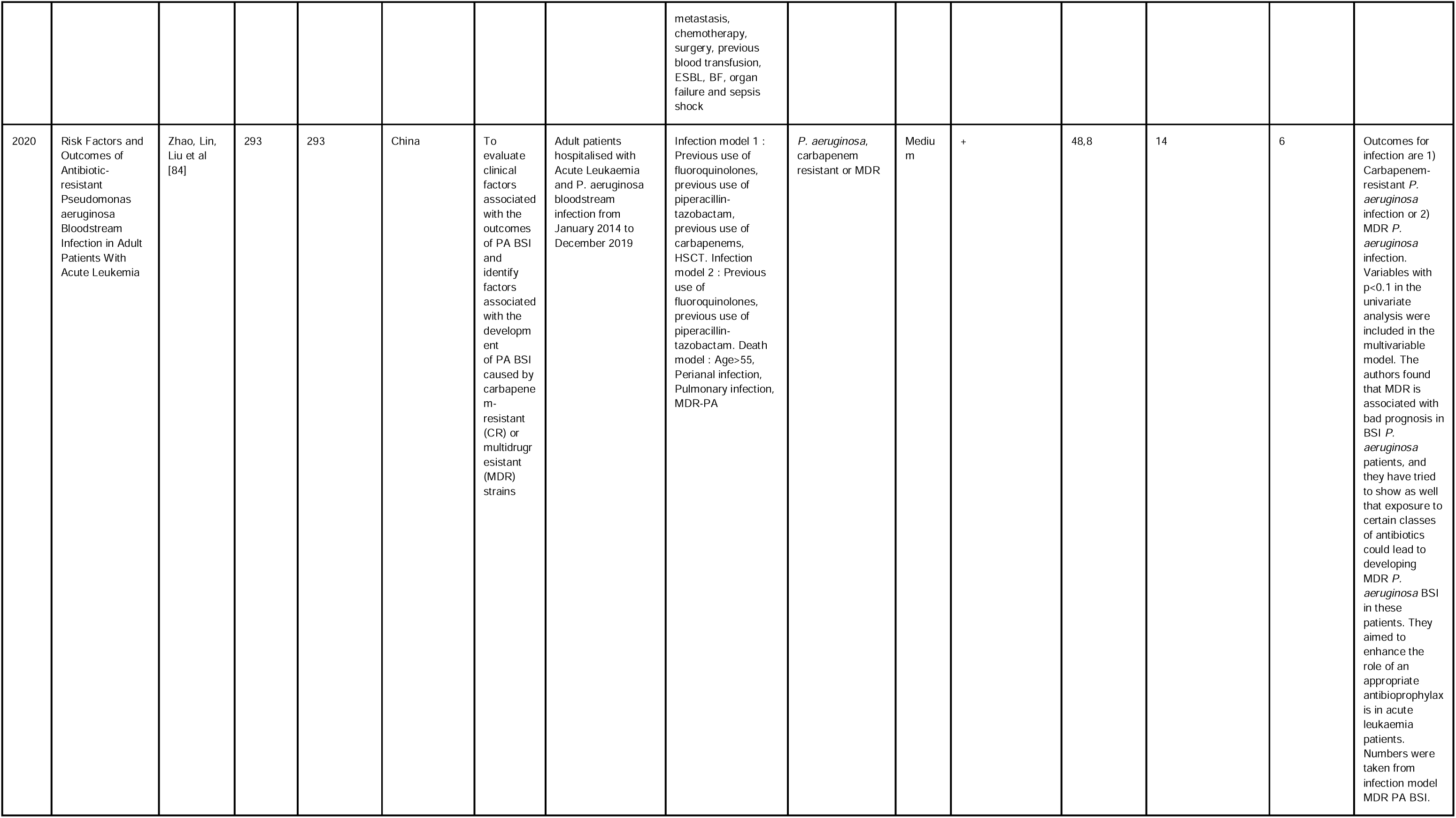
All included articles in the systematic review with both an infection/colonisation outcome and a mortality outcome.

## Supplementary material 6

**Table S6.** All risk factors investigated in the final, multivariable model of the included studies, and their respective categories.

| **Infection/colonisation** |  |
| --- | --- |
| **Category** | **Included risk factors** |
| Antibiotic use | Antibiotic use, Mean length of CDI treatment, CDI treatment failure, Ciprofloxacin, Levofloxacin, Meropenem, Antipseudomonal penicillin, Cumulative glycopeptide exposure, Susceptibility to applied perioperative antibiotics, β-lactam/β-lactamase inhibitor, Cephalosporin, Metronidazole, Previous antibiotic therapy, Prior antibiotic use, Days of empiric antibiotics, Cycling vs carbapenem period, Days of carbapenem, Days of cefepime or piperacillin/tazobactam, Antibiotics prior 3 mo, Antibiotics (no.), Antibiotics (d), Antibiotic therapy within 30 days, Prior piperacillin-tazobactam therapy (within 1 mo), Prior fluoroquinolone prophylaxis (within 1 mo), Prior antipseudomonal carbapenem therapy (within 1 mo), Antibiotic days of therapy, Vancomycin IV, Cephalosporin, Previous azole exposure history, Daptomycin, Cefepime received within 90 days, Cumulative Daptomycin days within 90 days, Cumulative Fluoroquinolone days within 90 days, Usage of quinolones inside 95 days of catalogue sample, Usage of third group cephalosporins inside 95 days of index sample, Usage of Piperacillin/Tazobactam within 95 days of index sampling, Usage of carbapenems inside 95 days of index sample, No. of previous antibiotic use $\geq 3$, Previous TMP/SMX use, Antibiotic prophylaxis (vs. G-CSF support), Prior use of extended-spectrum cephalosporin, Prior use of glycopeptides, Antibiotic use within 30 days before transplantation (No antibiotic, Any antibiotic except TMP-SMX, TMP-SMX), The total length of antibiotic administration for more than 5 days before diagnosis of anastomosis site leakage. Adjustment variables indeterminable., Days of antibiotics received, Antibiotic use previous 90 days (Antibiotic therapy >14 days), Specific antibiotics previous 90 days (PTZ, Vancomycin), Beta-lactams during the previous month, Oral vancomycin prophylaxis, prophylactic antibiotherapy received, treatment antibiotic duration, Antibiotic exposure, b-lactam/b-lactamase inhibitor within previous 30 days, Carbapenem within previous 30 days, Receiving TMP-SMX at BSI onset, Receiving cephalosporin at BSI onset, Receiving fluoroquinolone at BSI onset, Antibiotic exposure (ceftriaxone, fluoroquinolones, piperacillin/tazobactam, other (daptomycin), other (vancomycin IV), number of antibiotic exposures), Prophylactic antibiotic, IV antibiotics in previous 90 day, Concurrent antibiotics use for PCN infection, antifungals, Antibiotic exposure prior to CDI, non betalactam antibiotic previous exposure, Carbapenem previous exposure, empirical treatment of EB-BSI with carbapenems, empirical treatment with antibiotics combination, prior quinolone use, specific anti SA treatment, cefepime treatment, piperacillin-tazobactam treatment (all vs Carbapenem treatment), previous antimicrobial therapy (6 months)., Quinolone prophylaxis, piperacilline-tazobactam previous 90 days, carbapenem previous 90 days, use of quinolones within 90 days of index sampling, use of third generation cephalosporins within 90 days of index sampling, use of Piperacilin/Tazobactam within 90 days of index sampling, use of carbapenems within 90 days of index sampling., prior antifungal use, vancomycin resistant enterococcus (VRE) colonization pre-transplant, isolation of carbapenem resisant *A. baumannii* (CRAB) in the prior 1 year, antibiotics at the onset of gram negative bacteria (GNB) bacteremia, inappropriate empiric antibiotics, previous antifungal use, duration of antifungal, previous antifungal use, duration of antifungal, Prior use of any antimicrobial, prior use of anti-pseudomonal lactams including carbapenems, prior use of Aminoglycoside, prior use of Fluoroquinolones. Infection with carbapenem-resistant PA model: hospitalization in preceding 3 months, anti-pseudomonal lactams, carbapenems, aminoglycoside, fluoroquinolones, previous antibiotic therapy, antibiotic prophylaxis with fluoroquinolones, C3G (or cephalosporin (model1) exposure previous month, betalactam with or without inhibitor of betalactamase exposure previous month, carbapenems exposure previous month, combined exposure previous month, aminoglycoside exposure previous month, previous use of fluoroquinolones, previous use of piperacilline- tazobactam, previous use of carbapenems, Previous use of fluoroquinolones, previous use of cephalosporines, previous use of piperacilline- tazobactam, previous use of carbapenems |
| Basic characteristics | Age, BMI, Male sex, Age, Age, Age, Male (vs. female), Gender, Age, Age, Age, Age, Gender female, ESBL-PE. Adjusted for the following confounders: age, sex, Age ($\geq 60$ years vs <60), Gender (male), Asian race, Man sex, Age fewer than 19 years, Age (per ten years), Hospitalization during previous year, Gender, Race, Age, Year, Age categories, Race/Ethnicity, Female, Obesity, Median household income in quartile., Age at |
|  | diagnosis, Age (>65), Smoking history (+ vs -), Sex, Age (<60 vs >60), BMI, age, gender, age., Age, sex, race, year of admission., Age, gender, age, gender, race, age, gender, sex, age less than 18 years, age, age, Age, race, gender, age, age, sex, age, sex |
| Cancer-related | Hematological malignancy, Tumour stage, Tumour localisation, Hematologic malignancy, AML, Karnofsky performance Status, and type of malignancy., HCl-CI ($\geq 3$ vs 0-2), KPS (>80 vs $\leq 80$), Stage of malignancy, Hematologic malignancy, Hematology/oncology model: Stage of disease (Hematologic malignancy–newly diagnosed, Hematologic malignancy–continuation/consolidation), Neoplasia in remission, Type of Malignancy, Gastrointestinal cancer, Breast cancer, Lung cancer, Prior CRE at any site within the previous 90 days, salvage lymphoma, salvage lymphoma), Leukemia, Lymphoma, Site (Colon vs Rectum), Morphology (Ulcerative vs Exophytic), Differentiation (Poor vs Well), T stage (Non-T4 vs T4), N stage (LNM neg. Vs LN pos.), hematological neoplasm, solid neoplasia, Hodgkin lymphoma, acute leukemia, acute myeloid leukemia, leukemia type, haematological malignancy, acute lymphocytic leukaemia not in remission, malignancy type, hematologic malignancy, hematologic malignancy, severity of underlying disease (Ultimately fatal, Rapidly fatal), hematologic malignancy, multiple myeloma, solid tumor, hematological cancer |
| Chemotherapy/immunosuppressants | Immunosuppressant use, Conditioning regimen, Chemotherapy dose modifications, Intervall chemotherapy to diarrhea, Administration of chemotherapy, Conditioning (RIC vs MAC), Number induction chemotherapy regimens, Clofarabine within 30 days, Previous chemotherapy inside 1 month, No prophylaxis (vs. G-CSF support), ≥VGPR before ASCT (vs. ≤PR), Oncology treatment previous 90 days (Rituximab, Clofarabine)., Immunosuppressive therapy, No chemotherapy, Early de-escalation vs hematopoietic recovery de-escalation, chemotherapy exposure, decitabine, CHOP, Chemotherapy delay OR and mean difference., chemotherapy, intensive chemotherapy, previous chemotherapy within 30 days, chemotherapy, chemotherapy, chemotherapy, chemotherapy |
| Comorbidities/clinical condition | Comorbidities, Diabetes mellitus, Pleural effusion, Hematologic diseases, Diabetes, Hematological disease, Congestive heart failure, Haematological distortion, Carlson score, Diabetes mellitus, Peripheral vascular disease, Chronic renal disease, Atrial fibrillation, Congestive heart failure, Hypertension, Chronic liver failure, Elixhauser score, Renal disease, Chronic renal diseases, Gastroenteritis diseases, CCI, Charlson comorbidity index, Charlson index, underlying disease, Charlson score, cold ischemia time, Charleston Co-Morbidity Index, severity of underlying disease (Ultimately fatal, hypertension, chronic heart disease, diabetes mellitus) |
| Gastrointestinal | Gastrointestinal GVHD, Number of stools/day, Preoperative biliary stenting (yes vs. no), Gastrointestinal bleeding, Diarrhea at presentation, Inflammatory bowel disease, IBD, biliary stricture post- transplant, biliary infection |
| Hospital-related | Previous hospitalization, Same ward, Length of ICU stay >3 days, Hospitalization time, LOS, Hospitalization prior 2 mo, Hospital LOS prior to positive culture, Hospitalization inside 1 month before contagion, ICU admission, Hospital stay of longer than 14 days, Demographic model: Unit ICU, LOS >14 days prior to culture, Healthcare exposures previous 90 days (ICU admission), Transfer from outside hospital, Prolonged hospitalization ($\geq 7$ days), Hospital bed size, Hospital teaching status, Expected primary payer, Total prior Hospital Stay, Total Prior Admission in ICU, ICU stay within previous 30 days, Longitudinal model: Hospitalized (days), LOS prior to culture, Model 1 Risk for CDI: Length of stay before CDI, Model 2 outcomes linked to CDI : LOS, Length of hospitalization, Excessive bed rest, prior hospitalization during previous month, inpatient management, intensive care admission, admission days prior index culture, length of stay, hospital teaching status, hospitalization within 30 days prior to infection, intensive care unit admission, hospitalization, presence of preceding bacteremia during the hospitalization, admission days, hospital region, hospital location, in intensive care unit (ICU) at diagnosis, ICU at diagnosis, hospitalization in preceding 3 months, intensive care unit admission, LOS (model 1), previous hospital admission within 60 days, duration of hospital stay |
| Infection-related | Severe/fulminant CDI disease, S. maltophilia oral abundance, Previous bacteremia with another pathogen, Bacterial infection post- UCBT, Hospital-acquired pneumonia, Development of pneumonia within the first 10 days, Prior history of Clostridioides difficile, Healthcare-associated infection, Linezolid-resistant S. epidermidis, VRE colonization (within 30 days of HCT), Culture specimen–BAL fluid, Prior enterococcal infection, Polymicrobial infection, Previous S. maltophilia isolation, Breakthrough infection during carbapenem therapy, CMV reactivation, Herpesviridae reactivation other than |
|  | CMV, culture from sputum specimens, Source of positive culture (Respiratory), Resistance model: History of PTZ-R, Outpatient CDI diagnosis., Type of Organism, Infection, Demographical and traditional model: Severe CDI, PCN exchange within 4 days of infection, CDI exposure, presence of BSBL-Resistant Enterobacteria, ESBL-EB, and non E. coli EB in blood cultures, CRKP rectal colonization, Fever days, healthcare associated bloodstream infections (BSI), hospital acquired BSI, foci as healthcare associated pneumonia, gram negative bacteria coinfection, gram positive bacteria coinfection, septic shock, relapse after S. aureus bacteremia., pathogene type, fungal infection within 30 days, index culture >48h, infection, GNB infection previous 3 months and total parenteral nutrition., >= 7 days colonized by carbapenem resistant K. pneumoniae, Candida spp Species, C. tropicalis, any bacterial pneumonia, other bacterial infection, isolation of VRE in the prior 1 year, isolation of ESBL in the prior 1 year, central line associated infection, E.coli, Klebsiella spp., Pseudomonas spp, S. maltophilia, Acinetobacter spp, days from chemotherapy to GNB bacteremia, hospital days to GNB bacteremia, P. aeruginosa infection, Pitt bacteremia score, multi drug resistant bacteria culture-positive surveillance rectal swabs, abdominal infection, pulmonary infection, catheter-related infection, unknown origin infection, ESBL |
| Laboratory findings (non-microbiological) | Histology, Admit serum albumin, albumin, Preoperative hemoglobin, Albumin (<35 vs >35), FBG (>7 vs <7), Triglyceride, Total cholesterol, HDL, Hemoglobin, OB, Hypoproteinemia, Procalcitonin, C Reactive Protein, Serum Amyloid, polymorphonuclear leukocytes < 500/mm<sup>3</sup> for at least 10 days |
| Neutropenia | Severe neutropenia (<100/mm<sup>3</sup>), Febrile neutropenia, Duration of neutropenia, Length of neutropenia ≥3 days, Severe neutropenia, Severe neutropenia, Typhilitis, ANC less than 110 cells/mm<sup>3</sup>, Profound neutropenia, Neutropenia, neutropenic days, Neutropenia, Haematological risk factors (neutropenia (days), Death during neutropenia, Severe neutropenia, neutropenia, absolute neutrophil count (ANC) less than 100 cells/mm<sup>3</sup>, neutropenia at any time, any bacterial pneumonia and neutropenia at any time., neutropenia, neutropenia, neutropenia, neutropenia |
| Other | Preceding fallout inside 1 month, Stem cell amount ≥2.5 * (vs. <2.5), Mucositis, [Indeterminable], mucositis, Pancreatic texture (soft vs hard), denture, objective dry mouth scores, NACS (?), Level 2 of patient's I tensity of Treatment Rating Scale, IMV., Indeterminable, performance status, complications (several), skin lesion, dissemination, treatment. 30 days, sepsis, recent endoscopic procedures, others" |
| Other treatments/medications | NSAID use, PPI use, Radiation, Proton pump inhibitors, Preoperative antacid use, Significant use of corticosteroids, proton pump inhibitor, Corticosteroid use, PP inhibitor use, Receiving glucocorticoids at BSI onset, Hemodialysis in previous 30 day, radiotherapy, previous radiation within 30 days, corticosteroid, use of steroids |
| Respiratory | COPD, Respiratory failure, Prior Mechanical Ventilation |
| Surgery-related | Surgical treatment, Portal vein embolization, Operation, Type of operation (pancreatoduodenectomy vs. others), Operation time, Estimated blood loss, Gastro-intestinal/abdominal surgery, Previous surgery inside 1 month, Invasive devices at the time of culture, Previous surgery, Invasive operation, gastro intestinal surgery, previous surgery within 30 days, previous surgery last 3 months, abdominal surgery, abdominal surgery, surgery in preceding 3 months, invasive procedure, surgery previous month |
| Transplantation-related | Organ transplantation, Prior HSCT, ASCT at relapse (vs. first-line treatment), History of HSCT, Stem cell transplantation, Autologous transplant, Allogeneic transplant, level 3 with HCST, level 3 without HCST and the HCST status, hematologic stem cell transplant, GVHD, living donor transplant, cytomegalovirus donor seropositivity, hepatic artery thrombosis post- transplant, intra-abdominal fluid collection post-transplant, number of re-operations in 1st month post- transplant., history of hemotologic stem cell transplantation, allogeneic matched unrelated transplant, hematologic stem cell transplantation |
| Urinary | Urinary catheter, Diversion type, Urinary catheter at culture, Prior Urinary Catheterization, urinary tract infection |
| Intravascular access | Blood transfusion, Presence of CVC, Dominant intravenous tube use, Presence of a CVC, Peripheral or Central CVP Line, central venous catheter (CVC), CVC removal, days from infection to CVC removal, presence of central venous catheter long term during index culture, GNB infection previous 3 months and total parenteral nutrition., central venous catheter use, parenteral nutrition, central venous catheter (CVC) related candidemia, Intravenous group, parenteral nutrition |
| **Mortality** |  |
| **Category** | **Included risk factors** |
| Antibiotic use | received vancomycin within 4 weeks prior to bacteraemia, meropenem exposition (yes/no), appropriate empirical antibiotic treatment, inadequate empirical antibiotic therapy, inadequate initial antibiotic treatment, imipenem empirical antibiotic regimen, bacterial growth during antibiotic treatment, |
| | administration of appropriate antibiotic treatment for minimum 72 hours, Use of antibiotics ($\geq 3$ agents), appropriate treatment/Non-appropriate treatment, Antimicrobial prophylaxis, initial antimicrobial failure, echinocandin pre-exposure, appropriate treatment within 48 h after blood culture collection, 3rd and 4th generation cephalosporines, fluoroquinolones., received appropriate therapy for XDR-ABC bacteraemia, inappropriate antibiotic therapy for multi drug resistant strains, inappropriate antibiotic therapy for multi drug resistant *P. aeruginosa*, Appropriate treatment vs Inappropriate treatment, previous antibiotics <3, early empirical TMP/SMX antibiotic use within 72 h, exposure to empirical antibiotics, azole monotherapy, Empirical antibiotic combination therapy, Inappropriate empirical antibiotic therapy, antibiotic treatment for the last 3 months before admission, appropriate antibiotic, empirical antifungal, antibiotic prophylaxis, carbapenem empirical therapy, vancomycin empirical therapy, Model 3: guided antibiotics, duration of guided antibiotics, Model 4: empirical antibiotics, guided antibiotics, guided antibiotics, Model 6: duration of empirical antibiotics, Model 7: guided antibiotics, effective empirical antibiotics, inappropriate empirical antibiotic treatment, ultrasonography-driven necrotising enterocolitis NEC therapy with antibiotic regimens including tigecycline., adequate combined treatment within 48 hours, empirically, antibiotic prophylaxis, Appropriate empirical antibiotics within 48 h, 72-h IIAT, prior antimicrobial exposure, inadequate antibiotic treatment, inadequate initial antimicrobial therapy, received antibiotherapy with Sulfamethoxazole / trimethoprim, received appropriate treatment for *S. maltophilia*, timing of appropriate antibiotherapy, anti-staph treatment < 48 h, cefepime treatment, appropriate empirical antimicrobial therapy, inappropriate treatment, delay for appropriate treatment, antibiotic lock therapy, initial adequate therapy, inappropriate empirical therapy., prior antifungal use, inappropriate empiric antibiotics, empirical treatment (No active drug, inappropriate empirical therapy, appropriate definitive therapy |
| Basic characteristics | Paediatric (Compared to adults), sex, age, Age years, Age >60, Age > 65 years, Age>45, Age, male gender, Age <60, Age, Ethnicity, Older age (>65 years old), age, sex, Age, sex, Age <60 years vs. Age >60 years, Clinical model: Demographic data (age), Smoking history (Never smoker/Former smoker/Current smoker), Age, Age, sex, Haematological malignancies: age, sex, Solid tumours: age, sex, Early case fatality: age, sex, Overall case fatality: age, sex, Age, gender, sex, age, Age (>60 vs. <60), year of diagnosis, age, Age group, sex, Sex, age, Sex, age, age >55 yr, Sex, age > 65 years, age, male gender, age at diagnosis, Age > 60 years, Age >60, sex, Age>55 years old, Age group, Model 2: age <65 years, 14 days mortality models 1 and 2 : age >65 years old, 30 days mortality models 1 and 2 : sex, age >65 years old, age, sex, Age, gender, age <65, gender, race, sex, age less than 18 years, Age, age 65 and over, age, Age, race, gender, age, sex, sex, age, age>55, sex |
| Cancer-related | type of malignancies, lung cancer, prostate cancer, advance neoplasm, adverse genetic group acute myeloid leukemia, Complete remission, relapsed/refractory disease, metastatic cancer, solid tumor/Hematologic malignancy, recent diagnosis (vs relapse), Performance status >2, site (respiratory vs other), fatal prognosis according to McCabe index, community site of acquisition, BCLC stage C or D vs. A or B, ALBI grade 3 vs. grade 1 or 2, Solid tumor vs Hematological, Recent diagnosis or remission vs Progression/re-lapse, Existence of distant metastasis, Primary sites of infection (Respiratory tract/Urinary tract/BSI), Underlying leukemia, leukemia, lymphoma, refractory or recurrent disease, primary disease acute myeloid leukemia, remission, Myelodysplastic syndrom, advanced neoplasm, advanced neoplasm, hematological malignancies, hematological malignancies, Nephroblastoma, neuroblastoma, sarcomas, Performance status score, tumor lysis, acute myeloid leukemia, acute myeloid leukemia, 14-day mortality model: solid tumor, cytology Poor vs. Favorable, cytology Poor vs. Intermediate, cytology Poor vs. Unknown, AML/ALL intermediate, AML/ALL/MDS advanced, MDS advanced, Karnofsky performance scale <90, Hodgkin's lymphoma, Myelodysplastic syndrome, Solid neoplasia, haematological malignancy (Compared to solid tumours), solid tumor, malignancy-related complications, pancreatic tumour, biliary tumour, malignancy-related complications, underlying malignancies, disease relapsed or uncontrolled, relapsed or uncontrolled malignancy, acute myeloid leukemia, non Hodgkin's lymphoma, Hodgkin's lymphoma, cancer with metastasis, hematologic malignancy, Hematological malignancies, acute leukemia, acute myeloid leukemia, performance status, haematological malignancy, recurrent Hilar cholangiocarcinoma, acute myeloid leukaemia not in remission, malignancy type, hematologic malignancy, complete remission, solid tumor, hematological cancer, metastasis, complete remission of acute leukemia |
| Chemotherapy/immunosuppressants | type of the last chemotherapy treatment, day 15 bone marrow blast clearance, no recent chemotherapy (vs recent), sorafenib treatment, Chemotherapy, intensive chemotherapy. Model 2: Initial active treatment, intensive chemotherapy., High-dose cytarabine-containing chemotherapy, standard dose cytarabine chemotherapy, receiving chemotherapy, intensive Chemotherapy., previous chemotherapy within 30 days, induction or re-induction chemotherapy, chemotherapy |
| Comorbidities/clinical condition | APACHE-II score at onset of bacteraemia, Dependent functional capacity at baseline, pulmonary disease, liver disease, hæmotologiv stem cell transplant comorbidities index, renal failure, functional capacity at bacteraemia onset, comorbidities, type II diabetes mellitus, cerebrovascular disease, |
|  | congestive heart failure, APACHE II, SAPS II score >40, SOFA score <10/>10, chronic kidney disease, multiorgan failure, end organ disease, APACHE II score, acute kidney infection or acute renal failure, diabetes Mellitus, SOFA score, SOFA score at diagnosis, SOFA score at healthcare associated infection (HAI) diagnosis, acute kidney injury within 30 days after HAI diagnosis, diabetes mellitus, chronic liver disease, and comorbidities., underlying cirrhosis, Child–Pugh class C vs. A or B, ECOG performance status, Comorbidities (Liver disease), Charlson comorbidity index CCI (1–2), CCI≥3, Intrapleural/abdominal infusion (within 30 days), Signs of severity, qSOFA score ≥2, APACHE II score ≥19, hypotension (<90/50), CRI non hemorrhagic, Charlson comorbidity index I>3, renal Replacement Therapy, initial SOFA score, acute kidney injury, APACHE II score, Charlson Comorbidity index>3, Charlson scoring system, Charlson scoring system, comorbidities, chronic obstructive pulmonary disease, diabetes, hypertension, ECOG score binary, Charlson comorbidities index (CCI), number of organ dysfunction, chronic liver disease, HIV-infection. Adjusted for: chronic liver disease, diabetes mellitus, chronic renal failure, Charlson index, ECOG, Charlson index > 4 points, SOFA score, renal replacement therapy, SOFA score, ECOG 1, ECOG 2, ECOG 3, ECOG 4, diabetes mellitus, renal insufficiency, hematological disease newly diagnosed / relapsed after one remission, altered state of consciousness, acute renal failure, acute hepatic failure, comorbidities, qSOFA score, APACHE II score >20, APACHE II score >20, organ failure, charlson comorbidity index (CCI) >4, underlying disease., complications (hypoxia, SOFA score, Charlson comorbidity index, Charlson index score, the presence of complications or comorbidities (eg, hypertension, diabetes, Charlson score, cerebrovascular disease, dementia, Charleston Co-Morbidity Index, altered consciousness, acute renal failure, acute hepatic failure, hypertension, chronic heart disease, diabetes mellitus, organ failure and sepsis shock |
| Gastrointestinal | gastrointestinal symptoms |
| Hospital-related | location in hospital at onset of bacteraemia, inpatient, length of hospital stay in days before onset of bacteraemia, admission-days before bacteraemia, Hospital length of stay, Numbers of co-infected locations, hospital stay >30 days, Intensive care unit stay, intensive care unit admission for healthcare associated infection (HAI), admission status, Length of hospitalization (≥21.0), intensive care unit admission, hospital stay, intensive care unit stay, previous hospitalisation, intensive care unit, intensive care unit admission, previous hospital admission, intensive care unit entrance, Length of hospital stay, days of hospitalization, intensive care unit requirement., duration of stay in hospital, ICU admission, duration of hospitalization prior to bacteremia, intensive care unit stay, ICU stay, intensive care unit stay, length of stay, intensive care unit residence, ICU admission, teaching status of hospital, and the performance of bone marrow vs stem cell transplant during hospitalization., hospitalisation within 30 days prior to infection, admission to intensive care unit, Hospitalization, intensive care unit, presence of preceding bacteremia during the hospitalization, admission days, hospital region, hospital location, ICU at diagnosis, intensive care unit admission, LOS, ICU, duration of hospital stay. |
| Infection-related | shock at onset of bacteraemia), Clostridium difficile status (no infection/asymptomatic/symptomatic), type of bacteria, polymicrobial bacteraemia, CRGNB, septic shock, bacteremia origin (healthcare/nosocomial), clinical manifestations (Low grade fever (<38 °C/ Shock at presentation), colonization with carbapenem resistant enterococci, gram-negative bacterial infection, infection with coagulase negative Staphylococci, infection with Acinetobacter baumannii, infection with Klebsiella spp., infection with extended spectrum beta lactamase-producing bacteria, infection with Gram-negative multi drug resistant bacteria, catheterization focus of infection, isolation of Gram + bacteria in culture, isolation of Gram - bacteria in culture, extended spectrum beta lactamase Enterobacteriaceae, carbapenem resistance, septic shock, multi-resistant Gram-negative BSI, pneumonia, Hemograms of bloodstream infections (white blood cell, Pathogens for bloodstream infections (among them 11 bacteria, fungi, polymicrobial and other), gram negative organisms, gram positive organisms, polyorganisms, and fungi, positive culture of multi drug resistant organism, Source of infection (Pneumonia), polymicrobial bloodstream infection, shock, colonized/Infected, septic shock, concurrent bacterial sepsis, albicans vs non albicans, Biofilm (strong vs weak), colistine resistance, fungal infection, phenotype, multi drug resistant (MDR) vs non-MDR bloodstream infection (BSI), P. aeruginosa vs other BSI, strong vs weak biofilm-producer, septic shock, infection related mortality, septic shock rate, multi drug resistant-polymicrobial bloodstream infections mortality, septic shock, fever, catheter-related candidaemia, mixed bloodstream infection, caspofungin resistance, polymicrobial bacteremia, vaccine serotype VPV23, septic shock at diagnosis of XDR-ABC bacteraemia (extended drug resistant), days from 1st positive culture to initial treatment, concomitant infection, bloodstream infection, received effective therapy for Klebsiella producing carbapenemase-Kp infection, catheter as a source of bacteremia, pulmonary source of infection, intra abdominal source of infection, cONs, E.coli, P.aeruginosa, shock at onset, C. difficile infection, multi drug resistant organism-colonization, Gram-positive vs Gram-negative, Monomicrobial vs Polymicrobial, Non-multidrug resistant (MDR) vs MDR and/or extended spectrum beta lactamase, Gram negative bacteria, E.coli, K. pneumoniae, Enterobacter spp, Fungi, C. albicans. Candida spp, enterococcus, Septic shock, documented clinical source, breakthrough fungemia, prior vancomycin resistant enterococcus |
|  | bacteremia (VREB), persistent VREB >48 hours, CoNS (coagulase negative staphylococci). Mortality 7 days model : coNS, polymicrobial infection, previous isolation of *S. maltophilia*, Focus of infection catheter related infection (CRI), focus of infection hemorrhagic, previous colonization, coinfection, >15 days VRE, MASCC score <21, MASCC score <21, coagulase-negative staphylococci, multi drug resistant gram negative bacteria, septic shock., septic shock at onset, pneumonia, Exacerbation of interstitial pneumonia, interstitial pneumonia, microbiology with or without non-fermenting Gram(-) bacilli, extended spectrum beta lactamase enterobacteria (ESBL-E), multiple bacterial infections, bacteria/fungal coinfections, colonisation by *Klebsiella* producing carbapenemase, colonisation by *Acinetobacter*, colonisation by *Pseudomonas*, pre-chemo infection, shock, shock, catheter related infection, extended spectrum beta lactamase, catheter related infection, PITT bacteremia score, VRE bacteremia, other bloodstream infections, Vancomycin resistant enterococcus bloodstream infection, Pulmonary source, Abdominal source, Shock, Candidemia, HIV, catheter-related source, coagulase-negative staphylococci bacteremia, *S. pneumoniae* bacteremia, primary source of infection, catheter related source of infection, septic shock, *C. tropicalis*, *Candida albicans* (Compared to non-*C.albicans* or mixed), bacteraemia, shock, receiving Amphotericin B (Compared to those not receiving), after positive culture, MASCC risk score < 21, persistent bacteremia, septic Shock, fever >38C, multi drug resistant organism colonization (MDRO), multidrug resistant gram negative bloodstream infections (BSI), nonfermenter BSI, carbapenem resistant gram negative bacteria (CR - GNB), gram negative bacteria, pulmonary aspergillosis, ventilated at infection onset, Multi drug resistant organism colonisation (MDRO), Pitt score $\geq 4$, MASCC score <21, carbapenem resistant gram negative bacteria, non fermentative bacteria, septic shock, carbapenem resistant by *Klebsiella pneumoniae*, type of bacteremia, caspofungin non-susceptibility, multidrug resistance, caspofungin non-susceptibility, multidrug resistance, Bacterial multi drug resistant infection, coexistence of fungal infection, clinically documented infection, Abdominal infection, unknown origin of infection, septic shock, extended spectrum beta lactamase carriage, carbapenem resistant enterobacteriaceae carriage, abdominal source, Meticillin-resistant *Staphylococcus aureus* (MRSA), infectious endocarditis, Pitt bacteremia score, extended spectrum beta lactamase-positive bacteremia, Pitt bacteremia score $\geq 4$, *C. difficile* infection., septic shock etc.), *A.baumannii*, polymicrobial infection, pneumonia, breakthrough bacteremia occurrence within 48 h of ongoing antibiotics, multi drug resistant *E.coli*, *Candida* spp Species, *C. parapsilosis*, abdominal infection, Bloodstream infection, *C. difficile* infection, isolation of vancomycin resistant enterobacteriaceae (VRE) in the prior 1 year, isolation of extended spectrum beta lactamase (ESBL) in the prior 1 year, isolation of CRAB in the prior 1 year, history of GNB bacteremia in the prior 1 year, pneumonia, Pitt score, septic shock, carba-non sensitive, days from chemotherapy to GNB bacteremia, hospital days to GNB bacteremia., sepsis, severity of sepsis (Sepsis, Severe sepsis or septic shock), *P. aeruginosa* infection, Pitt bacteremia score, septic shock, 3rd generation cephalosporins resistance by *E. coli* isolate., ESBL, perianal infection, pulmonary infection, CR-PA, MDR-PA |
| Laboratory findings (non-microbiological) | disseminated Intravascular Coagulation (DIC) score, systolic blood pressure, diastolic blood pressure, Hemoglobin, platelet), Serum albumin, Serum albumin, Alpha-fetoprotein > 400 ng/ml, Laboratory examination results (Hemoglobin g/L <110.0, Platelet count $\times 10^9/L$ <100.0, Lymphocytes count $\times 10^9/L$ <1.0, PCT ng/mL $\geq 1.0$, Albumin g/L <30.0), hypoalbuminemia, T cell depletion, platelet count, median albumin level, median Albumin level, histology with or without squamous cells, Albumin to Globulin Ratio, C reactive protein, prothrombin activity, D-Dimer, creatinin, albumin, alkaline phosphatase, total bilirubine, blood lactate level (mmol/L), creatinine, Human leucocyte antigen 7/8, C reactive protein (CRP), Model 2: creatinine, CRP, Model 5: CRP, platelet count, creatinine level, histology: squamos cell carcinoma, histology: others, platelet < $10 \times 10^3 \text{ mm}^{-3}$, hemoglobin <70g/dl, platelet < $10 \times 10^3 \text{ mm}^{-3}$, albumin <30g/L, AST >120U/L, total bilirubin >34.2$\mu\text{mol/L}$, prothrombin time >14s, glucose > 140 mg/dL, Nadir WBC count |
| Neutropenia | prolonged neutropenia prior to infection, Absolute neutrophil count, duration of neutropenia (days), Duration of neutropenia before blood culture, Neutropenia at the onset of bloodstream infection, severe neutropenia at the onset of bloodstream infection (<100/mm<sup>3</sup>), duration of neutropenia >21 days, neutrophil recovery, monocytopenia, neutropenia past month, current neutropenia, severe neutropenia past month, current severe neutropenia, febrile neutropenia, prolonged neutropenia, Neutrophils >500 vs Neutrophils 500, persistent neutropenia, neutropenia, $5 \times 10^9/L$ neutrophil count while vancomycine resistant enterococcus (VRE) +, absolute monocyte count <100, monocytic acute myeloid leukemia, monocytes, 30-day mortality model: Neutropenia, Model 1: neutropenia, absolute neutrophile count 500 or more, ANC < 500/$\mu\text{l}$ duration until BSI (days), neutrophil count, absolute neutrophil count, ANC <100/mm<sup>3</sup>, ANC <500/mm<sup>3</sup>, neutropenia, duration of neutropenia before infection, ANC less than 100 cells/mm<sup>3</sup>, neutropenia at any time., neutropenia, duration of neutropenia before BSI |
| Other | others(OPD/ ER), disease risk (favourable/intermediate/high), nasogastric tube, Disease status, combination therapy, mucositis, mucositis, < 0, DM, , nasogastric tube, etiology (secondary vs. de novo), white blood cell at diagnosis, Time period, prior therapies >2, Initial low tidal volume |
| | ventilation, status of disease, prone positioning treatment, extensive diagnosis, Indeterminable, type of CT last 3 months, palliative care, hemodynamic instability, adherence to protocol, etc.), skin lesion, dissemination, mean (± SD), At least one active drug), definitive treatment (One active drug, More than one active drugs)., BF |
| Other treatments/medications | current corticosteroid therapy, steroid administration, corticosteroids, steroid exposure, corticosteroid therapy, corticosteroid therapy, corticosteroid therapy, corticosteroid therapy., administration of steroid, radiotherapy dose in Gy, aciclovir, anthracycline, duration of vasopressors, hemodialysis, hemodialysis, corticosteroid use, receiving steroids, corticosteroid therapy, corticosteroid therapy, vasopressors treatment, use of vasopressors, vasopressors, vasopressors, previous radiation within 30 days, corticosteroid |
| Respiratory | Co-infections (respiratory), mechanical ventilation, positive culture in blood/respiratory tract/urinary tract/skin and soft tissue, respiratory symptoms, mechanical ventilation, mechanical ventilation, chronic lung disease, intubation, mechanical ventilation, bronchial stump fistula, mechanical ventilation, mechanical ventilation, respiratory source, severe acute respiratory distress syndrome, acute respiratory failure, acute respiratory failure, acute respiratory failure, mechanical ventilation, mechanical ventilation, mechanical ventilation, avute respiratory failure, mechanical ventilation |
| Surgery-related | emergent surgery, removal of invasive devices within the first 48 h, removal of invasive device or control deep infection, resection as hepatocellular carcinoma treatment, Surgery, presence of Drains postoperation; Infection-related model: Sample type (Urine/Blood culture), invasive procedures, invasive procedures, previous surgery within 30 days, invasive procedure, surgery |
| Transplantation-related | donor type (related/unrelated), human Leukocyte Antigen compatibility (match/mismatch), stem cell transplantation as consolidation therapy, prior hæmatologic stem cell transplant, Mortality 30 days model : Auto hematologic stem cell transplant treatment, Cord blood allograft, cytomegalovirus donor or receptor positive, hematologic cell transplant during 2010-2012, hematologic stem cell transplant HSCT, autologous HSCT, Post-transplant C. difficile infection, history of hemotologic stem cell transplantation, autologous transplant |
| Urinary | urinary catheter, source of bacteraemia (Urinary tract), urinary tract infection, Presence of indwelling urinary catheters, Model 1: Klebsiella producing carbapenemase-K. pneumoniae (KPC-KP) bloodstream infection (BSI) developing during inactive antibiotic treatment, KPC-KP BSI developing during inactive antibiotic treatment, foley catheter, urinary catheter at infection onset, urine tube, Urinary tract infection |
| Intravascular access | central catheterization, central venous catheter-related bloodstream infection (BSI), central venous catheter removal, central venous catheter removal within 48 h after blood culture collection, central venous catheter, central venous catheter removal within 48 hours, receiving total parenteral nutrition, blood transfusion, Model 1: central venous catheter removal, previous blood transfusion, catheter removal, CVC duration, CVC related candidemia, removal of CVC., Intravenous group, previous blood transfusion |

## Supplementary material 7

### PRISMA 2020 Main Checklist

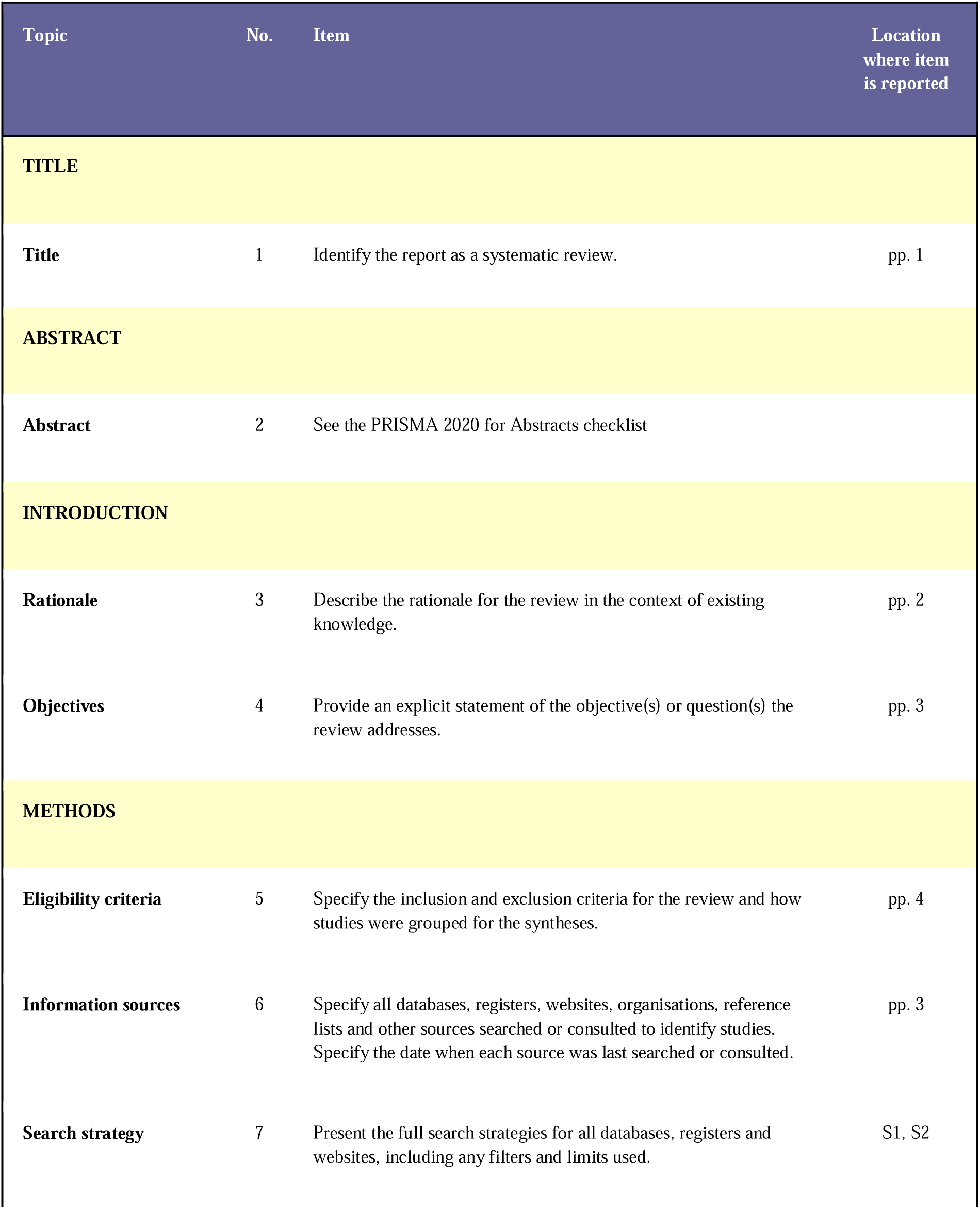
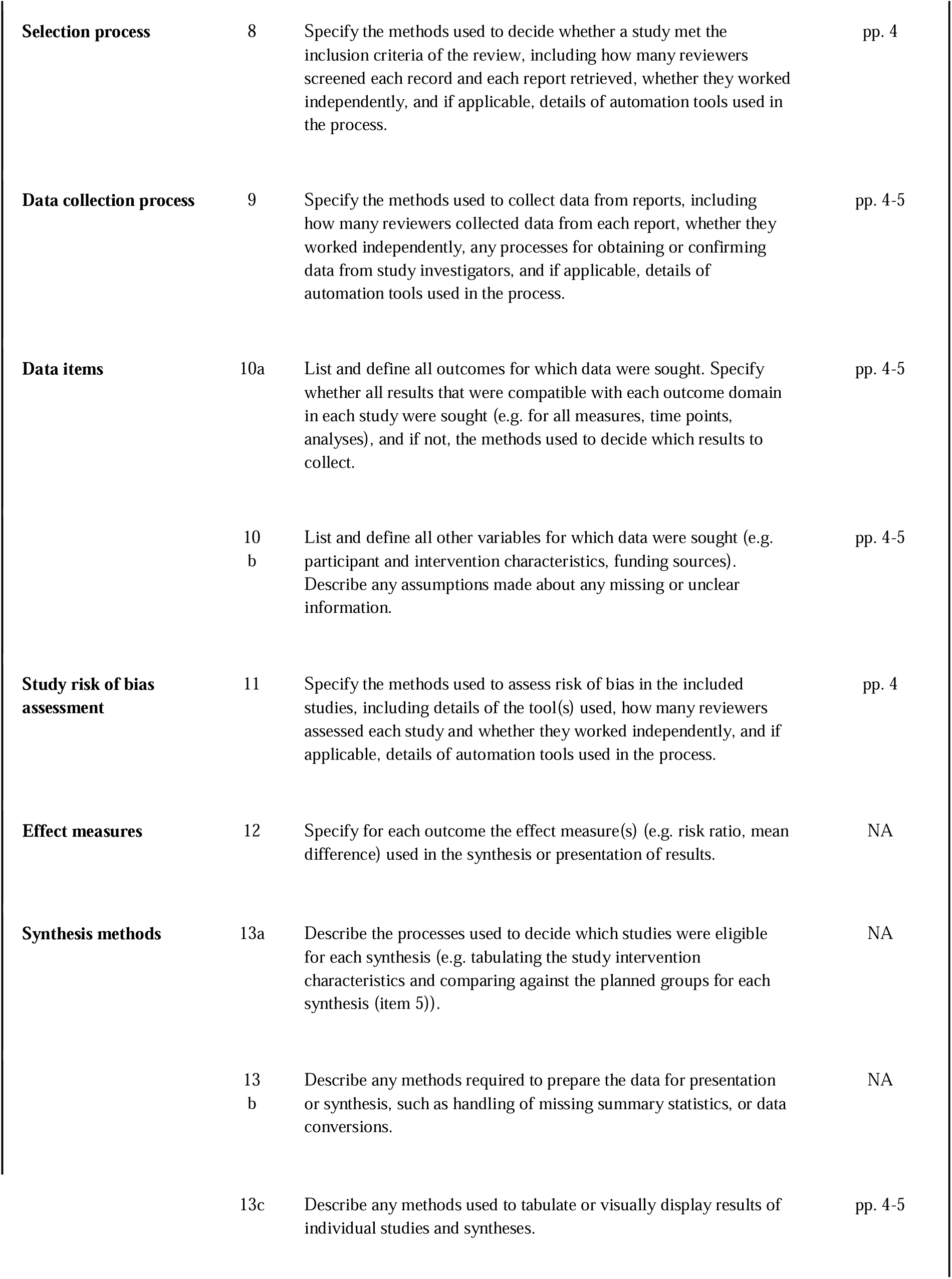
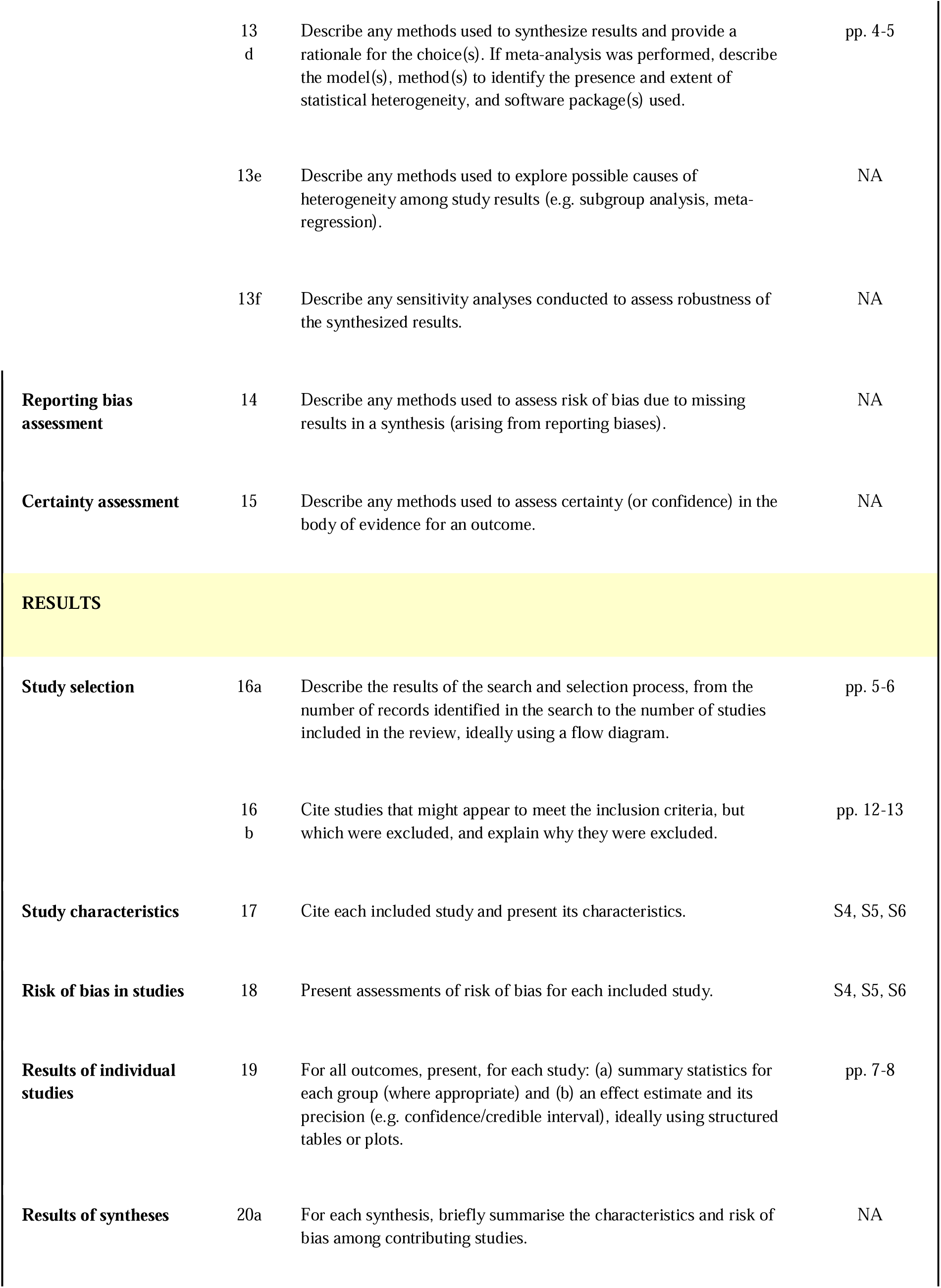
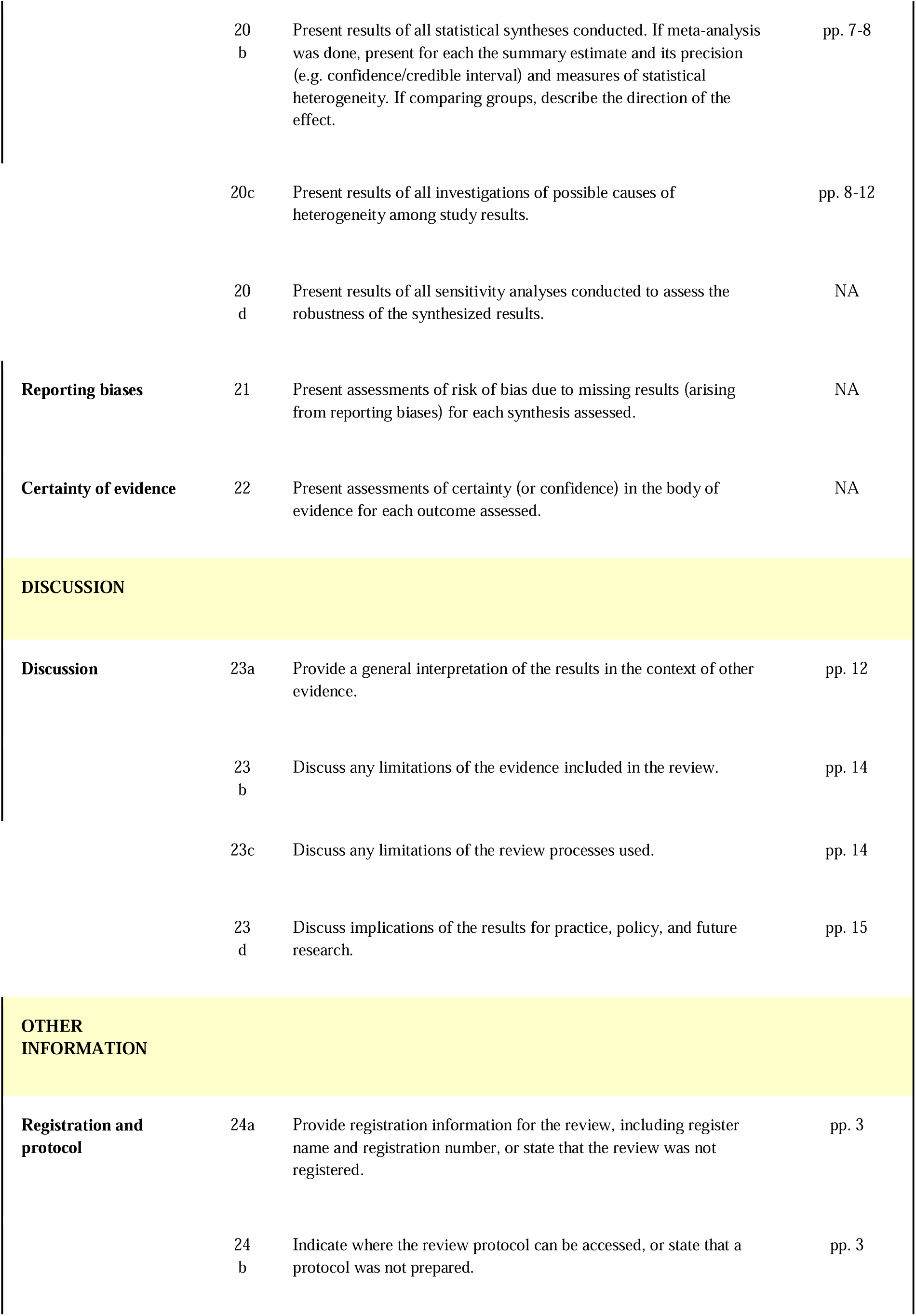
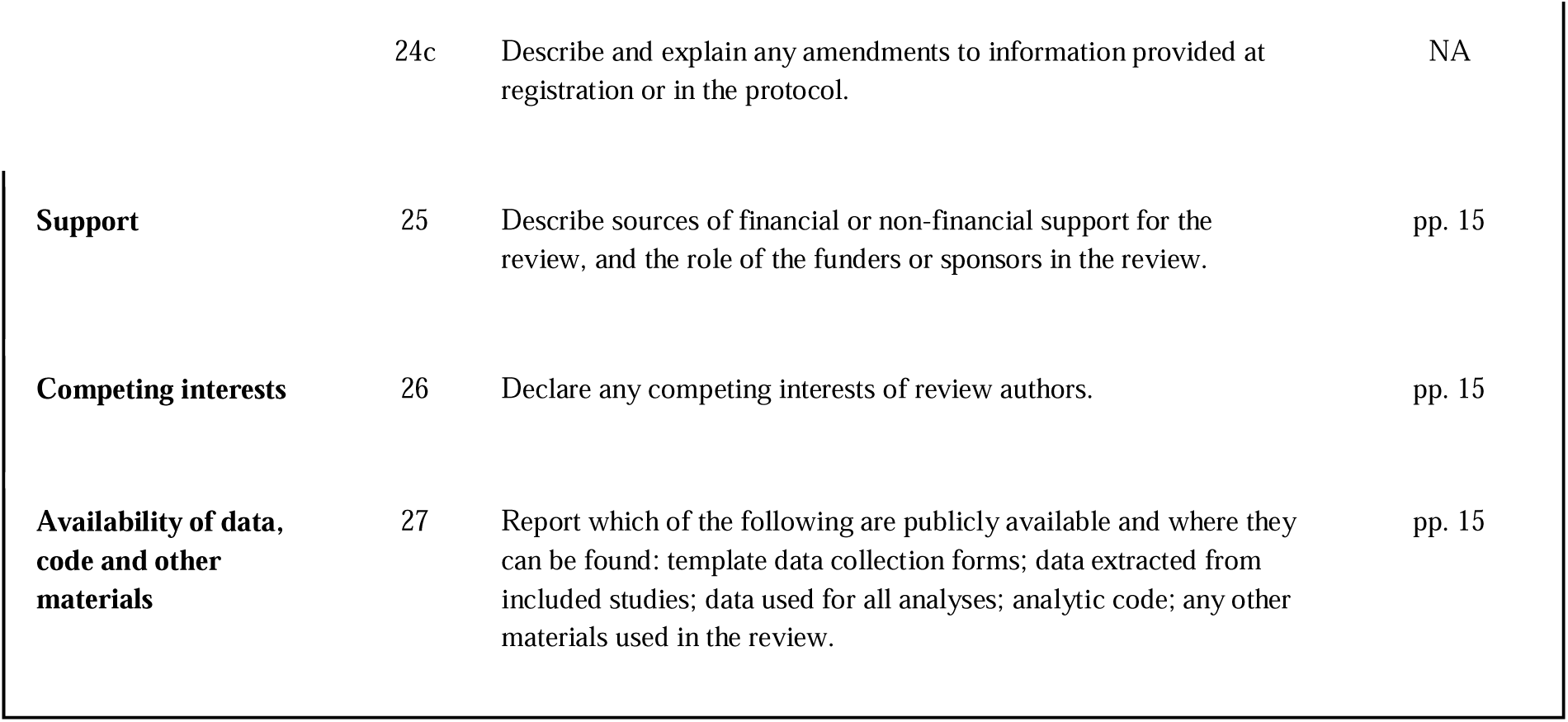

Supplementary material 7 PRISMA 2020 Main Checklist
| Topic | No. | Item | Location where item is reported |
| --- | --- | --- | --- |
| **TITLE** |  |  |  |
| **Title** | 1 | Identify the report as a systematic review. | pp. 1 |
| **ABSTRACT** |  |  |  |
| **Abstract** | 2 | See the PRISMA 2020 for Abstracts checklist |  |
| **INTRODUCTION** |  |  |  |
| **Rationale** | 3 | Describe the rationale for the review in the context of existing knowledge. | pp. 2 |
| **Objectives** | 4 | Provide an explicit statement of the objective(s) or question(s) the review addresses. | pp. 3 |
| **METHODS** |  |  |  |
| **Eligibility criteria** | 5 | Specify the inclusion and exclusion criteria for the review and how studies were grouped for the syntheses. | pp. 4 |
| **Information sources** | 6 | Specify all databases, registers, websites, organisations, reference lists and other sources searched or consulted to identify studies. Specify the date when each source was last searched or consulted. | pp. 3 |
| **Search strategy** | 7 | Present the full search strategies for all databases, registers and websites, including any filters and limits used. | S1, S2 |
| **Selection process** | 8 | Specify the methods used to decide whether a study met the inclusion criteria of the review, including how many reviewers screened each record and each report retrieved, whether they worked independently, and if applicable, details of automation tools used in the process. | pp. 4 |
| **Data collection process** | 9 | Specify the methods used to collect data from reports, including how many reviewers collected data from each report, whether they worked independently, any processes for obtaining or confirming data from study investigators, and if applicable, details of automation tools used in the process. | pp. 4-5 |
| **Data items** | 10a | List and define all outcomes for which data were sought. Specify whether all results that were compatible with each outcome domain in each study were sought (e.g. for all measures, time points, analyses), and if not, the methods used to decide which results to collect. | pp. 4-5 |
|  | 10b | List and define all other variables for which data were sought (e.g. participant and intervention characteristics, funding sources). Describe any assumptions made about any missing or unclear information. | pp. 4-5 |
| **Study risk of bias assessment** | 11 | Specify the methods used to assess risk of bias in the included studies, including details of the tool(s) used, how many reviewers assessed each study and whether they worked independently, and if applicable, details of automation tools used in the process. | pp. 4 |
| **Effect measures** | 12 | Specify for each outcome the effect measure(s) (e.g. risk ratio, mean difference) used in the synthesis or presentation of results. | NA |
| **Synthesis methods** | 13a | Describe the processes used to decide which studies were eligible for each synthesis (e.g. tabulating the study intervention characteristics and comparing against the planned groups for each synthesis (item 5)). | NA |
|  | 13b | Describe any methods required to prepare the data for presentation or synthesis, such as handling of missing summary statistics, or data conversions. | NA |
|  | 13c | Describe any methods used to tabulate or visually display results of individual studies and syntheses. | pp. 4-5 |
|  | 13d | Describe any methods used to synthesize results and provide a rationale for the choice(s). If meta-analysis was performed, describe the model(s), method(s) to identify the presence and extent of statistical heterogeneity, and software package(s) used. | pp. 4-5 |
|  | 13e | Describe any methods used to explore possible causes of heterogeneity among study results (e.g. subgroup analysis, meta-regression). | NA |
|  | 13f | Describe any sensitivity analyses conducted to assess robustness of the synthesized results. | NA |
| **Reporting bias assessment** | 14 | Describe any methods used to assess risk of bias due to missing results in a synthesis (arising from reporting biases). | NA |
| **Certainty assessment** | 15 | Describe any methods used to assess certainty (or confidence) in the body of evidence for an outcome. | NA |
| **RESULTS** |  |  |  |
| **Study selection** | 16a | Describe the results of the search and selection process, from the number of records identified in the search to the number of studies included in the review, ideally using a flow diagram. | pp. 5-6 |
|  | 16b | Cite studies that might appear to meet the inclusion criteria, but which were excluded, and explain why they were excluded. | pp. 12-13 |
| **Study characteristics** | 17 | Cite each included study and present its characteristics. | S4, S5, S6 |
| **Risk of bias in studies** | 18 | Present assessments of risk of bias for each included study. | S4, S5, S6 |
| **Results of individual studies** | 19 | For all outcomes, present, for each study: (a) summary statistics for each group (where appropriate) and (b) an effect estimate and its precision (e.g. confidence/credible interval), ideally using structured tables or plots. | pp. 7-8 |
| **Results of syntheses** | 20a | For each synthesis, briefly summarise the characteristics and risk of bias among contributing studies. | NA |
|  | 20b | Present results of all statistical syntheses conducted. If meta-analysis was done, present for each the summary estimate and its precision (e.g. confidence/credible interval) and measures of statistical heterogeneity. If comparing groups, describe the direction of the effect. | pp. 7-8 |
|  | 20c | Present results of all investigations of possible causes of heterogeneity among study results. | pp. 8-12 |
|  | 20d | Present results of all sensitivity analyses conducted to assess the robustness of the synthesized results. | NA |
| **Reporting biases** | 21 | Present assessments of risk of bias due to missing results (arising from reporting biases) for each synthesis assessed. | NA |
| **Certainty of evidence** | 22 | Present assessments of certainty (or confidence) in the body of evidence for each outcome assessed. | NA |
| **DISCUSSION** |  |  |  |
| **Discussion** | 23a | Provide a general interpretation of the results in the context of other evidence. | pp. 12 |
|  | 23b | Discuss any limitations of the evidence included in the review. | pp. 14 |
|  | 23c | Discuss any limitations of the review processes used. | pp. 14 |
|  | 23d | Discuss implications of the results for practice, policy, and future research. | pp. 15 |
| **OTHER INFORMATION** |  |  |  |
| **Registration and protocol** | 24a | Provide registration information for the review, including register name and registration number, or state that the review was not registered. | pp. 3 |
|  | 24b | Indicate where the review protocol can be accessed, or state that a protocol was not prepared. | pp. 3 |
|  | 24c | Describe and explain any amendments to information provided at registration or in the protocol. | NA |
| **Support** | 25 | Describe sources of financial or non-financial support for the review, and the role of the funders or sponsors in the review. | pp. 15 |
| **Competing interests** | 26 | Declare any competing interests of review authors. | pp. 15 |
| **Availability of data, code and other materials** | 27 | Report which of the following are publicly available and where they can be found: template data collection forms; data extracted from included studies; data used for all analyses; analytic code; any other materials used in the review. | pp. 15 |

### PRISMA Abstract Checklist

| Topic | No. | Item | Reported? |
| --- | --- | --- | --- |
| **TITLE** |  |  |  |
| **Title** | 1 | Identify the report as a systematic review. | Yes |
| **BACKGROUND** |  |  |  |
| **Objectives** | 2 | Provide an explicit statement of the main objective(s) or question(s) the review addresses. | Yes |
| **METHODS** |  |  |  |
| **Eligibility criteria** | 3 | Specify the inclusion and exclusion criteria for the review. | Yes |
| **Information sources** | 4 | Specify the information sources (e.g. databases, registers) used to identify studies and the date when each was last searched. | Yes |
| **Risk of bias** | 5 | Specify the methods used to assess risk of bias in the included studies. | Yes |
| **Synthesis of results** | 6 | Specify the methods used to present and synthesize results. | Yes |
| **RESULTS** |  |  |  |
| **Included studies** | 7 | Give the total number of included studies and participants and summarise relevant characteristics of studies. | Yes |
| **Synthesis of results** | 8 | Present results for main outcomes, preferably indicating the number of included studies and participants for each. If meta-analysis was done, report the summary estimate and confidence/credible interval. If comparing groups, indicate the direction of the effect (i.e. which group is favoured). | Yes |
| **DISCUSSION** |  |  |  |
| **Limitations of evidence** | 9 | Provide a brief summary of the limitations of the evidence included in the review (e.g. study risk of bias, inconsistency and imprecision). | No |
| **Interpretation** | 10 | Provide a general interpretation of the results and important implications. | Yes |
| **OTHER** |  |  |  |
| **Funding** | 11 | Specify the primary source of funding for the review. | Yes |
| **Registration** | 12 | Provide the register name and registration number. | Yes |

